# Altered Sensorimotor Neuroplasticity in the Subacute Period Following Burn Injury

**DOI:** 10.1101/2025.10.16.25338168

**Authors:** Grant Rowe, Mark Fear, Dale W. Edgar, Aleksandra Miljevic, Tyler Osborne, Natalie Morellini, Merrilee Needham, Ann-Maree Vallence, Fiona Wood

**Author notes:** **Corresponding author:** Grant Rowe, School of Psychology, College of Health and Education, Murdoch University, Australia, Murdoch University, 90 South Street, Murdoch, Perth, Australia 6150. **Author Contribution Statement:** All authors meet the criteria for authorship as stated in the Uniform Requirements for Manuscripts Submitted to Biomedical Journals. All authors have made substantial contributions to all the following: (1) the conception and design of the study, or acquisition of data, or analysis and interpretation of data, (2) drafting the article or revising it critically for important intellectual content, (3) final approval of the version to be submitted. Specifically, MF, DE, MN, AMV, and FW primarily designed the study. GR and AMV led the drafting and editing of the manuscript. GR and AMV designed the statistical analysis. GR, AM, TO, and NM collected the data, while GR collated and cleaned the dataset. All authors assisted with the interpretation of results and editing of the final manuscript.

## Abstract

**Purpose:** Burn injuries affect the central nervous system, and evidence suggests they induce changes in cortical excitability related to motor dysfunction. The current study investigated markers of neuroplasticity in the motor cortex and motor performance in the subacute period following burn injury.

**Methods:** Thirty-four patients with a minor burn injury (total body surface area: 1.0 ± 2.0%; 16 females; age: 46.0 ± 14.0 years) participated in a longitudinal study with assessments ranging from ∼1 to ∼17 weeks post-injury. Transcranial magnetic stimulation (TMS) was used to evaluate motor cortex excitability and short- and long-intracortical inhibition (SICI; LICI) before and after application of paired associative stimulation (PAS) to induce neuroplasticity. Motor performance was assessed using the Purdue Pegboard.

**Results:** Patients showed improvements in bilateral motor performance from ∼3 to ∼6 weeks post-injury. PAS did not induce any change in net motor cortex excitability or SICI but increased LICI, which was greatest ∼6 weeks post-injury. Adjusted modelling showed that greater PAS-induced increase in LICI was associated with better bilateral motor performance, suggesting an association between PAS-induced neuroplasticity of GABA_B_-mediated inhibition and motor function following burn injury.

**Conclusions:** These findings highlight a potential role for neuroplasticity of inhibitory circuits in the motor cortex in recovery of motor function following burn injury. The data provide a neurophysiological basis to test whether targeting inhibitory networks in the subacute period after burn injury can enhance or expedite functional recovery.

## Introduction

Burn injuries cause extensive tissue damage, affecting not only the skin but also the peripheral neural architecture. Such changes can result in motor and sensory impairments that significantly diminish quality of life (1), which have been reported in both severe (>20% total body surface area (TBSA); (2)) and non-severe burns (<20% TBSA; (3)). Sensory and motor impairments following burn injury are likely mediated by multiple, interacting factors, including systemic inflammation (4, 5), sensory dysfunction (6, 7), and pain (8, 9), all of which may influence brain regions involved in motor control and affect functional outcomes (10). While much research has explored the functional consequences of burn injuries, increasing attention is now being directed toward the mechanisms underlying these impairments, particularly those involving the brain (11, 12). Neuroplasticity—the brain’s capacity to adapt in response to environmental stimuli and physiological change (13)—might play a key role in the functional impairments observed in burn patients. As such, it presents a potential target for therapeutic interventions to enhance recovery (11, 12).

Transcranial magnetic stimulation (TMS) is a valuable tool for studying neuroplasticity (14, 15) as it can be used to both measure and induce neuroplasticity. By generating a current in the brain via electromagnetic induction, TMS can depolarise neurons within a targeted cortical region (14). When a suprathreshold TMS pulse is applied to the primary motor cortex (M1), it produces a motor evoked potential (MEP), which can be recorded using electromyography (EMG) of the target muscle (15, 16). The peak-to-peak MEP amplitude provides a marker of corticospinal excitability, and changes in MEP amplitude indicate neuroplasticity (17, 18).

Single- and paired-pulse TMS can also be used to measure inhibition in the motor cortex. Single-pulse TMS can be used to measure the cortical silent period (cSP): a period of inactivity in the EMG from a voluntarily contracted muscle following a suprathreshold TMS pulse (19, 20). The early part of the cSP (<50 ms) is primarily mediated by spinal mechanisms and the later part (50–200 ms) is mediated by cortical mechanisms (21). The cSP is mediated by GABAergic inhibition, with both GABA_A_ and GABA_B_ receptor activity possibly playing a role (22–26). The ability to measure cortical inhibitory processes in conscious humans is important because cortical inhibition influences induction of neuroplasticity (27). Paired-pulse TMS can be used to measure GABAergic inhibition: when a subthreshold conditioning stimulus precedes a suprathreshold test stimulus by 1–5 ms, short latency and short-lasting inhibition of MEP amplitude is observed, known as short-interval intracortical inhibition (SICI) (28, 29). Pharmacological studies provide strong evidence that SICI is mediated by GABA_A_ receptor activity (30, 31).

Recently, TMS has been used to investigate neurophysiological changes in the motor cortex following burn injury. Garside et al. (32)—who tested burn patients from ∼6 weeks to 8 years post-injury—reported no overall difference in cortical silent period (cSP) duration compared with non-injured controls. However, exploratory subgroup analyses revealed shorter cSP durations on the burn-affected side in individuals with upper-limb burns, within two years post-injury, with less than 10% total body surface area (TBSA) affected, and those with partial-thickness burns—suggesting that non-severe burn injuries may reduce intracortical inhibition. Extending this work, recent exploratory analyses in individuals 1–3 years post-injury showed that baseline SICI (and long-interval intracortical inhibition: LICI) were significantly associated with bilateral motor performance on the Purdue Pegboard task (33). Greater inhibition at baseline was linked to better manual dexterity in burn patients, a relationship not observed in non-injured controls (33). These findings indicate that intracortical inhibition plays a role in upper limb motor control and may influence the extent of functional recovery following burn injury. In contrast, Whife et al. (34) reported no differences in SICI between burn patients (6–12 weeks post-injury) and controls, suggesting that intracortical inhibition may not be altered within this early subacute phase. However, as those assessments were not conducted earlier than ∼6 weeks post-injury, it is possible that inhibitory changes occurred sooner—closer to the time of injury—when physiological disruption is greatest (35). Together, these findings underscore the importance of early assessment to identify neuroplastic changes that may influence functional recovery trajectories in subacute burn patients.

When applied repetitively, TMS can induce neuroplasticity through long-term potentiation-(LTP) and long-term depression-(LTD) like mechanisms (36–38). Repetitive TMS (rTMS) produces lasting, bidirectional changes in MEP amplitude, with the direction determined by stimulation frequency and pattern; these effects are modulated by NMDA-receptor activity (39, 40). Recent work has examined whether markers of rTMS-induced of neuroplasticity are associated with functional outcomes following burn injury. Whife et al. (16) applied continuous theta burst stimulation (cTBS) to the M1 in subacute burn patients and showed a significant correlation between neuroplasticity induced by spaced cTBS at 12 weeks post-burn and general health outcomes, measured on the Short-form 36 (SF36) survey. Participants who exhibited the greatest neuroplastic responses to cTBS reported the highest general health scores. These findings suggest that M1 neuroplasticity may be a crucial factor in promoting functional recovery in burn patients. Together, these studies offer preliminary evidence of neuroplastic changes in the motor cortex following burn injury. It is, therefore, important to understand the time course and functional relevance of these changes following injury to optimise timely rehabilitation interventions and enhance functional recovery.

The aims of the current study were to investigate: (a) motor performance; (b) neuroplasticity induced by paired associative stimulation (PAS); and (c) intracortical inhibition within the motor cortex of individuals recovering during the subacute phase of burn injury (∼3–12 weeks post-injury). We hypothesised that pegboard performance would be poorer in the burns group than controls at ∼3 weeks post-injury and would improve steadily across recovery (∼6 and 12 weeks). We also hypothesised that baseline intracortical inhibition and PAS-induced neuroplasticity would be altered in the burns group at ∼3 weeks and that these differences would persist across the experimental protocol.

## Methods

### Participants

Thirty-four subacute burn patients were recruited through the Western Australian Adult Burn Unit databases at Fiona Stanley Hospital in Perth, Australia. Inclusion criteria included adult participants (age 18 years or older) with a TBSA injury of less than 20%, a burn injury sustained no more than 8 weeks prior to enrolment in the study, no previous burn injuries, MEP elicited by single-pulse TMS, and flame, scald, contact, friction, chemical, or electrical burn. After participants provided their informed consent, they completed a 17-item TMS safety questionnaire, which was used to screen and exclude individuals with contraindications to TMS (41–43). The study followed the Declaration of Helsinki guidelines and received ethical approval from the Murdoch University Human Research Ethics Committee (2021/047) and the South Metropolitan Health Service (RGS0000004279).

### Experimental Protocol

Figure 1 shows the experimental protocol. Participants who passed screening completed testing at three timepoints: (1) within five weeks post-injury, (2) ∼6 weeks post-injury, and (3) ∼12 weeks post-injury. The first timepoint occurred a median of 19 days post-injury (range: 4–32 days); the second, 44 days (25–66 days); and the third, 86 days (69–124 days). Twenty-two participants completed all three sessions; four completed <5 weeks and ∼6 weeks; and eight completed ∼6 and ∼12 weeks. Days since injury was treated as a continuous variable across the observed range (4–124 days). Model-based margins were taken from the full model, and contrasts were evaluated at 23, 44, and 82 days (∼3, 6, and 12 weeks) to align with clinical milestones and the densest sampling; these are analysis anchors, not fixed session time points. Each session lasted ∼2.5 hours and included neurophysiological testing and motor and sensory function assessments. Burn-specific and health-related well-being surveys were distributed electronically and completed within 24 hours either side of each session.

**Figure 1.**
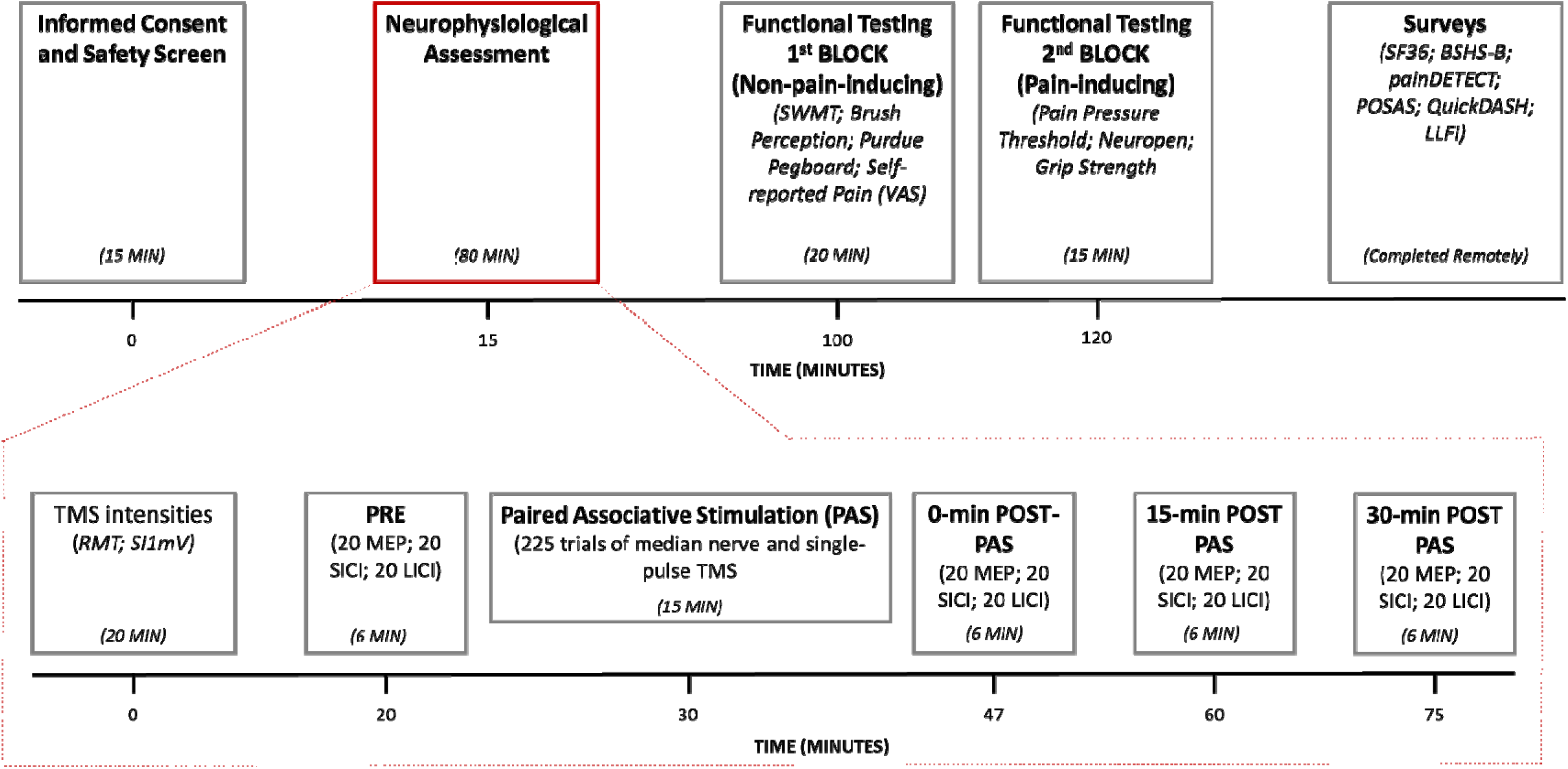
Timeline of the experimental protocol. SWMT: Semmes-Weinstein Monofilament Test; VAS: visual analogue scale; SF36: Short Form-36; BSHS-B: Burn Specific Health Scale-Brief; LLFI: Lower Limb Functional Index; TMS: transcranial magnetic stimulation; RMT: resting motor threshold; SI_1mv_: stimulus intensity required to elicit MEPs with a mean peak-to-peak amplitude of ∼1 mV; MEP: motor evoked potential; SICI: short-interval intracortical inhibition; LICI: long-interval intracortical inhibition. This figure is reproduced from earlier work (33).

### Neurophysiological Testing

#### Electromyography (EMG)

Neurophysiological testing was conducted with participants seated comfortably with their arms supported by a pillow. EMG signals were predominantly recorded from the first dorsal interosseous (FDI) using Ag-AgCl surface electrodes in a belly-tendon configuration (44). However, due to burn location over the FDI, one participant was recorded over the abductor digiti minimi (ADM), and another over the extensor digitorum muscle of the forearm (44). The EMG signals were amplified (x1000) using a CED 1902, bandpass filtered (20–1000 Hz), and digitised at 5 kHz with a CED 1401 (Cambridge Electronic Design, UK). EMG data were collected from the hand (or arm) on the burn-affected side. If both sides were affected by the burn, the side with greater burn surface area and depth was identified as the burn-affected side for the purpose of testing. Hand dominance was determined by asking participants which hand they use for writing. Eighteen participants were tested on their left dominant hemisphere (right-handed), 13 on their right non-dominant hemisphere (right-handed), 2 on their right dominant hemisphere (left-handed), and 1 on their left non-dominant hemisphere (left-handed).

#### Transcranial Magnetic Stimulation (TMS)

TMS was delivered using two MagStim 200^2^ stimulators connected via a MagStim BiStim module (Magstim, UK) to generate single and paired pulses. Monophasic pulses were administered through a 90-mm figure-of-eight coil positioned tangentially to the scalp. The coil handle was angled backward and rotated ∼45° from the midline to induce a posterior-anterior current in M1, targeting the hemisphere contralateral to the most-affected hand. The optimal M1 stimulation site was determined as the location that consistently produced the largest motor evoked potentials (MEPs) in the target muscle (43, 45). This site was marked on a skull cap to ensure consistent coil placement throughout testing. The resting motor threshold (RMT) was defined as the lowest stimulus intensity, expressed as a percentage of the maximum stimulator output (MSO), that elicited MEPs ≥ 50 µV in at least three of five consecutive trials (43). SI_1mV_ was defined as the stimulus intensity needed to produce MEPs with a mean peak-to-peak amplitude of ∼1 mV. All measurements were taken with the target muscle at rest. The hotspot, RMT, and SI_1mV_ were determined in each testing session.

#### Intracortical Inhibition

Short-interval intracortical inhibition (SICI) and long-interval intracortical inhibition (LICI) were evaluated using single- and paired-pulse TMS protocols. For single-pulse trials, a test stimulus (TS) was delivered at SI_1mV_. For SICI trials, a conditioning stimulus (CS) at 80% RMT preceded the TS at SI_1mV_ by a 3 ms inter-stimulus interval (ISI) (46). For LICI trials, the CS and TS were both delivered at SI_1mV_, separated by a 100 ms ISI (47). Trial conditions (SI_1mV_-alone, SICI, and LICI) were pseudo-randomised using Signal software (CED, UK), with a 5-second inter-trial interval (±20%). Each block consisted of 60 trials: 20 SI_1mV_-alone, 20 SICI, and 20 LICI. Each block lasted approximately 6 minutes. Participants were asked to stay quiet, keep their eyes open, and remain alert throughout the TMS testing.

#### Paired Associative Stimulation (PAS)

PAS was chosen to examine long-term potentiation (LTP)-like neuroplasticity through spike-timing-dependent mechanisms (48). We focused on neuroplasticity driven by sensorimotor integration in individuals with burns, as sensory dysfunction can impair motor function and long-term outcomes (49, 50). PAS was administrated by pairing electrical stimulation of the median nerve on the most-affected side with single-pulse TMS applied to the contralateral M1 (51). Peripheral nerve stimulation was delivered using 200 µs square-wave pulses via a direct current stimulator (Digitimer DS7, UK) and bipolar surface electrodes: the anode was positioned over the median nerve at the wrist, and the cathode 3 cm proximally. For three participants with wrist burns, the peripheral stimulating electrodes were placed on the palmar surface of the third digit on the most affected side, rather than over the median nerve at the wrist. This electrode placement stimulated the median nerve via the palmar digital branches (52). The sensory perceptual threshold was determined through a stepwise procedure involving two reversals. The intensity of peripheral nerve stimulation for PAS was set to three times the sensory perceptual threshold. Single TMS pulses were applied to the contralateral M1 using a Magstim 200^2^ stimulator at SI_1mV_, 25 ms after nerve stimulation. The PAS protocol consisted of 225 paired trials delivered at a frequency of 0.25 Hz over 15 minutes (48). To direct participant attention during PAS, a light-emitting diode (LED) was placed near the target hand and flashed randomly in 20–40 trials throughout the protocol, as controlled by Signal software. Participants were instructed to focus on their hand during PAS and report the number of LED flashes after PAS finished (53). The mean absolute error between reported and true counts was 1.9, with a standard deviation of 2.8. No participants were excluded from the analysis due to errors in LED counting.

Single-pulse MEP amplitude, SICI, and LICI were measured at four time points: before PAS (PRE), immediately after PAS (0 min POST-PAS), 15 minutes post-PAS (15 min POST-PAS), and 30 minutes post-PAS (30 min POST-PAS) (see Figure 1). Participants also rated their self-perceived alertness/sleepiness using the 10-step Karolinska Sleepiness scale after each testing block, including immediately after PAS (54).

### Motor Function: Purdue Pegboard

Motor performance was assessed using the Purdue Pegboard (Lafayette Instrument, USA), which consists of four subtests: unilateral dominant hand, unilateral non-dominant hand, simple bilateral, and bilateral assembly (55). Unilateral subtests were defined as dominant and non-dominant (rather than most-affected and least-affected) to avoid subgrouping complexity and loss of power, given the strong influence of hand dominance on outcomes (56). Accordingly, bilateral pegboard performance was emphasised in the main manuscript to ensure that deficits on the most-affected side were reflected in the overall outcome. Participants were instructed to perform each task as quickly as possible. In the unilateral subtests, participants picked up and placed small pegs into vertically arranged holes on the pegboard in front of them, with the number of pegs placed within 30 seconds recorded for each hand. The testing order for the unilateral subtests was randomised. In the simple bilateral subtest, participants used both hands simultaneously to place pegs in pairs of holes, with the total number of completed pairs scored within 30 seconds. For the assembly subtest, participants alternated hands to assemble 4-item units (peg, washer, sleeve, washer) into the pegboard holes. The number of items in the completed assemblies within 60 seconds was recorded.

### Surveys

All participants were asked to complete the surveys remotely via an email link (REDCap Electronic Database, USA) and completed within 24 hours either side of each session.

#### Short-form 36 (SF36)

Eight domain scores were calculated from the SF36 survey (57–59): physical functioning (assesses the ability to perform daily physical activities, such as walking, climbing stairs, and lifting objects), role limitations due to physical health (measures the extent to which physical health problems interfere with work or other daily activities), role limitations due to emotional problems (measures the extent to which emotional problems affect work or other daily activities), energy/fatigue (captures energy levels and feelings of fatigue), emotional well-being (evaluates psychological well-being, including symptoms of depression, anxiety, and overall mood), social functioning (assesses the impact of physical and emotional health on social activities and interactions), pain (evaluates the intensity of pain and its impact on daily functioning), and general health (reflects overall self-perception of health, including current health status and future health expectations). These domain scores were used to calculate the Physical and Mental Component Summary scores, as described in the Data Processing section.

The SF36 assessment is reported in the main manuscript; the Burn-specific Health Scale-Brief (BSHS-B), Quick Disabilities of the Arm, Shoulder, and Hand (QuickDASH), Lower Limb Functional Index (LLFI), Patient and Observer Scar Assessment Scale (POSAS), and painDETECT assessments are reported in the Supplementary Materials (Sections *S4.1 – S4.5*).

### Perceptual Pain, Sensory Function, and Grip Strength Analysis

Testing of motor and sensory function was conducted in two blocks. Tests that were unlikely to generate a painful response were assessed in the first block (visual analogue pain scale, Semmes-Weinstein Monofilament Test, brush touch perception, and Purdue Pegboard; administered in random order). Tests that might generate a painful response were assessed in the second block (Pain Pressure Threshold, Neuropen, and grip strength; administered in random order). Pegboard performance results are presented in the main manuscript, while the remaining functional assessments are detailed in the Supplementary Materials (Sections S4.*6 – S4.11*).

### Data Processing

All single- and paired-pulse TMS trials were recorded using Signal software (CED, UK) for data acquisition and analysis. Peak-to-peak MEP amplitude 10–60 ms following the stimulus were calculated (and 110–160 ms for LICI). The root mean square (RMS) of background EMG activity was assessed 100 ms preceding the stimulus. Trials were excluded from analysis if the background EMG activity exceeded the RMS mean plus two standard deviations (RMS + 2SD) of all trials in the session (60). The median trial removal rate was 2.9%, with an interquartile range (25th–75th percentile) of 1.3% to 4.9% across all sessions.

For self-reported health and well-being, domain scores from the SF36 were standardised using means and standard deviations from normative population data for gender and age (58, 61). Z-scores for each of the eight subscales were calculated and weighted according to published factor scoring coefficients to derive the Physical Component Summary and Mental Component Summary scores. Composite scores were then transformed to have a mean of 50 and a standard deviation of 10, consistent with conventional scoring methods.

### Data Analysis

Statistical analyses were conducted in R (version 4.4.3) (62). Primary data and code are available at OSF (view-only link:

https://osf.io/n3euw/overview?view_only=a67c490f40864baea2dfce1ba1426490) and in Supplementary Materials (Section *S6*).

#### TMS Stimulator Intensities

To examine whether TMS intensities changed over time, stimulator output required to elicit a 1 mV MEP (SI_1mV_) and resting motor threshold (RMT) were modelled using linear mixed-effects models with a natural cubic spline for days since injury (DAYS) (df = 3) and participant-level random intercepts (63). Spline models were preferred over linear models due to lower Akaike Information Criterion (AIC) values on comparison. Models with 3 degrees of freedom were selected to allow for flexible yet interpretable estimation of non-linear trajectories across time. Model assumptions, including normality, homoscedasticity, and linearity, were assessed using residual plots and the performance diagnostic package (64), all of which indicated adequate model fit. Marginal means (intensities) and trends (slopes) were computed at 23, 44, and 82 days using emmeans and emtrends (65), respectively, noting these align closely with 3, 6, and 12 weeks post-injury—clinically meaningful milestones and the densest scheduled assessment periods. Holm-adjusted comparisons were then used to evaluate time-dependent changes in stimulator intensity.

#### Modelling of Intracortical Inhibition and Neuroplasticity Over Time

The current analysis approach was designed to evaluate changes in intracortical inhibition over time and to better understand motor cortex neuroplasticity during subacute burn recovery. Generalised linear mixed models (GLMMs) were fitted using the lme4 package (63), specifying a Gamma distribution with a log link to model trial-level MEP amplitudes. The models included fixed effects for days since injury (DAYS, modelled using a natural cubic spline with 3 degrees of freedom), stimulation condition (CONDITION: single-pulse, SICI, LICI), and PAS block (BLOCK: PRE, 0-, 15-, and 30-MIN POST-PAS), including all interactions. Participant-specific intercepts and slopes for DAYS and CONDITION were included as random effects. Despite a minor convergence warning (max|grad| = 0.0176), results were consistent across optimisers, and the bobyqa-based model was retained (63). Model fit was evaluated using the AIC and likelihood ratio tests. Residual plots stratified by CONDITION revealed mild heteroscedasticity and right-skew, particularly for single-pulse and LICI trials. However, no substantial violations of linearity or distributional assumptions were observed, and the residual patterns were consistent with the Gamma distribution specified in the model. Estimated marginal means (amplitudes) and marginal trends (slopes) were extracted at 23, 44, and 82 days post-injury using the emmeans and emtrends packages (65). Instantaneous slopes, representing the first derivative (tangent) of the fitted curve, were used to assess the rate of change in MEP amplitude over time. Model-based contrasts estimated differences in marginal means and trends between SICI and LICI versus single-pulse trials, expressed on the response scale as ratios (marginal-mean ratios and slope ratios) interpretable as relative inhibition (<1) or facilitation (>1). Post hoc contrasts were performed within the PRE block and across PAS blocks with Holm correction. Percent changes for contrasts were also reported.

#### Analysis of Pegboard Performance Over Time

To model changes in pegboard performance over time, separate linear mixed-effects models were fit for each subtest (dominant, non-dominant, simple bilateral, assembly), with DAYS modelled as a natural cubic spline (df = 3) and random intercepts for participants (63). Spline models were preferred over linear alternatives due again to lower AIC values across all subtests, and residual diagnostics confirmed appropriate model fits (64). Both estimated marginal means (scores) and marginal trends (slopes) were extracted at 23, 44, and 82 days post-injury to capture changes in performance levels and the rate of change over time (65). Slopes derived from the first derivative of the fitted spline were used to quantify the instantaneous rate of performance change across time. Holm-adjusted pairwise comparisons were used to evaluate differences across timepoints for both approaches. For the score-based analyses, percent change was calculated by dividing the estimated difference in marginal means by the earlier timepoint value, multiplied by 100. For the slope-based analyses, differences were based on estimated marginal trends extracted at 23, 44, and 82 days.

#### Changes in SF36 Physical and Mental Component Summary Scores Over Time

The Physical Component Summary and Mental Component Summary scores were calculated by adjusting SF36 subscale scores against age and gender-based normative means and standard deviations from healthy individuals (58, 61). These standardised, weighted scores reflect participants’ health relative to expected values in a healthy population, enabling direct comparison of physical and mental functioning. To assess changes over time since injury, separate linear mixed-effects models incorporating natural cubic splines (df = 3) for days post-injury were fitted for Physical and Mental Component Summary scores using the lme4 package (63), including participant-specific random intercepts. Spline models were preferred over linear alternatives with lower AIC values for both outcomes. Residual diagnostics confirmed appropriate model fits (64), with no substantial violations of linearity, normality, or homoscedasticity. Mild right-skew and heteroscedasticity were observed for Mental Component Summary scores, but the deviations were within acceptable bounds for linear mixed-effects modelling (66). These findings supported the use of spline-based models to capture non-linear recovery trajectories for both physical and mental health. Estimated marginal means (scores) and marginal trends (slopes) at days 23, 44, and 82 were obtained using emmeans and emtrends (65), and pairwise comparisons between these timepoints were adjusted for multiple testing using the Holm method. For the score-based analyses, percent change was calculated by dividing the estimated difference by the earlier timepoint value and multiplying by 100. For the slope-based analyses, differences in the estimated daily rate of change were calculated between timepoints using model-derived estimates.

#### Time-Varying Associations Between Pegboard Performance and Baseline and Neuroplastic TMS Outcomes and SF36 Summary Scores

To assess whether baseline intracortical inhibition could predict pegboard performance over time, separate models were fit for aggregated SICI and LICI amplitude ratios, each including an interaction between DAYS and simple bilateral or assembly pegboard subtests (63). This interaction tested whether the strength of the pegboard–inhibition association varied across the recovery period. Analyses used bilateral subtests to align the hemisphere-specific neurophysiological measure—assessed in the hemisphere contralateral to the most-affected hand—with pegboard performance; because bilateral scores aggregate both hands, the most-affected hand’s deficit is embedded in the pegboard score and thus informs both measures. Assumptions of linearity and homoscedasticity were met for all four models (64). While residuals deviated from normality, this is not considered a major concern, as linear mixed-effects models are generally robust to such violations (66). No influential outliers were detected. Overall, the models appeared suitable for interpretation. Slopes of inhibition predicted by pegboard scores were extracted using the emtrends package at 23, 44, and 82 days (65), with day-specific significance determined when the 95% confidence interval (CI) did not cross zero, and between-day comparisons assessed using Tukey-adjusted *p*-values.

The same analytical approach was also used to examine whether changes in single-pulse MEP amplitude following PAS (calculated as the difference between pre- and post-stimulation blocks, with all post-stimulation trials collapsed into a single block) could predict bilateral pegboard performance over time. Assumption checks confirmed that both the simple bilateral and assembly models demonstrated acceptable linearity and homoscedasticity, with residuals approximately normally distributed and no influential outliers identified (64). These results support the robustness and interpretability of the fitted models.

An exploratory analysis was conducted to examine whether the association between PAS-induced changes in LICI and pegboard performance varied across the recovery period. PAS-induced change was calculated as the difference between baseline (PRE) and 30-minute POST-PAS blocks, based on visual inspection indicating that the largest LICI responses were evident at this timepoint. Separate linear mixed-effects models were fitted for the simple bilateral and assembly subtests, including an interaction between pegboard score and days since injury, with participant-level random intercepts (63). Visual inspection of diagnostic plots indicated approximate linearity and no influential outliers for both models (64). Mild heteroscedasticity and deviations from normality were observed; however, given the exploratory nature of the analysis and the robustness of linear mixed-effects models to such violations (66), these issues were not considered likely to meaningfully compromise model interpretation. Estimated marginal trends were extracted at 23, 44, and 82 days post-injury to assess the time-varying association between pegboard performance and LICI response (65). Pairwise comparisons of these slopes were used to evaluate changes in the strength of association across time.

For all analyses, statistical significance was defined as *p* < .05. Data preprocessing and visualisation were performed using R packages: dplyr (67), tidyverse (68), ggplot2 (69), and patchwork (70).

## Results

### Characteristics of Subacute Burn Patients

Thirty-four subacute burn patients (16 females; age: 46.0 ± 14.4 years) participated in this longitudinal study. The average total body surface area (TBSA) affected was 1.0 ± 2.0%. Burn causes included contact (n = 7), scald (n=14), flame (n = 9), chemical (n = 3), and friction (n = 1). Burn depths were classified as superficial (n = 12), partial-thickness (n = 14), deep (n = 8), and full-thickness (n = 0). Two-thirds of injuries (n = 20) involved the upper limb—hand, forearm, or upper arm.

### Pegboard Performance

#### Score Differences

Pegboard performance was modelled over the observed range (4–124 days), with margins estimated and contrasts evaluated at 23, 44, and 82 days post-injury for each subtest. Figure 2 (top panel) shows estimated trajectories of bilateral pegboard performance during recovery. For the assembly subtest (B), performance improved significantly from day 23 to 44 (+5.0%, adjusted *p* = .034), but no further changes were observed between day 44 and 82 (−0.8%, adjusted *p* = .782), and the overall gain from day 23 to 82 was not significant (+4.2%, adjusted *p* = .328). In contrast, simple bilateral scores (A) showed no significant improvement from day 23 to 44 (+1.6%, adjusted *p* = .319) but declined significantly from day 44 to 82 (−6.6%, adjusted *p* = .019).

**Figure 2.**
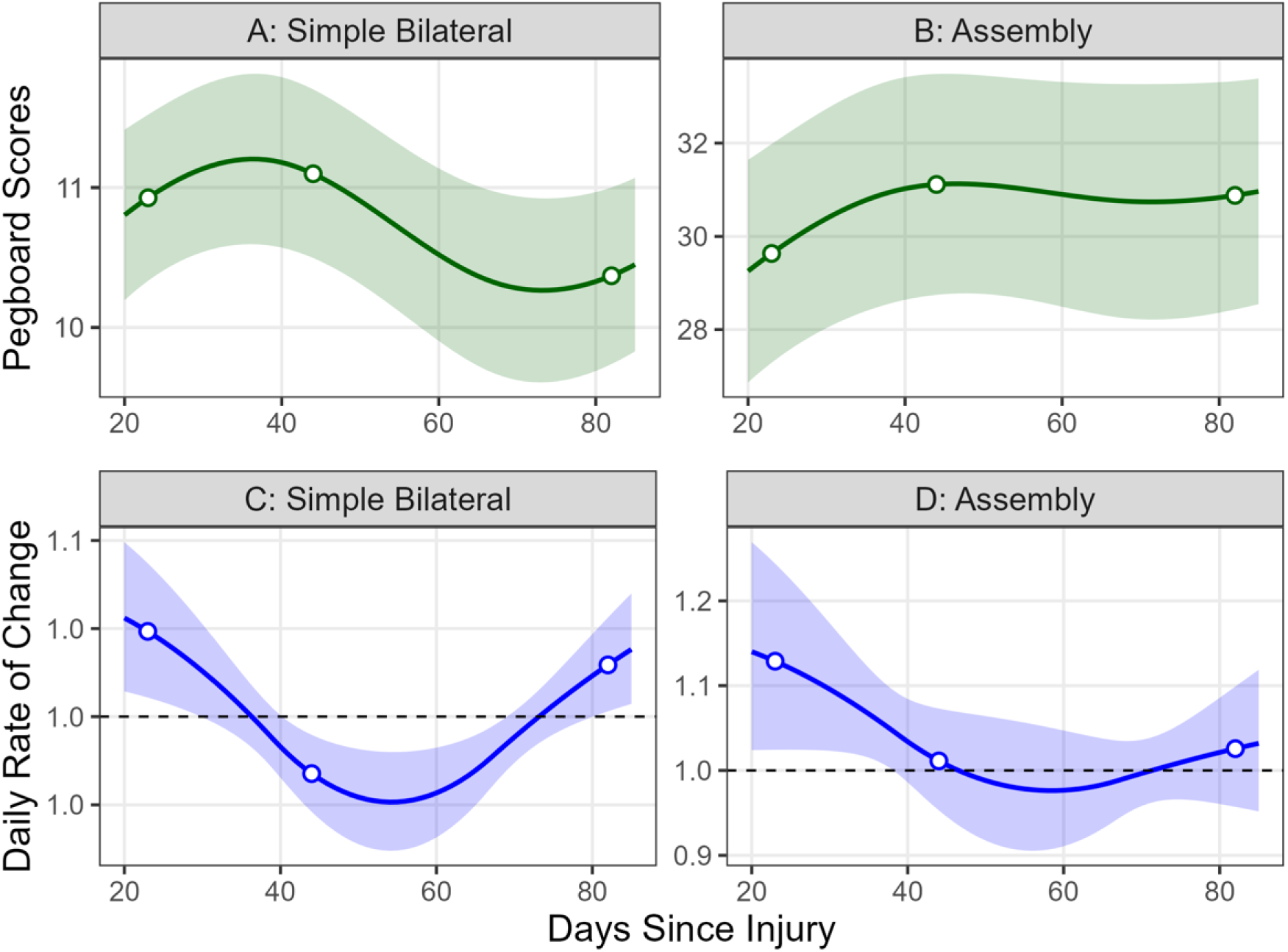
Panels A and B show modelled trajectories of score-level differences in pegboard task performance (A: simple bilateral; B: assembly) over time following injury. Shaded bands in (A) and (B) indicate 95% confidence intervals. Panels C and D show the daily rate of change in pegboard task performance (C: simple bilateral; D: assembly) following injury. Curves show performance improvements or slowing over time: a value of 1.0 (dashed line) indicates no change; values >1.0 reflect improvement in pegboard performance over time; values <1.0 reflect reduced performance over time. Shaded bands in (C) and (D) represent uncertainty in the estimates. Open circles represent the estimates at 23, 44, and 82 days post-injury.

For the dominant hand, scores improved significantly over time, with a +4.9% increase from day 23 to 44 (adjusted *p* = .012) and a +8.7% increase from day 23 to 82 (adjusted *p* = .006). There was no significant change between days 44 and 82 (+3.7%, adjusted *p* = .142). Similarly, non-dominant hand performance improved, showing a +4.4% increase from day 23 to 44 (adjusted *p* = .018) and a +5.7% increase from day 23 to 82 (adjusted *p* = .044), with no further change between days 44 and 82 (+1.2%, adjusted *p* = .583). Unilateral performance is shown in Supplementary Materials (Section *S1*, *Figure S1*).

#### Rate of Change Differences

Pegboard slope analysis revealed task-specific differences in recovery trajectories over time. Figure 2 (bottom panel) shows the modelled day-by-day change in bilateral pegboard task performance following injury. No significant differences in the daily rate of change were detected for the assembly task (all adjusted *p* ≥ .278). The simple bilateral subtest’s daily improvement rate significantly declined from days 23 to 44 (−0.06 units/day, adjusted *p* = .005), indicating slower gains, followed by a significant increase in the rate of improvement from days 44 to 82 (+0.05 units/day, adjusted *p* = .017). These results suggest a transient slowdown in bilateral performance gains around 6 weeks post-injury, which resolved by 82 days post-injury.

For the unilateral hand subtests, there were no significant changes in the daily rate of improvement between any timepoints (all adjusted *p* ≥ .447), indicating relatively stable recovery trajectories. Unilateral performance is shown in Supplementary Figure S1.

### Short-form 36 (SF36) Summary Scores Across Recovery Period

#### Score Differences

Figure 3 (top panel) shows modelled trajectories of Short-form-36 (SF36) Physical Component Summary and Mental Component Summary scores across days since injury. Marginal estimates were extracted and contrasts evaluated at 23, 44, and 82 days post-injury from models fit over the observed range (4–124 days). For the Physical Component Summary, a significant 8.1% increase was observed from day 23 to day 44 (adjusted *p* < .001), which was unchanged at day 82 with a similar 8.2% increase compared to day 23 (adjusted *p* = .009). No significant change occurred between days 44 and 82 (adjusted *p* = .963). For the Mental Component Summary, the 3.7% increase from day 23 to day 44 was not significant (adjusted *p* = .121), while the 6.7% increase from day 23 to day 82 approached significance (adjusted *p* = .070). No significant difference was observed between days 44 and 82 (adjusted *p* = .283). These results suggest early improvements in physical health-related quality of life during the first six weeks post-injury, with Physical Component Summary scores stabilising and Mental Component Summary scores continuing to trend upward by 12 weeks. Physical Component Summary values remained above the population norm of 50, suggesting better-than-average perceived physical health relative to healthy individuals, whereas Mental Component Summary values were more closely aligned with population norms.

**Figure 3.**
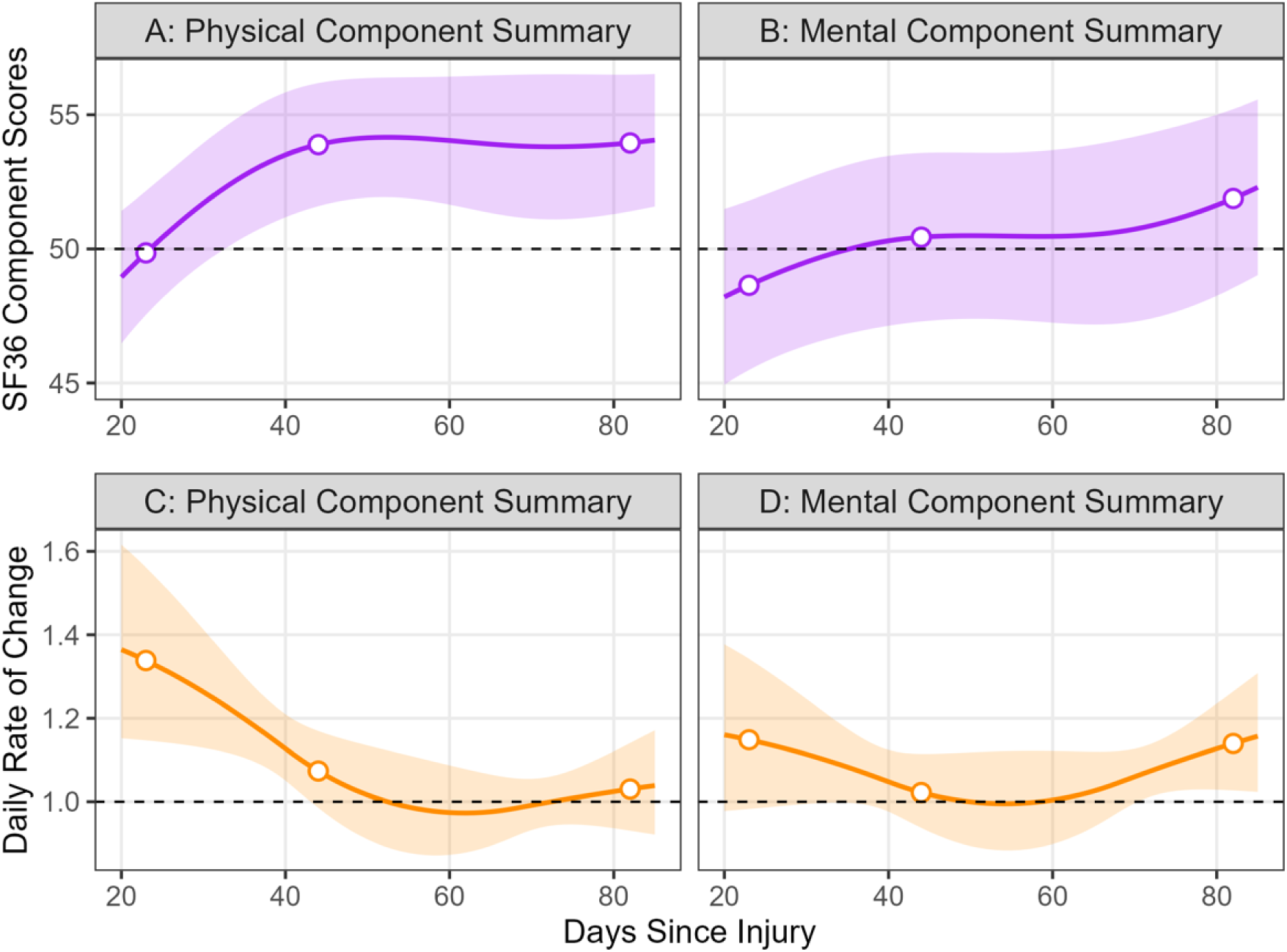
Panels A and B show estimated marginal means of SF36 Physical and Mental Component Summary scores over time post-injury. Scores are standardised to a population mean of 50 (SD = 10); the dashed line marks this reference. Panels C and D show estimated relative daily rates of change in Physical and Mental Component Summary scores over time. The dashed line at 1.0 indicates no change; values above 1.0 reflect improvement, and values below 1.0 indicate reduced recovery. For all panels, shaded bands represent 95% confidence intervals. Open circles show the model-predicted values at days 23, 44, and 82.

#### Rate of Change Differences

Figure 3 (bottom panel) shows instantaneous slopes in SF36 Physical and Mental Component Summary scores at days 23, 44, and 82 post-injury. For the Physical Component Summary, the rate of improvement declined from day 23 to 44 (–0.22 units/day, adjusted *p* = .052), approaching statistical significance, and was significantly lower by day 82 compared to day 23 (–0.26 units/day, adjusted *p* = .013). No significant difference was observed between days 44 and 82 (–0.04 units/day, adjusted *p* = .641), indicating a plateau in physical recovery rate beyond ∼6 weeks post-injury. For the Mental Component Summary, slope contrasts were not statistically significant across timepoints (e.g., 23 to 82: –0.01 units/day, adjusted *p* = .929), although the slope remained positive at day 82

### Neurophysiological Measures

#### Baseline TMS Intensities (RMT, SI_1mV_)

##### Baseline Intensity Differences

Resting motor threshold (RMT) and SI_1mV_ (stimulator output required to elicit a 1 mV MEP) were modelled as a function of days since injury. Margins were estimated from models fit over the observed range (4–124 days) and extracted—with contrasts evaluated—at 23, 44, and 82 days post-injury. SI_1mV_ remained stable over time, with percent changes ranging from −1.4% to +2.0%. No differences reached statistical significance after Holm correction (all adjusted *p* ≥ .080). RMT values showed similar stability, with no significant differences observed between any timepoints (all adjusted *p* = 1.000). Data are presented in Supplementary Materials (Section *S2*, *Figure S2*).

##### Rate of Change Differences

Pairwise contrasts of slope estimates showed no statistically significant changes in corticospinal excitability (SI_1mV_) across timepoints (adjusted *p* ≥ .102). No contrasts for RMT were significant (all adjusted *p* = 1.000), indicating no evidence of time-dependent changes in RMT across recovery. Data are presented in Supplementary Materials (Section *S2*, *Figure S2*).

#### SICI and LICI in Subacute Burn Patients

##### Baseline Intracortical Inhibition Changes

Figure 4 (top panel) show modelled trajectories of single-pulse MEP amplitude and intracortical inhibition (SICI and LICI) amplitude ratios over time during the baseline (PRE) block. We evaluated marginal mean MEP amplitudes, SICI, and LICI ratios during the baseline (PRE) block at 23, 44, and 82 days post-injury. At all three time points, both SICI and LICI conditions showed inhibition relative to single-pulse trials. Single-pulse trial amplitude estimates were close to 1.0 at each time point: pairwise comparisons indicated no significant differences across time (all adjusted *p* = 1.000). Single-pulse MEP amplitude remained stable across the subacute period and provided a stable reference for evaluating SICI and LICI.

**Figure 4.**
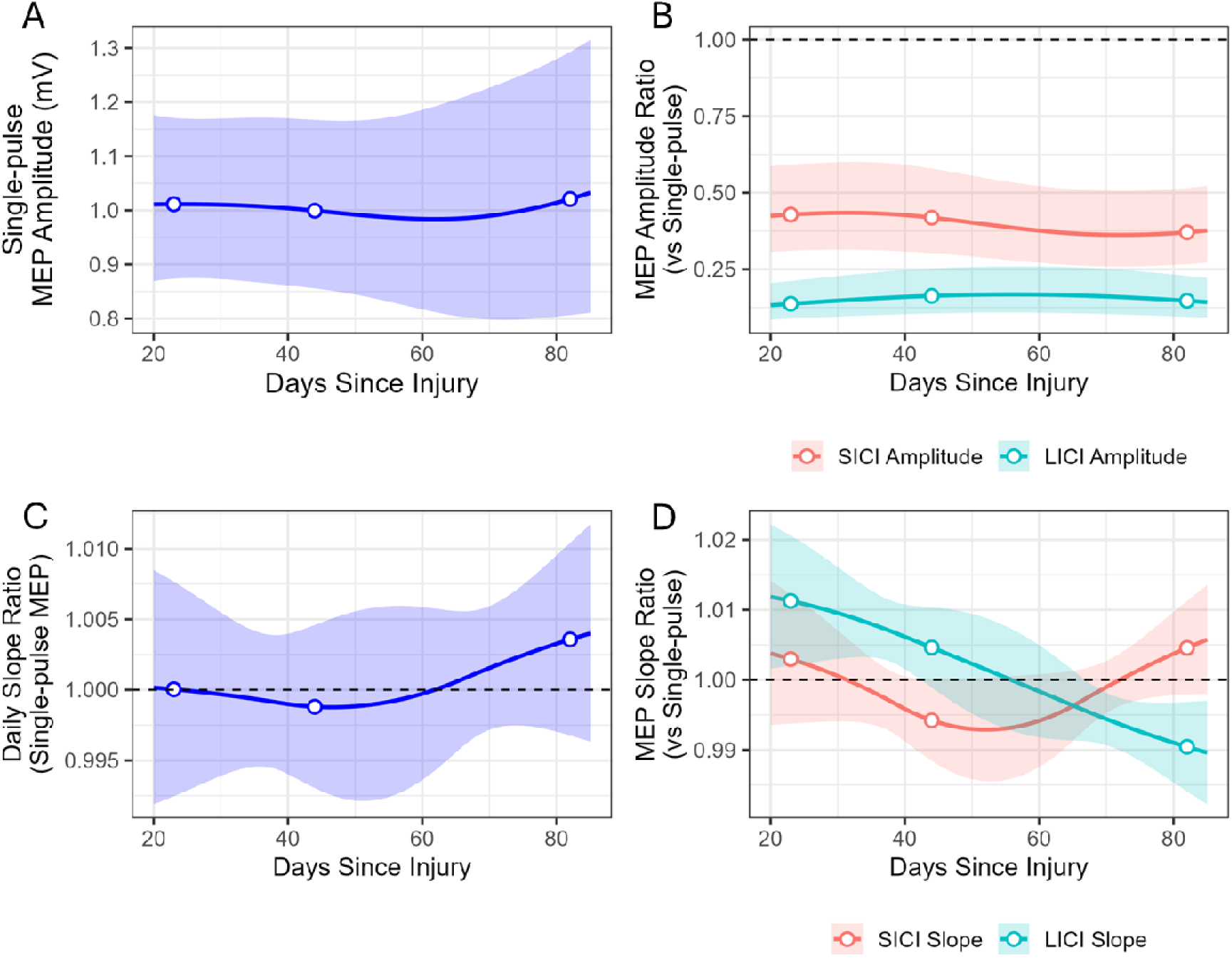
Panels A and B show modelled changes in single-pulse MEP amplitude (A) and SICI and LICI amplitude ratios (B) across days since injury during the baseline (PRE) block. SICI and LICI amplitude ratios (relative to single-pulse) <1 indicate inhibition compared to single-pulse trials, with smaller ratios indicating greater inhibition. Panels C and D show modelled changes in daily MEP slope ratios (A) and SICI and LICI slope ratios (B) across days since injury during the baseline (PRE) block. Values greater than 1.0 indicate an increase in single-pulse MEP amplitude over time, whereas values less than 1.0 reflect a decrease across days. For SICI and LICI slope ratios, values above 1.0 indicate a relative reduction in inhibition over time, while values below 1.0 reflect increasing inhibition across days. For all panels, shaded bands represent 95% confidence intervals and open circles show modelled points correspond to key analysis days (23, 44, 82).

Both SICI and LICI amplitude ratios remained stable across time. Pairwise comparisons showed no significant differences in amplitude for either SICI or LICI ratios across days (all adjusted *p* = 1.000).

##### Rate of Change Differences

Figure 4 (bottom panel) shows the day-by-day change in MEP slope ratios during the baseline (PRE) block following injury. PRE-block marginal slopes were estimated from models fit over the observed range (4–124 days) and evaluated at 23, 44, and 82 days post-injury. Slope estimates for single-pulse trials indicated stable corticospinal excitability across the subacute period. No significant differences were found between days (all adjusted *p* ≥ .887)

No significant changes were detected in the SICI slope ratio (relative to single-pulse trials); there was a modest increase from days 44 to 82 (+0.010 units/day, adjusted *p* = .065) that did not reach significance. For LICI, slope ratios decreased significantly from day 23 to 82 (−0.021 units/day, adjusted *p* < .001) and from day 44 to 82 (−0.014 units/day, adjusted *p* = .002), indicating a progressive increase in LICI over the subacute period. Values above 1.0 with a negative slope indicate that LICI is increasing over time (i.e., reduced inhibition is reversing), while values below 1.0 with a negative slope reflect further increases in LICI, indicating progressively stronger inhibition.

#### Single-pulse MEP Amplitude and Intracortical Inhibition Following PAS

##### Amplitude Differences

Figure 5 (left panel) shows the day-by-day changes in corticospinal excitability and intracortical inhibition following paired associative stimulation (PAS), shown relative to the PRE block. Marginal mean MEP amplitudes were estimated from models fit over the observed range (4–124 days) and evaluated across post-PAS blocks (0-, 15-, and 30-min POST-PAS) at 23, 44, and 82 days post-injury. Single-pulse amplitudes showed small, non-significant changes across time (all adjusted *p* ≥ .916), indicating that corticospinal excitability following PAS remained relatively stable over the subacute recovery period.

**Figure 5.**
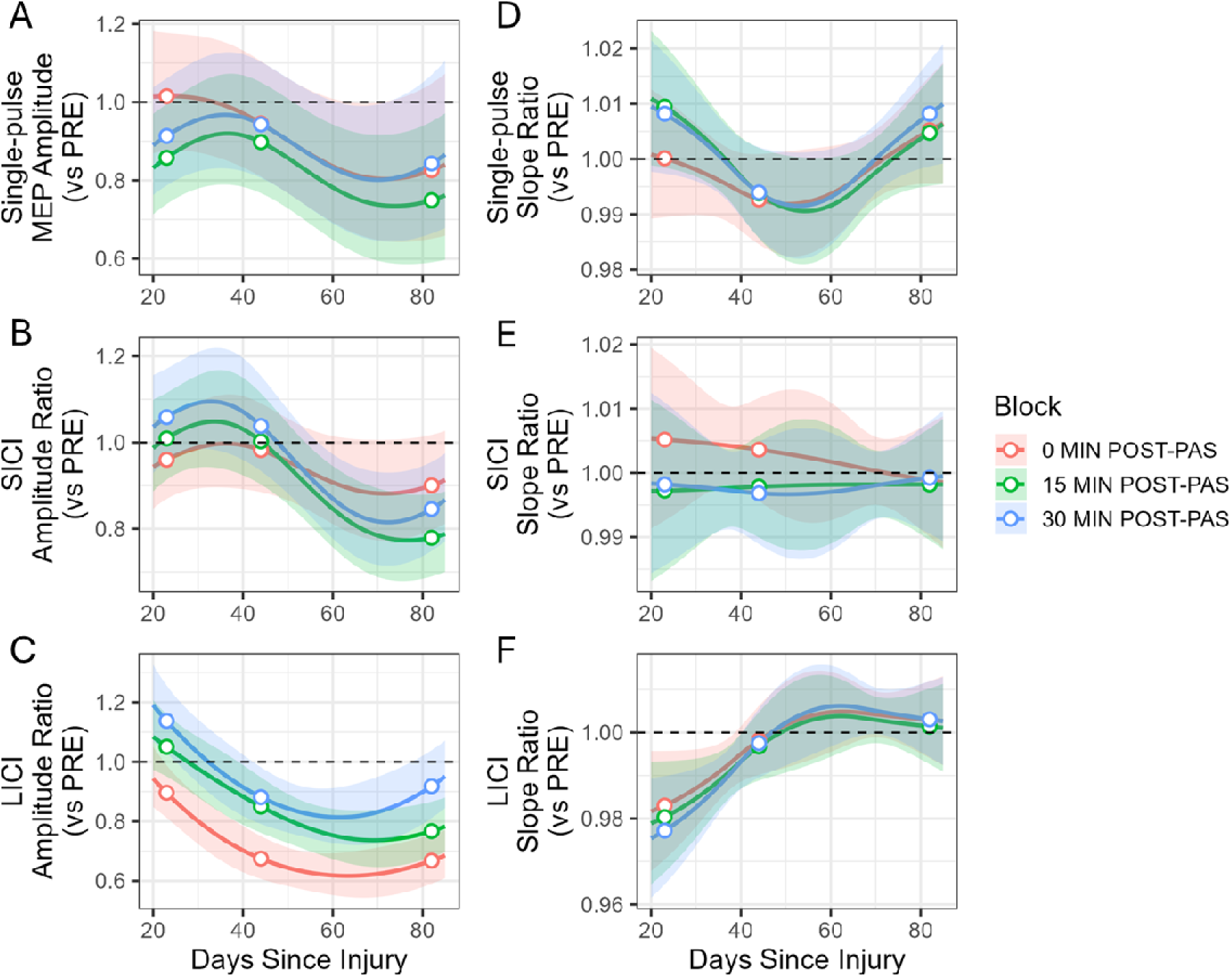
Left panel shows modelled changes in MEP amplitude (A), SICI (B), and LICI (C) following paired associative stimulation (PAS), expressed relative to the PRE block (dashed line at ratio = 1.0). Ratios below 1.0 reflect increased inhibition relative to the PRE block. Block conditions are color-coded: 0 min POST-PAS (red), 15 min POST-PAS (green), and 30 min POST-PAS (blue). Right panel shows model-estimated slope ratios (relative to the PRE block) for single-pulse (D), SICI (E), and LICI (F) conditions following paired associative stimulation (PAS) at 0, 15, and 30 min POST-PAS. Ratios <1.0 indicate reduced daily MEP slope compared to PRE. For all panels, shaded bands represent 95% confidence intervals, and open circles indicate values at key assessment days (23, 44, and 82 days post-injury).

SICI amplitude ratios showed no significant changes across any post-PAS block or timepoint (all adjusted *p* ≥ .606). In contrast, LICI amplitude ratios showed a significant increase in inhibition from day 23 to 44 at 15 min POST-PAS (−22.7%, adjusted *p* = .044) and 30 MIN POST-PAS (−24.9%, adjusted *p* = .020). No other LICI comparisons reached significance (all adjusted *p* ≥ .121), suggesting that the effect was transient.

##### Rate of Change Differences

Figure 5 (right panel) shows the day-by-day changes in corticospinal excitability and intracortical inhibition slope ratios following PAS, shown relative to the PRE block. We examined slope estimates for single-pulse trials, SICI, and LICI across the post-PAS time period. Single-pulse trial slope comparisons showed modest changes over time, with a trend toward reduced excitability from day 23 to 44 at 15 min POST-PAS (−0.016 units/day, adjusted *p* = .077) and 30 min POST-PAS (−0.014 units/day, adjusted *p* = .079), followed by a partial return from day 44 to 82 at 30 min POST-PAS (+0.014 units/day, adjusted *p* = .079). Overall, the rate of change in corticospinal excitability remained stable throughout the subacute period following PAS.

SICI slope contrasts were non-significant across all pairwise comparisons (all adjusted *p* = 1.000). LICI slope ratios progressively increased, consistent with a time-dependent reduction in the PAS-induced increase in LICI. Significant increases in LICI slope values from day 23 to 82 were observed across all post-PAS blocks: at 0 min POST-PAS (+0.020 units/day, adjusted *p* = .036), 15 min POST-PAS (+0.021 units/day, adjusted *p* = .026), and 30 min POST-PAS (+0.026 units/day, adjusted *p* = .003). An additional significant increase was also observed from day 23 to 44 at 30 min POST-PAS (+0.021 units/day, adjusted *p* = .016). Other comparisons did not reach significance following correction (e.g., 15 min POST-PAS: 23 to 44, +0.017 units/day, adjusted *p* = .069).

In summary, amplitude-ratio results show that PAS increased LICI within-session, with the largest increase around day 44. However, rate of change (slope) results across days show that the magnitude of this PAS-related increase decreased from day 23 to day 82. Taken together, PAS-induced LICI peaks near day 44 and then decreases over time.

#### Baseline SICI and LICI and Bilateral Pegboard Performance

Figure 6 shows the estimated associations between baseline intracortical inhibition and bilateral pegboard subtest scores at each timepoint. For SICI amplitude ratios, the association with simple bilateral performance was positive at all timepoints, with the steepest slope observed at day 23 (+0.089 units per unit increase, 95% CI [0.003, 0.175]). Pairwise comparisons of these slopes across days did not reach significance after correction (all adjusted *p* ≥ .390). The association with SICI and assembly performance was weak and nonsignificant at all timepoints, with no significant between-day differences (all adjusted *p* ≥ .949). For LICI amplitude ratios, the association with both simple bilateral performance and assembly performance was weak and not statistically significant at any timepoint (all adjusted *p* ≥ .778).

**Figure 6.**
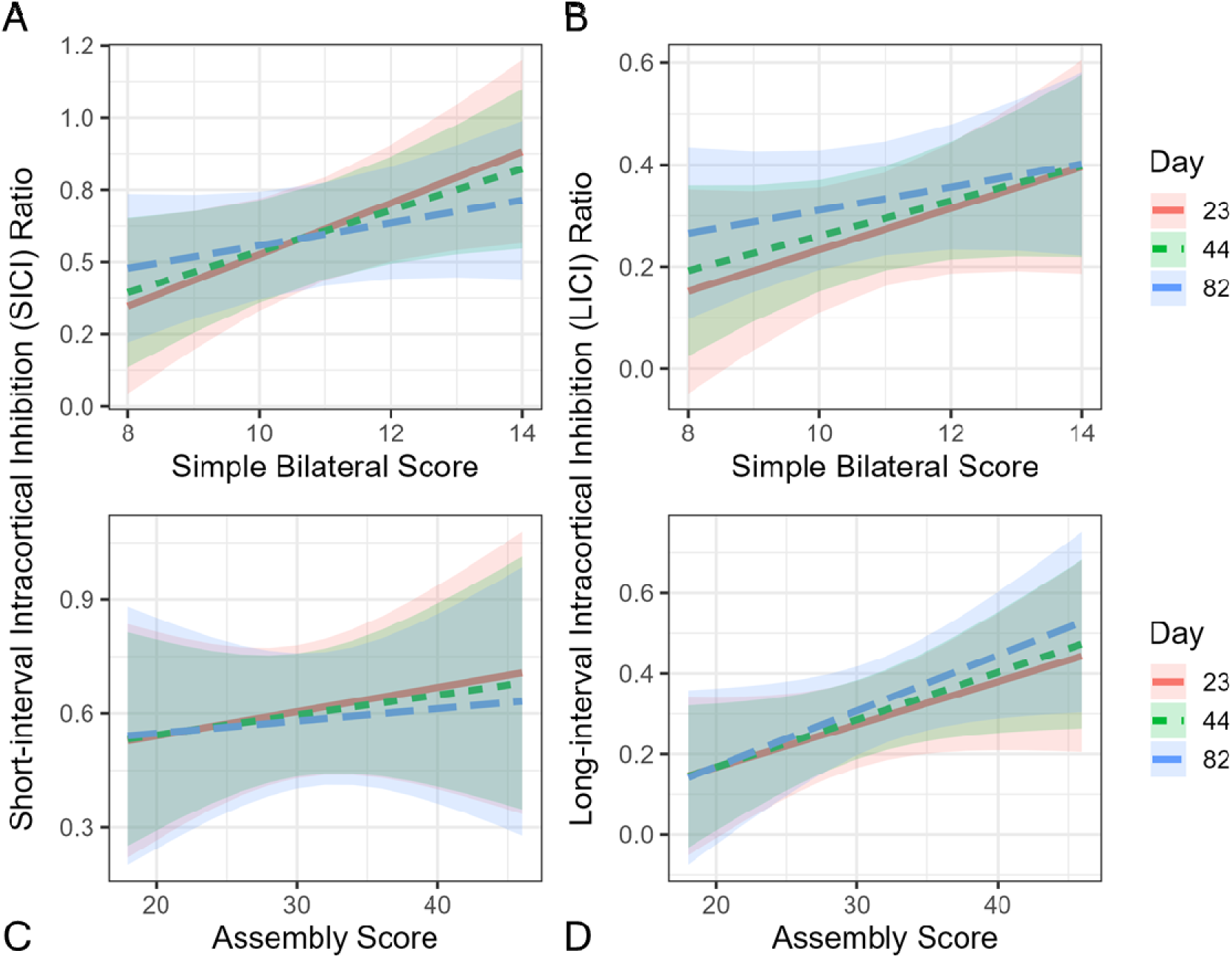
Estimated relationships between bilateral pegboard subtest scores and inhibition amplitude ratios across timepoints. (A–B) display relationships between simple bilateral scores and SICI (A) and LICI (B). (C–D) show relationships between assembly scores and SICI (C) and LICI (D). Lines represent model-estimated slopes, and shaded bands represent 95% confidence intervals.

#### Single-pulse MEP Amplitude Following PAS and Bilateral Pegboard Performance

Linear mixed models were used to examine whether bilateral pegboard task performance was associated with changes in single-pulse MEP amplitude following PAS. For both the simple bilateral subtest and the assembly subtest, no significant associations were observed between task performance and changes in MEP amplitude at any timepoint, and no differences in the strength of association were detected across days (all adjusted *p* ≥ .609). Together, these results suggest that changes in corticospinal excitability following PAS were not related to bilateral pegboard task performance at any of the assessed timepoints. Data are presented in Supplementary Materials (Section *S3*, *Figure S3*).

#### LICI Ratio Change Following PAS and Bilateral Pegboard Performance

Given the significant change in LICI following PAS, and the difference in PAS-induced LICI across days post-injury, day-specific slope estimates were derived to examine how changes in LICI following PAS (PRE – 30 min POST-PAS) related to bilateral pegboard performance at 23, 44, and 82 days post-injury. Figure 7 shows model-estimated associations between changes in LICI following PAS and bilateral pegboard performance at each timepoint. For simple bilateral performance, a significant negative association was observed at day 44, (95% CI: −0.073 to −0.004), indicating greater PAS-induced LICI increase was associated with better simple bilateral subtest performance. Non-significant associations were observed between simple bilateral subtest performance and PAS-induced change in LICI at 23 and 82 days post-injury (23 days: 95% CI: −0.089 to 0.001; 82 days: 95% CI: −0.064 to 0.007). Pairwise contrasts of slopes across days revealed no statistically significant differences (all adjusted *p* ≥ .802), suggesting that the strength of the relationship between LICI and sequenced bilateral performance was unchanged over the recovery period. For the assembly task, no significant associations were detected at any timepoint. Pairwise contrasts across days were likewise non-significant (all adjusted *p* ≥ .576), reflecting stable associations over time.

**Figure 7.**
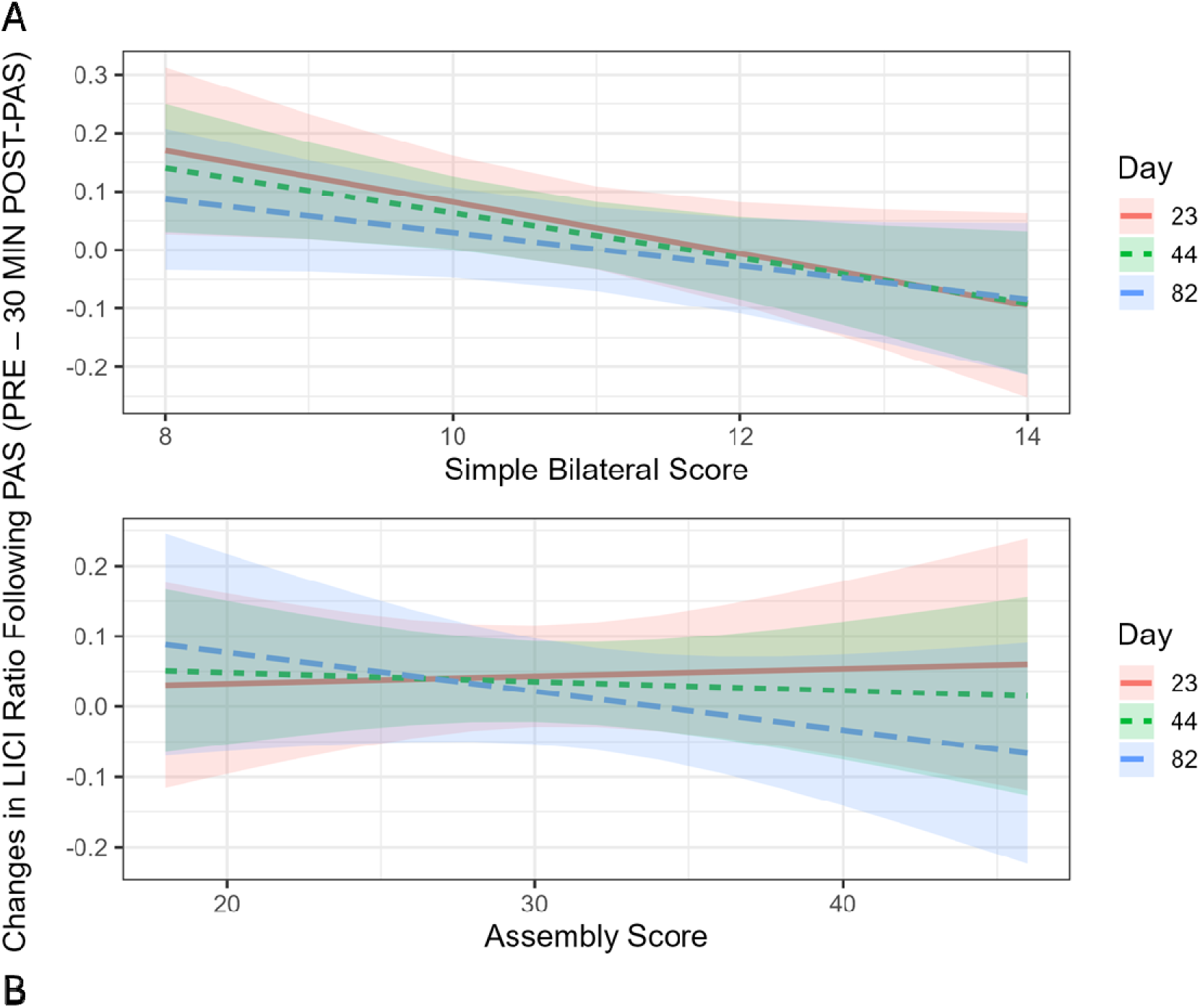
Model-estimated associations between changes in LICI amplitude ratio following PAS (PRE – 30 min POST-PAS) and simple bilateral (A) and assembly (B) pegboard performance across Days 23, 44, and 82 post-injury. Shaded bands represent 95% confidence intervals for each day-specific slope estimate.

#### Sleepiness Scores Following PAS

Participants reported a significant increase in sleepiness following PAS. Compared to PRE, sleepiness was significantly higher at all post-PAS blocks (all adjusted *p* ≤ .049), peaking immediately after PAS and gradually declining over time, with sleepiness in the final post-PAS block significantly lower than immediately after PAS (adjusted *p* = .002).

## Discussion

This study examined motor performance, neuroplasticity, and motor cortex inhibition changes during the first 12 weeks following minor burn injury. There were three key results. First, improvements in motor performance and overall physical and mental functioning were greatest between ∼3 and ∼6 weeks following burn injury and then plateaued to ∼12 weeks following burn injury. Second, baseline resting-state, LICI progressively increased across the first ∼12 weeks following injury, suggesting increasing GABA_B_-mediated inhibition over the studied period. Third, PAS increased LICI at 30 minutes following stimulation and the magnitude of this PAS-induced increase in LICI gradually reduced over the first ∼12 weeks following burn injury, suggesting PAS increased GABA_B_-mediated inhibition with the greatest change early following burn injury. In addition, the PAS-induced increase in LICI was associated with bilateral motor performance, such that a greater PAS-induced increase in LICI was associated with better bilateral motor performance.

### Subacute Functional Recovery Trajectories After Minor Burn Injury

Motor performance showed the greatest rate of improvement early following burn injury. Unilateral performance of both the dominant and non-dominant hand and the complex bilateral assembly task performance increased from ∼3 to ∼6 weeks following injury and then plateaued to ∼12 weeks following injury. The current findings are consistent with previous self-report data, measured by the QuickDASH, showing improvements in upper-limb motor function 1 to 3 months following non-severe burn injuries (3). Together, these findings are consistent with data showing significant positive correlations between self-report and objective measures of hand function ∼18 months following burn injury (71). Our findings extend this by providing objective evidence of motor performance improvements in the subacute period and showing that the greatest improvements occur within ∼6 weeks following injury. Motor performance in the ∼12 weeks following injury was not different to control participants (see Supplementary Materials Section *S5.2* and *Figure S5.2*). This trajectory is similar to previous findings of improvements in motor function in the subacute period for severe burn injuries (72), and consistent with previous research showing no difference in fine motor control, measured using the self-report Burn Outcome Questionnaire, between severe and minor burn injury patients 3 years following injury (73). This may indicate that the early post-burn period presents an important window of opportunity for interventions aimed at optimising the neuroplastic response, and conversely, provide insight into the possible mechanisms supporting rapid reinforcement of maladaptive behaviours early after burn injury.

In this study, simple bilateral performance differed from the unilateral tasks and complex bilateral assembly task, with a significant decline in simple bilateral performance between ∼6 and ∼12 weeks. It is unclear why patients showed a decline in a simple bilateral movement but not a complex bilateral movements, but could perhaps be due to the synchronous nature of the simple bilateral movement: it might reflect difficulties in timing and synchronisation between hands, which depend on interhemispheric connectivity (74, 75). This, however, is speculative and further research is needed to characterise potential changes in interhemispheric connectivity post-injury and examine any associations with bilateral motor performance.

Self-reported physical and mental health scores from the SF36 survey improved most between ∼3 and ∼6 weeks post-injury, particularly for physical health. Physical scores increased significantly during this period and then plateaued, while mental health scores continued to trend upward, albeit more gradually. These patterns suggest that perceived health and well-being recover most rapidly in the early subacute phase following minor burn injury, with physical recovery stabilising earlier than mental recovery—consistent with meta-analytic findings showing the greatest improvements in health-related quality of life during the first ∼6–8 weeks post-injury (76). These findings are also consistent with previous research showing that, compared to severe burns, individuals with minor injuries typically report faster functional recovery, and a quicker return to daily activities (76), and that health-related quality of life scores in individuals with minor burns often remain within normative ranges (1). SF36 scores at ∼12 weeks following injury were not different to control participants (see Supplementary Materials Section *S5.3* and *Figure S5.3*). Interestingly, Physical Component Summary scores from the SF36 remained above the normative population mean of 50 (61, 77), indicating higher perceived health and well-being compared to the general healthy population. This finding might reflect a self-selection bias: previous research has shown individuals participating in health research have high levels of education, employment, and health compared to individuals who do not participate in research (78).

### Increased Baseline LICI But Not SICI ∼3 to ∼12 Weeks Following Burn Injury

Baseline SICI amplitude ratios remained stable across time, suggesting no meaningful changes in the excitability of GABA_A_-mediate inhibition between ∼3 and ∼12 weeks following minor burn injury (24, 30, 79). These findings replicate prior reports of no SICI change from 6–12 weeks (34) and extend them by showing no difference at ∼3 weeks in our cohort. While there was no significant difference in SICI ∼3 to ∼12 weeks following injury, cross-sectional analysis comparing burn patients and non-injured controls showed SICI was greater for controls than burns patients at ∼3 and ∼6 weeks post-injury (adjusted *p* = .048, .045) but not ∼12 weeks post-injury (see Supplementary Materials Section *S5.4* and *Figure S5.4*). This might suggest a reduction in SICI up to ∼6 weeks following burn injury, which is normalised by ∼12 weeks following burn injury; however, this finding must be interpreted with caution as the burn samples across timepoints are semi-independent and only a single control timepoint was included in the analysis. In the current study, slope-based analysis revealed no change in SICI slope ratios over time. The same pattern of results was observed in the subgroup analysis, which only included participants with an upper limb burn (see Supplementary Materials Section *S6.1*). Previous research has shown no difference in SICI between burn patients 1–3 years following injury and controls (33). Together, these findings suggest GABA_A_-mediated intracortical inhibition remains largely unchanged following burn injury.

Baseline LICI amplitude ratios remained stable across time, suggesting no changes in the excitability of GABA_B_-mediated inhibition between ∼3 and ∼12 weeks following minor burn injury. While there was no significant difference in LICI ∼3 to ∼12 weeks following injury, cross-sectional analysis comparing burn patients and non-injured controls showed LICI was greater for controls than burns patients at ∼6 weeks post-injury (adjusted *p* = .039) but not ∼12 weeks post-injury (see Supplementary Materials Section *S5.4* and *Figure S5.4*). This might suggest a reduction in LICI ∼6 weeks following burn injury, which is normalised by ∼12 weeks following burn injury; however, this finding must be interpreted with caution as the burn samples across timepoints are semi-independent and only a single control timepoint was included in the analysis. Previous research examining cortical silent period (cSP) duration, which reflects both GABA_A_- and GABA_B_-mediated inhibition (80), reported no significant differences between burn participants and non-injured controls (32). It is worth noting, however, that Garside et al. (32) reported exploratory subgroup analyses (n = 13) showing shorter cSP durations on the burn-affected side in individuals with upper-limb burns, suggesting a reduction in long-acting inhibition following burn injury. In the current study, subgroup analysis on upper limb participants showed no change in LICI from ∼3 to ∼12 weeks following injury. These inconsistent findings could be due to differences in time since injury of the two studies: while the current study longitudinally measured LICI from ∼3 to ∼12 weeks following injury, the participants in Garside et al. (32) ranged from 6 weeks to 8 years following burn injury. It is also possible that there are differences in inhibition when measured during voluntary activation between burn patients and controls. Future research should examine task-related changes in intracortical inhibition in burn patients. The slope-based analysis in the current study revealed a significant reduction in LICI slope ratios over time, suggesting a time-dependent increase in GABA_B_-mediated inhibition from ∼3 to ∼12 weeks following minor burn injury. Taken together, these results suggest that while baseline intracortical inhibition remains stable at the output level, slope-based analyses may capture time-dependent changes in cortical inhibitory function during the subacute period.

### Greater Baseline SICI Associated with Poorer Bilateral Performance Early in Recovery

A positive association between SICI and simple bilateral pegboard performance was observed at day 23, suggesting that individuals with greater SICI performed poorly on the simple bilateral subtest early in recovery, at ∼3 weeks following injury, but this was not demonstrated at ∼6 or ∼12 weeks following injury. Although there was no change in SICI from ∼3 to ∼12 weeks following injury, this finding suggests that, at an individual level, the magnitude of SICI is associated with bilateral performance. In burn patients 1–3 years following injury (33), and in non-injured individuals (81), greater SICI is associated with better performance on the Purdue pegboard. Taken together, these findings suggest that the functional role of GABA_A_-mediated inhibition differs across the subacute and chronic periods following a burn injury, with an atypical association of greater GABA_A_-mediated inhibition and poorer manual dexterity very early following a burn injury, which normalises to the typical association of greater GABA_A_-mediated inhibition and better manual dexterity 1–3 years following a burn injury. Future research should examine whether early-recovery SICI and manual dexterity are predictive of long-term post-burn outcomes.

In this study, we found no evidence of an association between LICI and manual dexterity. Prior work shows that greater LICI 1–3 years post-injury is linked to better dexterity (33), suggesting GABA_B_-mediated inhibition may relate to motor performance in the chronic—but not subacute—period. These null results should be interpreted cautiously: the study was powered to detect differences, not equivalence, and the modest sample yields wide CIs, so small effects cannot be excluded. We also found no evidence that SICI or LICI were associated with SF36 Physical or Mental Component Summary scores from ∼3 to ∼12 weeks post-injury, indicating no detectable link between GABAergic inhibition and self-reported general function in the subacute period.

### MEP Unchanged and LICI Increased Following PAS

Following PAS, single-pulse MEP amplitude and slope ratios remained stable, suggesting no systematic change in corticospinal excitability induced by PAS ∼3 to ∼12 weeks following burn injury. In the current study, PAS was chosen to assess LTP-like plasticity due to its role in sensory-motor integration (51), which may be particularly relevant following burn injury where sensory and motor dysfunction is common, and its effectiveness in increasing MEP amplitude (48). Therefore, the current findings suggest PAS-induced LTP-like plasticity is reduced or impaired in the subacute period following burn injury and extends previous work showing no evidence of PAS-induced LTP-like plasticity in burn patients 1–3 years following injury (33). Previous research examining neuroplasticity in M1 in the subacute period following burn injury has shown reduced long-term depression (LTD)-like plasticity induced by spaced continuous theta burst stimulation (cTBS) at 6 weeks but not at 12 weeks post-injury (82), suggesting a reduced capacity for LTD-like neuroplasticity ∼6 weeks following burn injury, which normalises by ∼12 weeks following burn injury. Cross-sectional analysis comparing burn patients and non-injured controls showed a significantly greater PAS-induced increase in MEP amplitude in controls than burn patients ∼12 weeks following injury (see Supplementary Materials Section *S5.5* and *Figure S5.5.1*), which might suggest reduced PAS-induced LTP-like plasticity at ∼12 weeks following burn injury. However, this finding must be interpreted with caution as the burn samples across timepoints are semi-independent and only a single control timepoint was included in the analysis. Furthermore, it is important, however, to consider these findings of no PAS-induced change in corticospinal excitability in the background context of inconsistent results from previous PAS studies in healthy individuals: some studies have shown increased MEP amplitude post-PAS [for example see: (51)] others have shown no change in MEP amplitude post-PAS [for example see: (83)], and there are reports of high response variability, with only ∼50% of participants responding as expected (84).

In the current study, there was no PAS-induced change in SICI amplitude or slope ratios ∼3 to ∼12 weeks following burn injury, suggesting no modulation of GABA_A_-mediated inhibition by PAS. This is consistent with research in healthy individuals, showing no change in SICI following PAS (85–87) and previous work in burn patients 1–3 years following burn injury (33). Taken together, this suggests that PAS does not modulate the excitability of GABA_A_-mediated inhibitory circuits in M1, and this is not affected in either subacute or chronic post-injury periods for burn patients.

Following PAS, there were significant changes in LICI amplitude and slope ratios between ∼3 and ∼12 weeks post-injury, with amplitude ratios showing increased LICI at 15 and 30 minutes post-PAS ∼6 weeks following injury, indicative of enhanced GABA_B_-mediated inhibition (24). This finding of increased LICI post-PAS is consistent with previous research showing a short-lasting increase in LICI post-PAS (e.g., at 0 and 15 minutes post-PAS) in burn patients 1–3 years post-minor injury (33), and research in healthy young individuals that has shown increased LICI following PAS (88). In the current study, non-injured controls also showed an increase in LICI following PAS (see Supplementary Materials Section *S5.5* and *Figure S5.5.3*). It is important to note that one previous study found no change in LICI following PAS in healthy adults when using stimulation parameters comparable to the current study (89), suggesting changes in LICI following PAS might show high inter-individual variability in responses, similar to the changes in MEP amplitude following PAS. Taken together, these findings suggest the PAS-induced increase in GABA_B_-mediated inhibition is typical in burn patients in both the subacute and chronic recovery periods.

Slope-based metrics revealed a progressive reduction in the PAS-induced increase in LICI over time, with slopes becoming less negative from ∼3 to ∼12 weeks across all post-PAS blocks. In line with the significant rises in slope from day 23 to 82 (and from day 23 to 44 at 30 min post-PAS), these patterns indicate that PAS-induced modulation of GABA_B_-mediated inhibition varies within the first ∼12 weeks after injury, with the PAS-related increase in LICI weakening over time.

### Associations Between PAS-induced LICI Increase and Function

A significant negative association between PAS-induced increase in LICI and simple bilateral pegboard performance was evident ∼6 weeks following injury, suggesting that greater modulation of GABA_B_-mediated inhibition is associated with better bilateral performance. Similar associations were evident ∼3 weeks and ∼12 weeks following burn injury, though they did not reach statistical significance, suggesting the association is strongest ∼6 weeks following burn injury. As described above, greater baseline, resting state LICI 1–3 years following burn injury is associated with better motor performance (33). While the current study showed no association between baseline LICI and motor performance in the subacute period, it is interesting to speculate whether those who show the greatest PAS-induced change in LICI, potentially reflecting the greatest capacity for plasticity of LICI circuits, show the best recovery of motor performance in the chronic period following burn injury. Further research is needed to systematically investigate the potential of PAS-induced LICI modulation as a biomarker of functional recovery following burn injury.

In the current study, there were no significant associations between PAS-induced neuroplasticity and self-reported physical or mental health outcomes, suggesting no relationship between capacity for LTP-like plasticity and self-reported general function in this cohort. Previous research showed an association between LTD-like plasticity (induced by spaced cTBS) and general health perception scores ∼12 weeks following injury, suggesting those subacute burn patients with a greater plasticity response have the best recovery within this timeframe (82). Together, these findings might suggest the distinct neurophysiological processes of LTP and LTD plasticity are differentially associated with self-report functional recovery (90). It is also possible that the minor nature of the burn injuries in this cohort contributed to these results by limiting variability and reducing the magnitude of detectable associations.

### Limitations and Future Directions

While the current study provided the first comprehensive characterisation of neurophysiological changes in the subacute period following burn injury, there are some limitations that should be considered when interpreting the findings. First, there was considerable variation in the assessments relative to the date of injury. While this is somewhat expected when working with subacute burn injury patients, and the selected timepoints (23, 44, and 82 days following injury) align with periods of highest testing density and clinically relevant milestones, this variation relative to date of injury introduces some uncertainty into the estimated trajectories, particularly at the edges of the observation window where data were sparse. In addition, some participants only completed assessments ∼3 and ∼6 weeks following injury while others only completed assessments ∼6 and ∼12 weeks following injury, introducing imbalance in the data, with different participants contributing to different segments of the recovery timeline. To mitigate this, all analyses included participant-level random effects, which account for within-subject correlation across repeated measures and help control for between-subject variability arising from the sampling imbalance. However, estimates presented here should be interpreted with some caution. Notwithstanding the challenges of clinical research, future studies should take a structured approach to evenly distribute assessments across the target recovery period.

Second, in the PAS protocol, a standard interval of 25 ms was used between the peripheral stimulus and the TMS to M1: this approach does not account for individual differences in conduction time and might result in suboptimal timing for PAS plasticity induction (91). In addition, for three participants with wrist burns, the peripheral stimulating electrodes were placed on the palmar surface of the third digit on the most affected side, rather than over the median nerve at the wrist. This electrode placement stimulated the median nerve via the palmar digital branches (52), however, would have affected the precise timing of the PAS protocol, potentially affecting PAS-induced plasticity. Moreover, PAS was administered at various times of the day, which could have introduced variability in the PAS response due to circadian fluctuations (86, 92). Participants also reported increased sleepiness following PAS, with the most pronounced changes occurring immediately post-stimulation. This transient reduction in arousal may have influenced neuroplasticity, given that attentional state is known to modulate neuroplasticity responses to non-invasive brain stimulation (53, 93).

Third, during the pegboard tasks, several participants with hand burns noted that injury or dressings interfered with performance, introducing variability in motor scores unrelated to neurophysiological function. Separately, to ensure deficits on the most-affected side were reflected in the outcome, we emphasised bilateral pegboard performance in the main analyses; for the same reason—and because subgrouping by most-versus least-affected hand would have been underpowered—we did not stratify results by combinations of burn side and handedness.

Fourth, both pre-injury and post-injury medications, including pain relief and antidepressants, which act on the central nervous system (94), as well as surgical treatment for the burn injury (95), may have influenced the observed findings. Future studies should record these and account for these potential confounders by stratifying participants based on pain or medication use or incorporating these factors as covariates in analyses.

Finally, while the primary research question focused on changes in neurophysiology and function during the subacute period following burn injury, no age- and sex-matched non-injured control group was included. To address this, we conducted additional cross-sectional analyses in a subsample of participants with burn injury who had at least one session within prespecified post-injury windows (3 weeks: 14–28 days; 6 weeks: 35–49 days; 12 weeks: 77– 91 days). These data were compared with non-injured controls using a categorical statistical approach. The results of these cross-sectional analyses, along with corresponding figures, are presented in the supplementary materials.

### Conclusions

Subacute burn patients with minor injuries showed greatest improvements in motor performance and self-reported physical health from ∼3 to ∼6 weeks following injury. Changes in GABA_B_-mediated inhibition were evident in the first ∼12 weeks following burn injury: baseline resting-state LICI inhibitory influence increased over the first 12-weeks following injury, and PAS induced an increase in LICI, which was greatest at ∼6 weeks following injury and was significantly associated with bilateral motor performance. These findings suggest that burn patients who show stronger modulation of GABA_B_-mediated inhibition in response to PAS, potentially reflecting greater plasticity within these circuits, might have enhanced opportunity for motor recovery in the subacute phase following burn injury. Future research should comprehensively investigate GABA_B_-mediated inhibition as a marker of functional recovery following burn injury

## Data Availability

Primary data and code are available at OSF (view-only link: https://osf.io/n3euw/overview?view_only=a67c490f40864baea2dfce1ba1426490) and in Supplementary Materials (Section S6).

