## Supplementary Materials for "Altered Sensorimotor Neuroplasticity in the Subacute Period Following Burn Injury"

**S1: UNILATERAL PURDUE PEGBOARD PERFORMANCE IN SUBACUTE BURN PATIENTS**

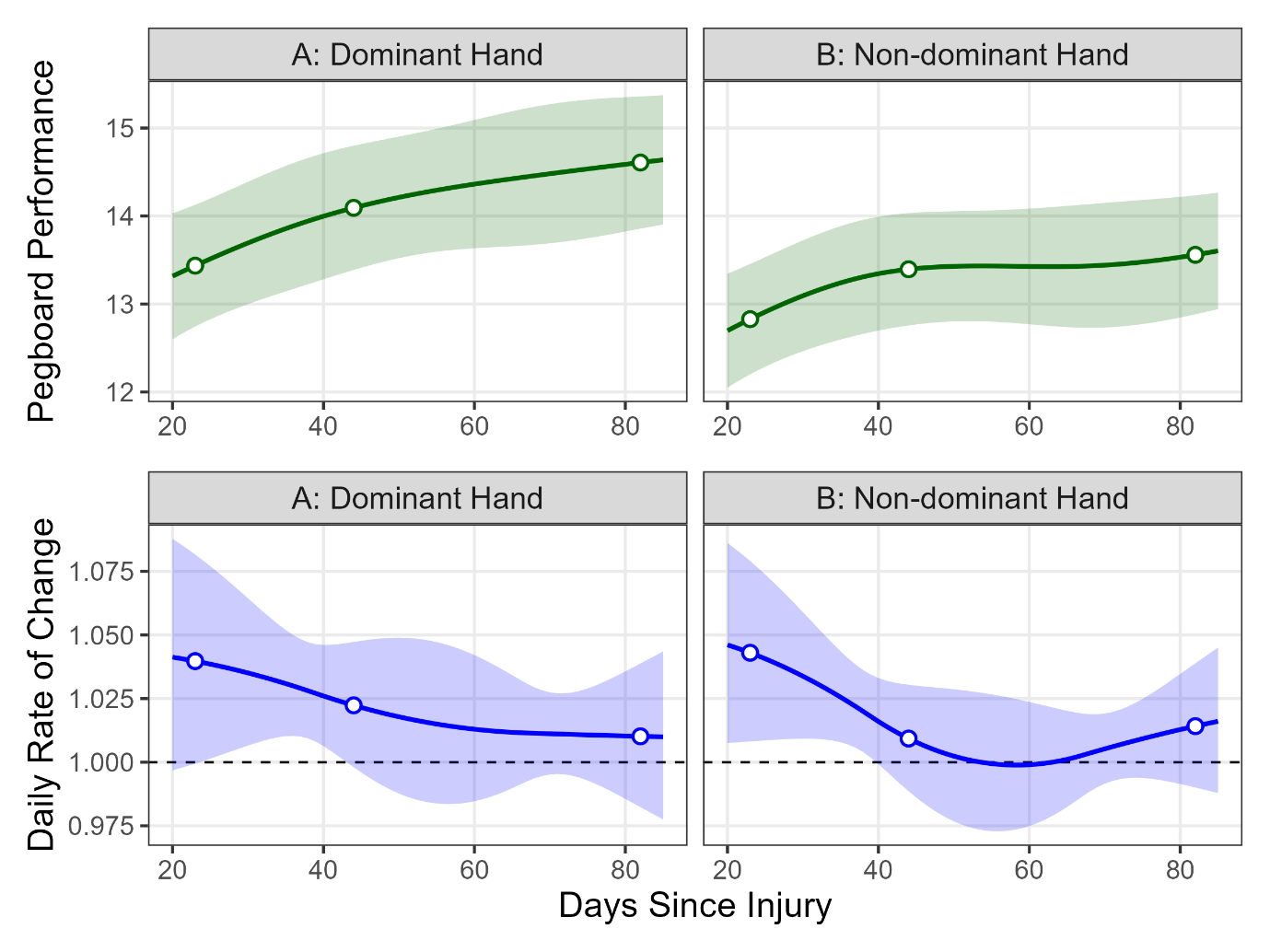

Supplementary Figure S1. Modelled trajectories of pegboard performance and daily rate of change following burn injury. Panels A and B show predicted pegboard task performance for the dominant (A) and non-dominant (B) hands across days since injury. Solid lines represent estimated trajectories, open circles show estimates at 23, 44, and 82 days post-injury, and shaded areas indicate 95% confidence intervals. Panels C and D display the estimated daily rate of change in performance for the dominant (C) and non-dominant (D) hands. The dashed horizontal line at 1.0 represents no change: values >1.0 indicate improvement, while values <1.0 indicate decline. Shaded areas represent uncertainty in the estimates.

**S2: BASELINE TMS INTENSITIES IN SUBACUTE BURN PATIENTS**

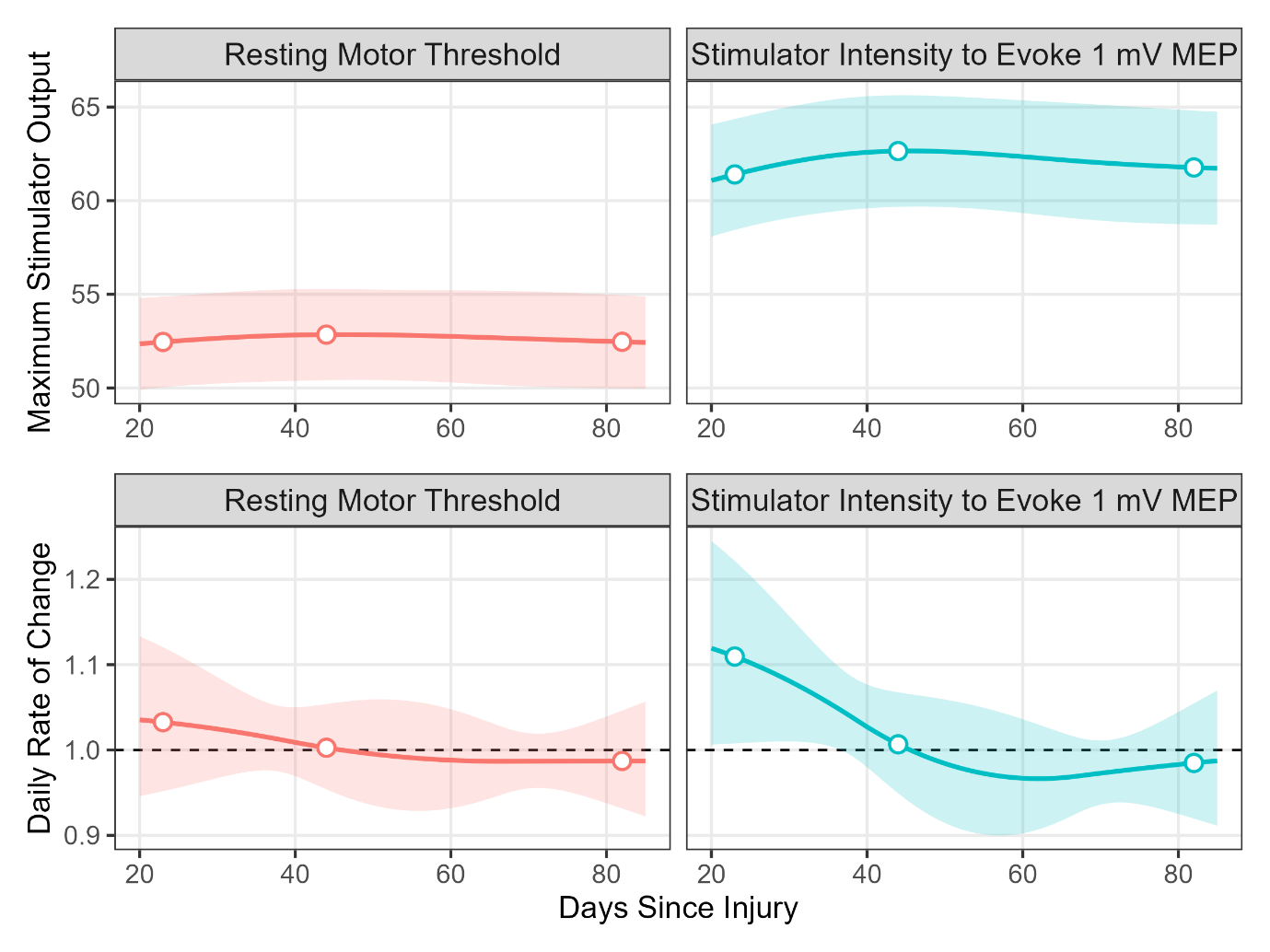

Supplementary Figure S2. Modelled trajectories of resting motor threshold (RMT; A) and stimulator intensity required to evoke a 1 mV MEP (SI_1mV_; B) across days since injury (top panels). Solid lines represent estimated marginal means, open circles indicate predicted values at 23, 44, and 82 days post-injury, and shaded areas denote 95% confidence intervals. Bottom panels (C, D) show the estimated daily rate of change in RMT and SI_1mV_. The dashed horizontal line at 1.0 indicates no change: values >1.0 reflect an increase in stimulator output over time, and values <1.0 reflect a decrease. Shaded areas represent uncertainty in the estimates.

**S3: ASSOCIATIONS BETWEEN CHANGES IN SINGLE-PULSE MEP AMPLITUDE FOLLOWING PAS AND BILATERAL PEGBOARD PERFORMANCE**

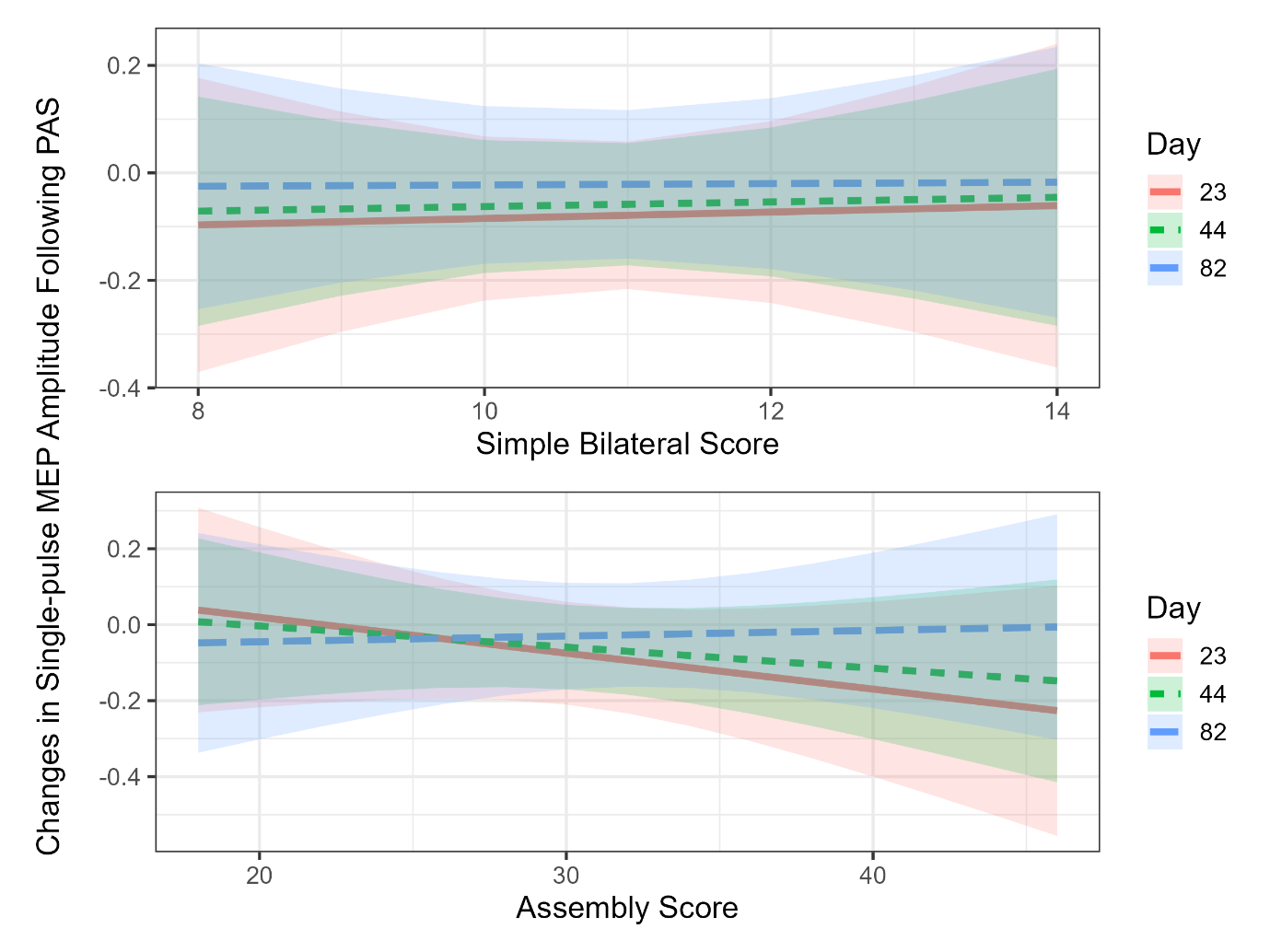

A

B

Supplementary Figure S3. Associations between pegboard subtest performance and paired associative stimulation (PAS)-induced changes in single-pulse MEP amplitude across recovery timepoints. Panel A shows the relationship between simple bilateral scores and MEP changes, with no significant differences in slope between days 23, 44, and 82 (all *p* ≥ .994). Panel B shows the corresponding relationship for assembly scores, where slopes became progressively less negative over time, but differences between days were also not statistically significant (all *p* ≥ .609). Shaded areas indicate 95% confidence intervals.

**S4 FUNCTIONAL ASSESSMENTS: METHODS AND RESULTS**

For the self-report assessments (*S4.1-S4.5* outlined below), participants were asked to complete the surveys remotely via an email link (REDCap Electronic Database, USA) within 24 hours either side of each session. For the sensory assessments (*S4.7-S4.10* outlined below), the testing locations were identified on the skin with a surgical marker approximately 3 cm away from the wound or scar (wound was classified as a scar once the clinical dressings were removed) at these locations: above the scar on the most-affected side; above homologue on least-affected side; below the scar on the most-affected side; and below homologue on least-affected side. When the burn injury was located on or around sensitive body areas such as genital or face some sensory measurements were not collected.

**Survey Data Analysis.** Burn-specific Health Scale-Brief, QuickDASH, Lower Limb Functional Index (LLFI), and Patient and Observer Scar Assessment Scale (POSAS) scores were processed according to their respective scoring rules, transformed to a 0–100 scale, and analysed using the same modelling approach. Linear mixed-effects models with natural cubic splines for days since injury (df = 3) and participant-level random intercepts were fit. Estimated marginal means were extracted at 23, 44, and 82 days post-injury, with pairwise contrasts between timepoints Holm-adjusted for multiple comparisons. Results are reported as model-predicted percentage changes with adjusted *p*-values.

painDETECT total scores were constructed according to scoring rules (including radiating and course recodes) and analysed on the original scale. A linear mixed-effects model with a natural cubic spline for days since injury (df = 3) and participant-level random intercepts was fit. Estimated marginal means were obtained at 23, 44, and 82 days, and Holm-adjusted pairwise contrasts were calculated. Percent change versus the earlier timepoint was also computed for interpretability.

**Sensory Function Data Analysis.** Monofilament threshold, pain pressure threshold, grip strength, Neuropen perception testing, and visual analogue scale outcomes were analysed using the same approach. For each measure, we fit linear mixed-effects models with days since injury modelled as a natural cubic spline (df = 3) and participant-level random intercepts to account for repeated measures and capture non-linear recovery. Monofilament thresholds were modelled on the log10 scale to account for the discrete, stepwise nature of the measure, with results back-transformed to the original scale for interpretability. Estimated marginal means were obtained at 23, 44, and 82 days post-injury, with pairwise contrasts Holm-adjusted for multiple comparisons. Results are reported as model-predicted percent changes relative to the earlier timepoint, with adjusted *p*-values.

Responses to brush perception testing were summarised descriptively rather than modelled. Days since injury were grouped into three windows (0–30, 31–60, and >60 days). Within each window, the frequency and percentage of each response category were calculated for each variable. Results are reported as counts and percentages of observed responses across the recovery windows.

***S4.1 Burn-specific Health Scale-Brief***

Methods. The Burn-specific Health Scale-Brief was used to assess burn-related health status (1). Two domain scores were calculated: the generic domain, reflecting psychological and social well-being, and the physical domain, representing physical functional abilities. Each item was rated on a 0–4 scale, with anchors ranging from 0 = extremely to 4 = not at all, indicating the extent to which a difficulty or limitation was experienced. Domain scores were derived by summing item responses, with higher scores reflecting better perceived health. To aid interpretation and comparability, raw domain sums were transformed to a standardised 0–100 scale.

Results. Figure S4.1 highlights the estimated marginal means (±95% CI) for Burn-specific Health Scale-Brief percentage scores at 23, 44, and 82 days post-injury: (A) Physical Function domain and (B) Generic domain. A total of 31 participants contributed 79 observations for both the physical and generic domains of the Burn-specific Health Scale-Brief. The physical and generic percentage scores were modelled across 23, 44, and 82 days post-injury. For the physical percentage score, estimates suggested small, non-significant improvements over time, with a +6.4% increase from day 23 to 44 (adjusted *p* = .410) and a +7.2% increase from day 23 to 82 (adjusted *p* = .534). No significant change was observed between days 44 and 82 (+0.8%, adjusted *p* = .896). For the generic percentage score, a modest but significant improvement was observed from day 23 to 44 (+2.1%, adjusted *p* = .047). However, changes from day 23 to 82 (+1.1%, adjusted *p* = .772) and from day 44 to 82 (–1.0%, adjusted *p* = .772) were not significant. Generic percentage scores therefore showed early psychosocial gains that plateaued by 12 weeks post-injury.

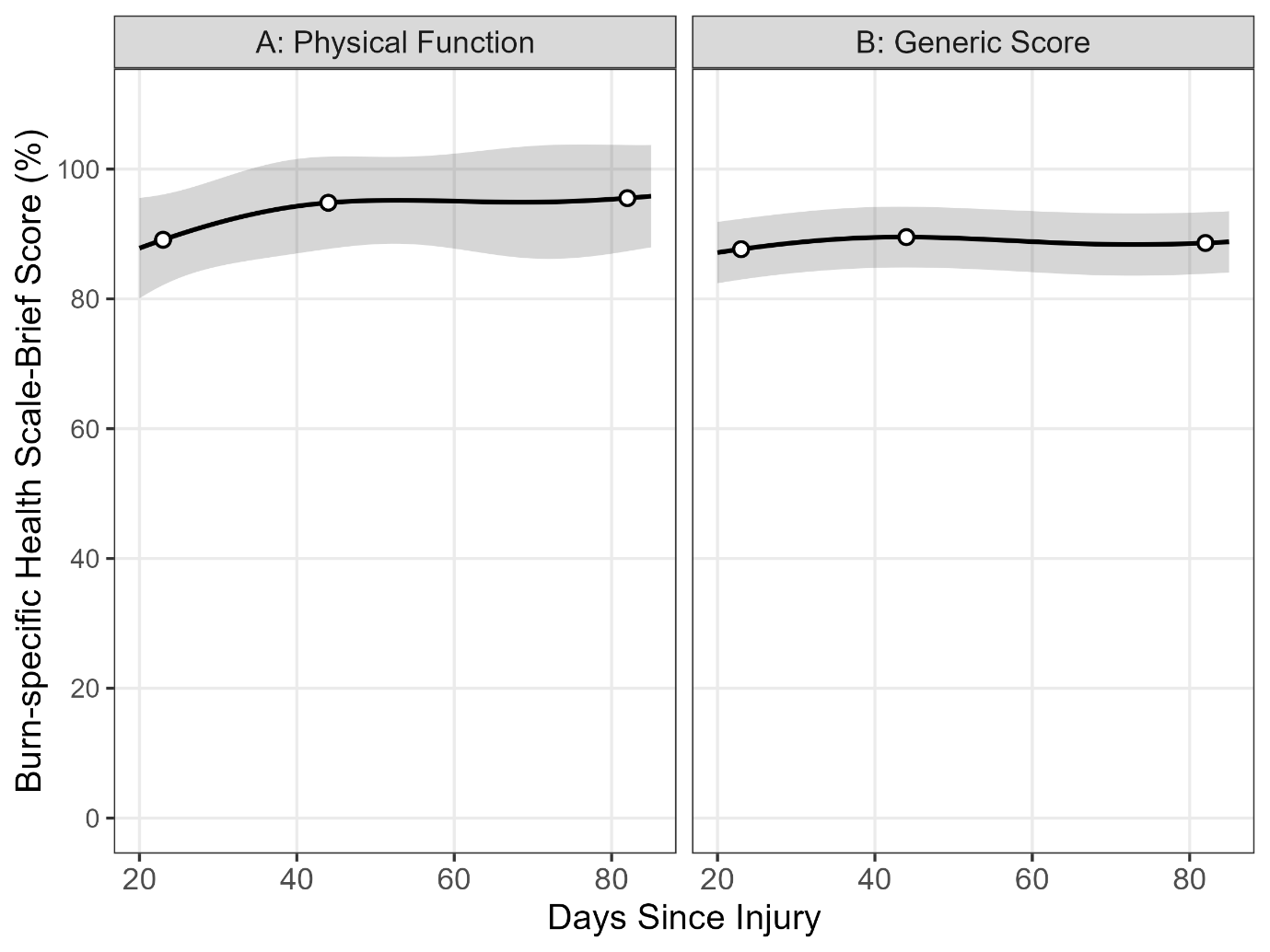
Figure S4.1. Estimated marginal means (±95% CI) for Burn-specific Health Scale-Brief percentage scores across 23, 44, and 82 days post-injury: (A) Physical Function domain, (B) Generic domain. Both domains showed high levels of function. Physical Function scores showed small, non-significant improvements over time (all adjusted *p* ≥ .410). Generic scores demonstrated a modest improvement from day 23 to 44 (+2.1%, adjusted *p* = .047), but no further significant change to day 82.

***S4.2 QuickDASH***

Methods. Participants rated their difficulty in performing specific activities and the severity of symptoms (e.g., pain, weakness) on a 5-point Likert scale: 1 = no difficulty or symptom, 5 = extreme difficulty or symptom (2). The QuickDASH score was calculated by averaging the item scores, dividing by the number of completed items, and multiplying the result by 25 to yield a score between 0 and 100, with higher percentages indicating greater self-perceived disability and symptom severity.

Results. Figure S4.2 shows the estimated marginal means (±95% CI) for QuickDASH disability percentage scores across 23, 44, and 82 days post-injury. A total of 31 participants contributed 78 observations to the QuickDASH analysis. QuickDASH disability scores were modelled across 23, 44, and 82 days post-injury. Scores decreased substantially from day 23 to 44 (−47.8%, adjusted *p* = .002) and from day 23 to 82 (−52.9%, adjusted *p* = .015), indicating marked improvements in upper-limb function. There was no further change between days 44 and 82 (−9.7%, adjusted *p* = .785).

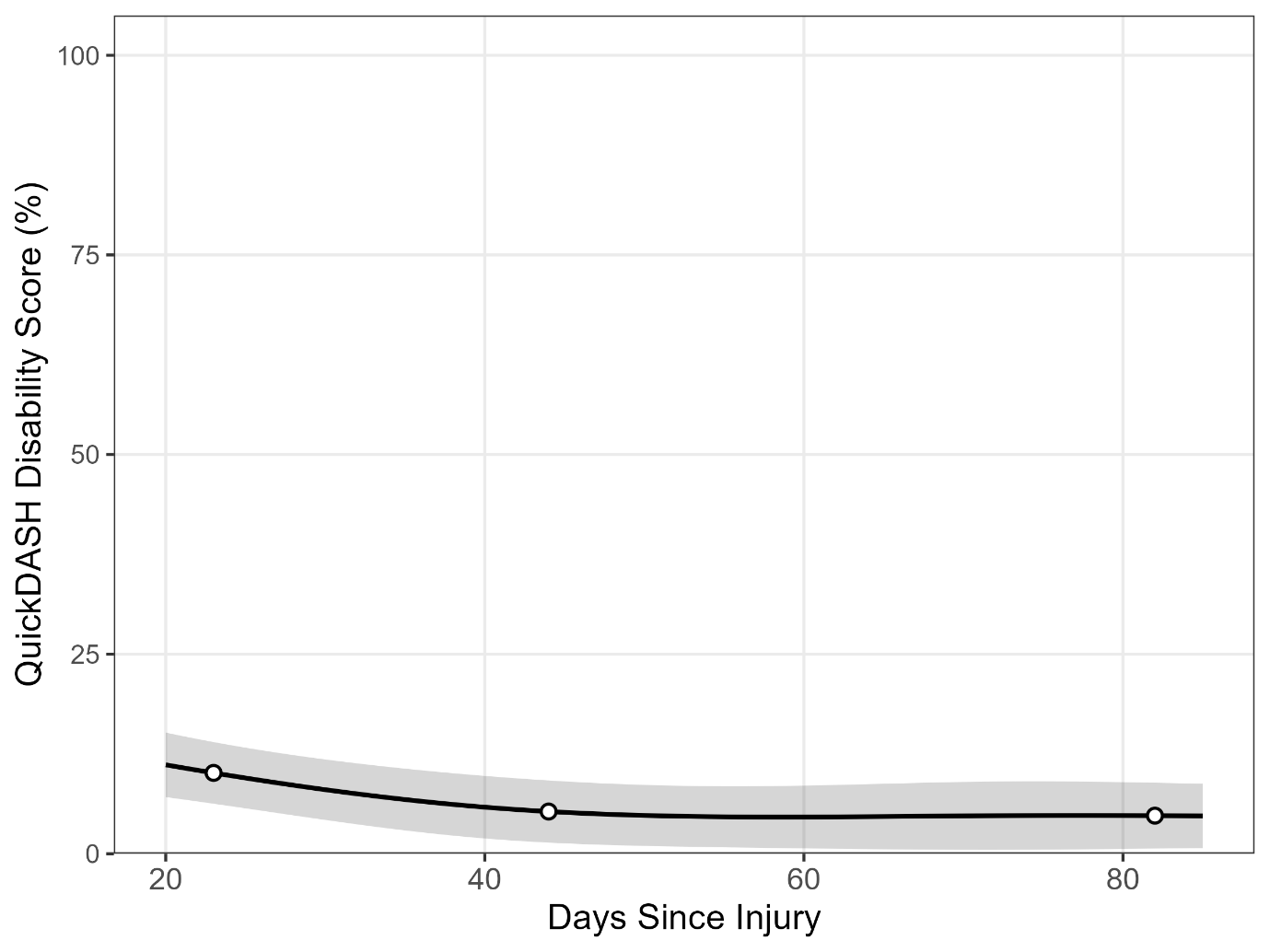

Figure S4.2. Estimated marginal means (±95% CI) for QuickDASH disability percentage scores across 23, 44, and 82 days post-injury. Scores decreased substantially from day 23 to 44 (−47.8%, adjusted *p* = .002) and from day 23 to 82 (−52.9%, adjusted *p* = .015), indicating marked improvements in upper-limb function. No further significant change was observed between days 44 and 82.

***S4.3 Lower-limb Functional Index (LLFI)***

Methods. Participants with lower-body injuries completed the 10-item Lower Limb Functional Index to rate their difficulty performing daily activities and the frequency of pain (3). Each item was scored on a 3-point scale, with lower scores indicating better function. The total score, ranging from 0 to 10, was then converted to a percentage for analysis.

Results. Figure S4.2 highlights the estimated marginal means (±95% CI) for QuickDASH disability percentage scores from 20–85 days post-injury, with highlighted points at days 23, 44, and 82. The Lower-limb Functional Index percentage scores (11 participants, 28 observations) were modelled across 23, 44, and 82 days post-injury. Scores declined from day 23 to 44 (−34.6%, adjusted *p* = .105) and from day 23 to 82 (−34.4%, adjusted *p* = .528), indicating non-significant reductions in self-reported lower-limb function. There was no further change between days 44 and 82 (+0.3%, adjusted *p* = .994).

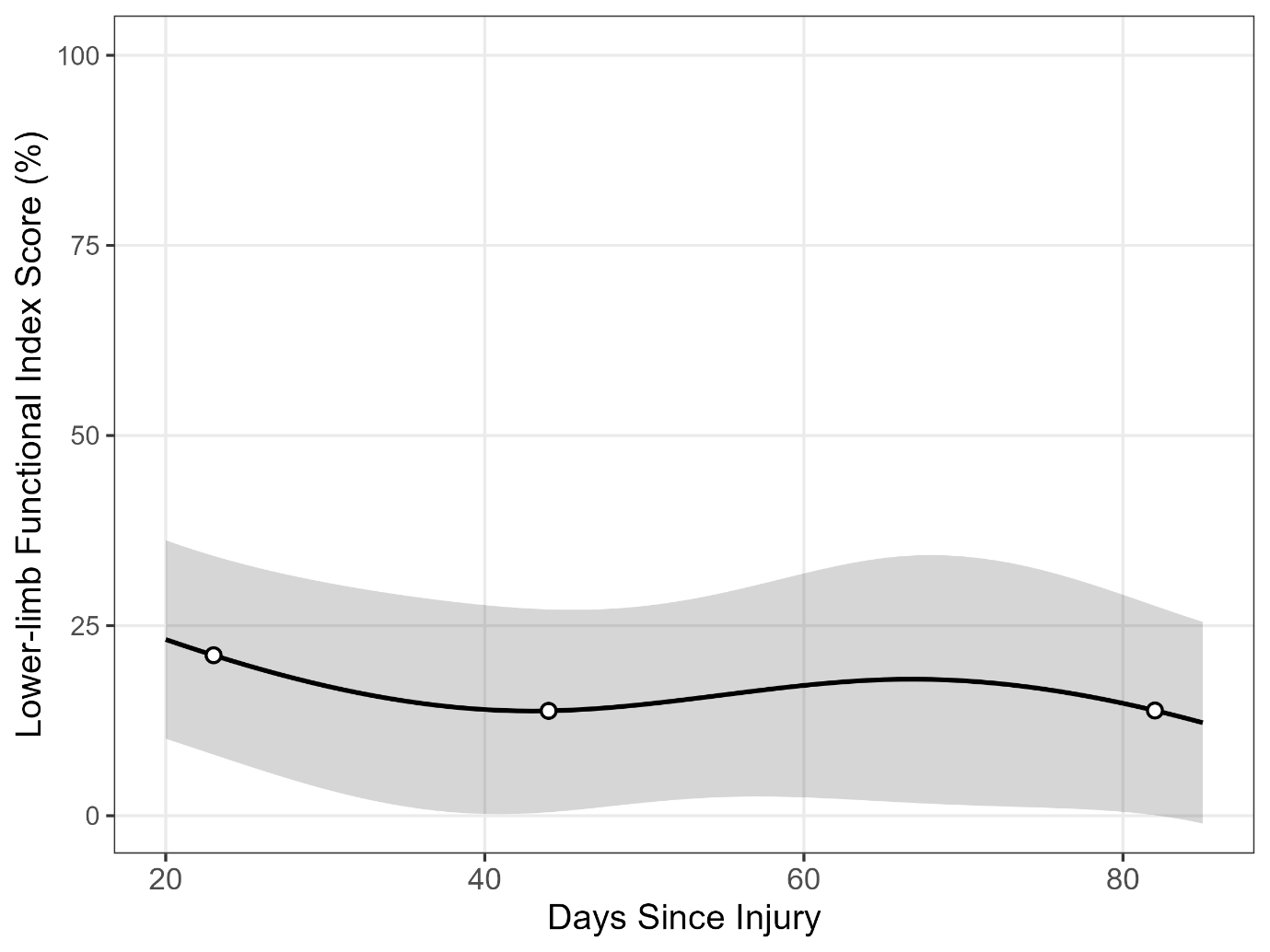

Figure S4.3. Estimated marginal means (±95% CI) are shown for Lower-limb Functional Index percentage scores from 20–85 days post-injury, with highlighted points at days 23, 44, and 82. Scores showed a downward trend from day 23 to 44, consistent with improved lower-limb function, before remaining stable through day 82. However, no pairwise comparisons between timepoints were statistically significant (all *p* ≥ .105).

***S4.4 Patient and Observer Scar Assessment Scale (POSAS)***

Methods. Participants completed the Patient and Observer Scar Assessment Scale, with scars defined as present once clinical dressings were removed (4). The patient scale included six items evaluating the scar from the patient’s perspective: pain, itchiness, colour, stiffness, thickness, and irregularity. Each item was scored on a 10-point scale (1 = normal skin, 10 = very different to normal skin), with lower scores reflecting better scar perception. Item scores were converted to a standardised 0–100 scale to aid interpretation and comparability.

Results. Estimated marginal means (±95% CI) for Patient and Observer Scar Assessment Scale percentage scores are shown from 20–85 days post-injury, with highlighted estimates at 23, 44, and 82 days (Figure S4.4). A total of 29 participants contributed 60 observations to the Patient and Observer Scar Assessment Scale analysis. The percentage scores were modelled across 23, 44, and 82 days post-injury. Scores declined from day 23 to 44 (−23.7%, adjusted *p* = .097) and from day 23 to 82 (−44.4%, adjusted *p* = .007), indicating a significant reduction in scar severity by 12 weeks. There was no further significant change between days 44 and 82 (−27.0%, adjusted *p* = .097).

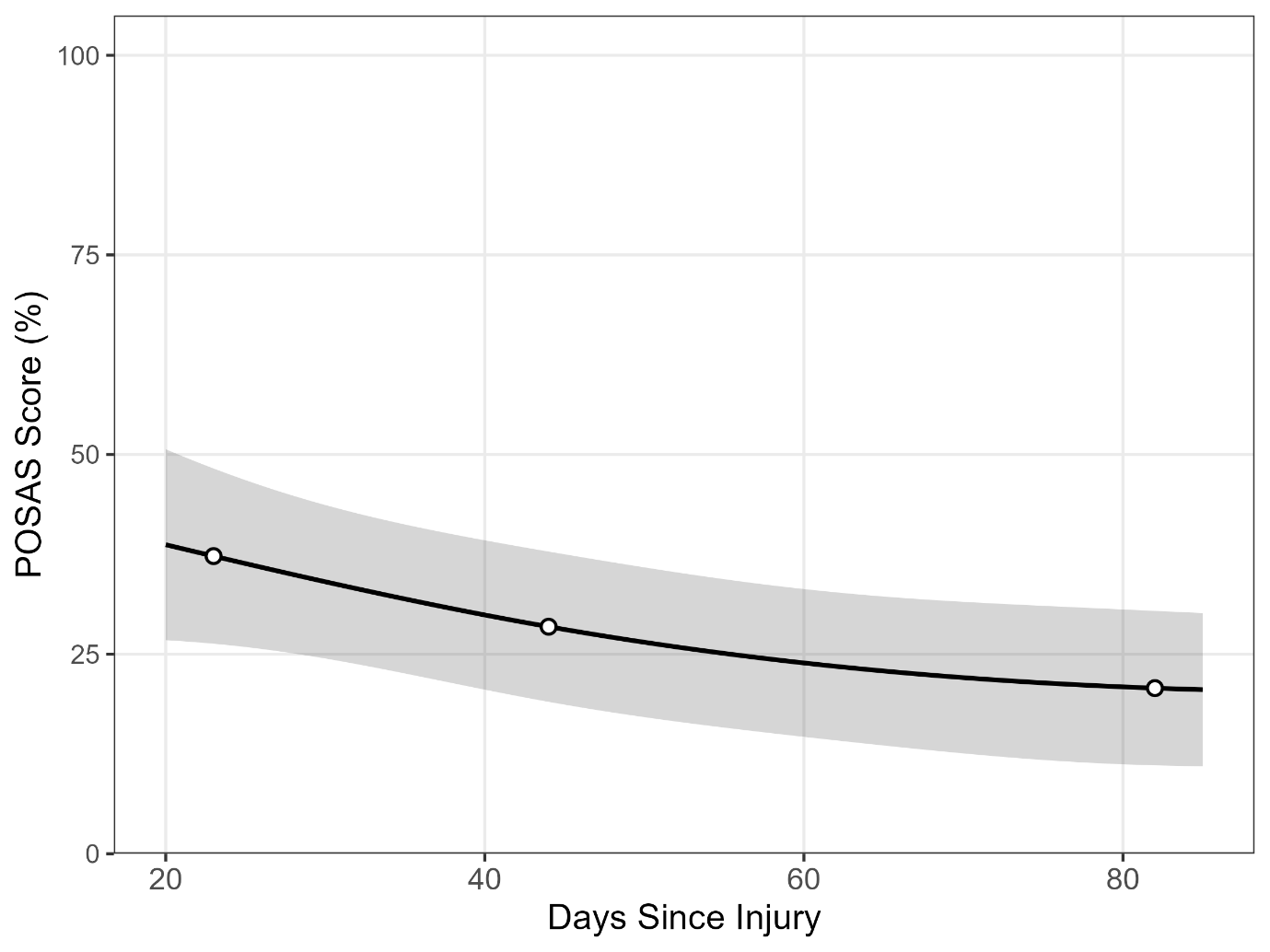

Figure S4.4. Estimated marginal means (±95% CI) are shown for Patient and Observer Scar Assessment Scale percentage scores from 20–85 days post-injury, with highlighted estimates at days 23, 44, and 82. Patient and Observer Scar Assessment Scale scores declined steadily over time, consistent with improving scar quality. Pairwise contrasts indicated a significant reduction between days 23 and 82 (adjusted *p* = .007), while differences between days 23 and 44 and between days 44 and 82 did not reach statistical significance.

***S4.5 PainDETECT***

Methods. The scores from the sensory descriptor items were summed, along with additional points from the pain pattern and radiation items, resulting in a total score ranging from -1 to 38 (5). Interpretation of the total score: 0–12: nociceptive pain with neuropathic pain component unlikely, 13–18: result is ambiguous; a neuropathic pain component can be present, ≥19: neuropathic pain component is likely.

Results. Estimated marginal means (±95% CI) for painDETECT scores are shown from 20–85 days post-injury, with highlighted estimates at 23, 44, and 82 days (Figure S4.5). A total of 21 participants contributed 32 observations to the analysis (some participants did not complete the survey because they reported no pain). The painDETECT total scores were modelled across 23, 44, and 82 days post-injury. Scores remained stable from day 23 to 44 (+0.7%, adjusted *p* = .975). By day 82, scores had increased relative to both day 23 (+60.0%, adjusted *p* = .134) and day 44 (+59.0%, adjusted *p* = .134), indicating a non-significant trend toward greater neuropathic pain features over time.

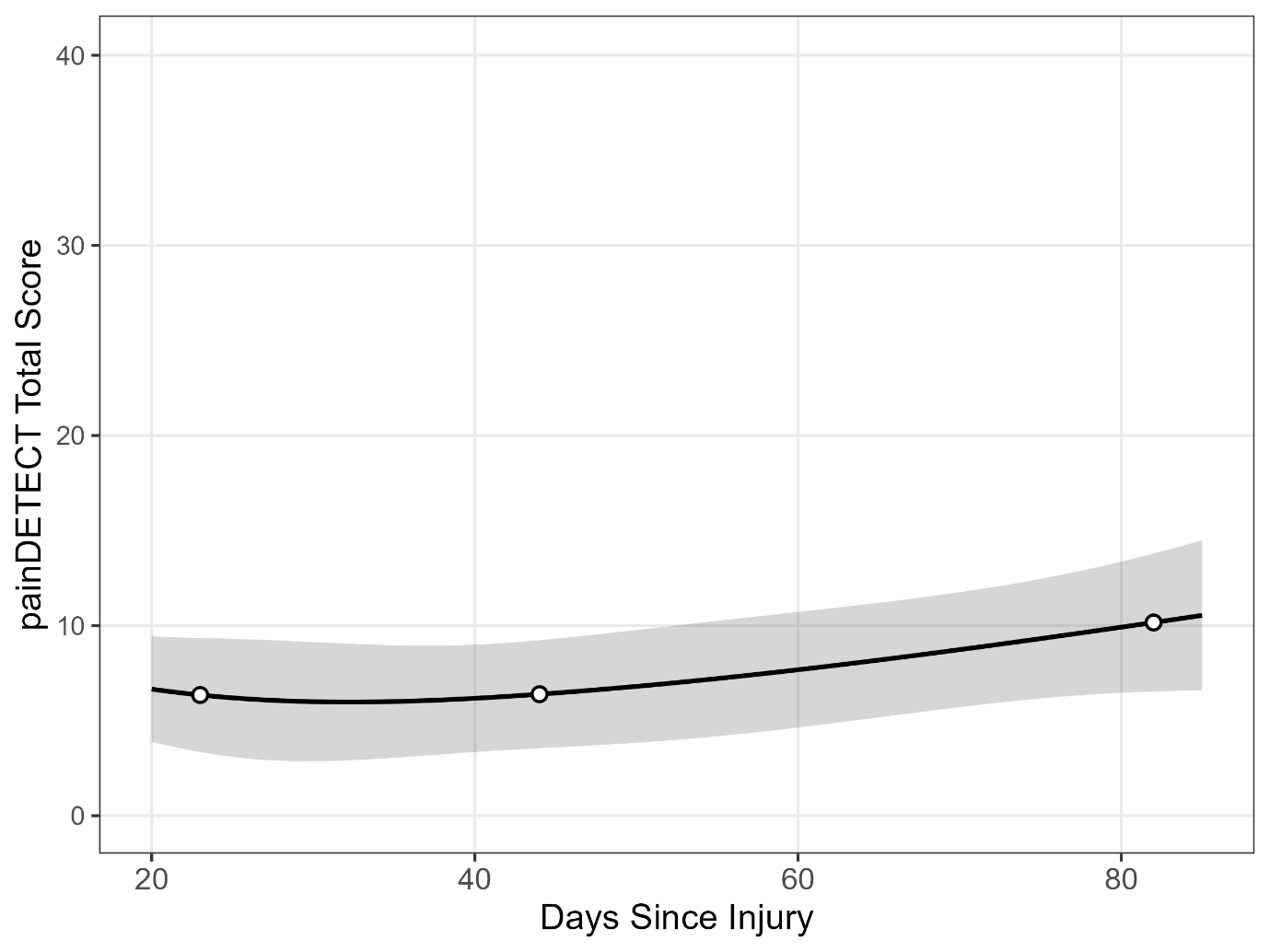

Figure S4.5. Estimated marginal means (±95% CI) are shown for painDETECT scores from 20–85 days post-injury, with highlighted estimates at days 23, 44, and 82. Scores remained stable between days 23 and 44, before showing an upward trend by day 82. Pairwise contrasts indicated no statistically significant differences between timepoints (all *p* ≥ .134).

***S4.6 Visual Analogue Scale: Burn-related Pain***

Methods. A visual analogue scale for pain was used to measure the level of burn injury-induced pain experienced at the time of testing. Participants were instructed to use a marker to indicate on a 100 mm line their level of pain ranging from 0 (no pain) to 100 (pain as bad as it could possibly be).

Results. Perceived burn-related pain ratings are shown across recovery, with model-predicted Visual Analogue Scale scores (black line ±95% CI) from 20–85 days post-injury and highlighted estimates at 23, 44, and 82 days (Figure S4.6). A total of 34 participants contributed 90 observations across the experimental period. The Visual Analogue Scale scores for burn-related pain were modelled across 23, 44, and 82 days post-injury. Although pain ratings appeared to decline over time, none of the changes reached statistical significance. Scores decreased by −27.7% from day 23 to 44 (adjusted *p* = .496) and by −81.4% from day 23 to 82 (adjusted *p* = .295). Similarly, no significant change was observed between days 44 and 82 (−74.3%, adjusted *p* = .496).

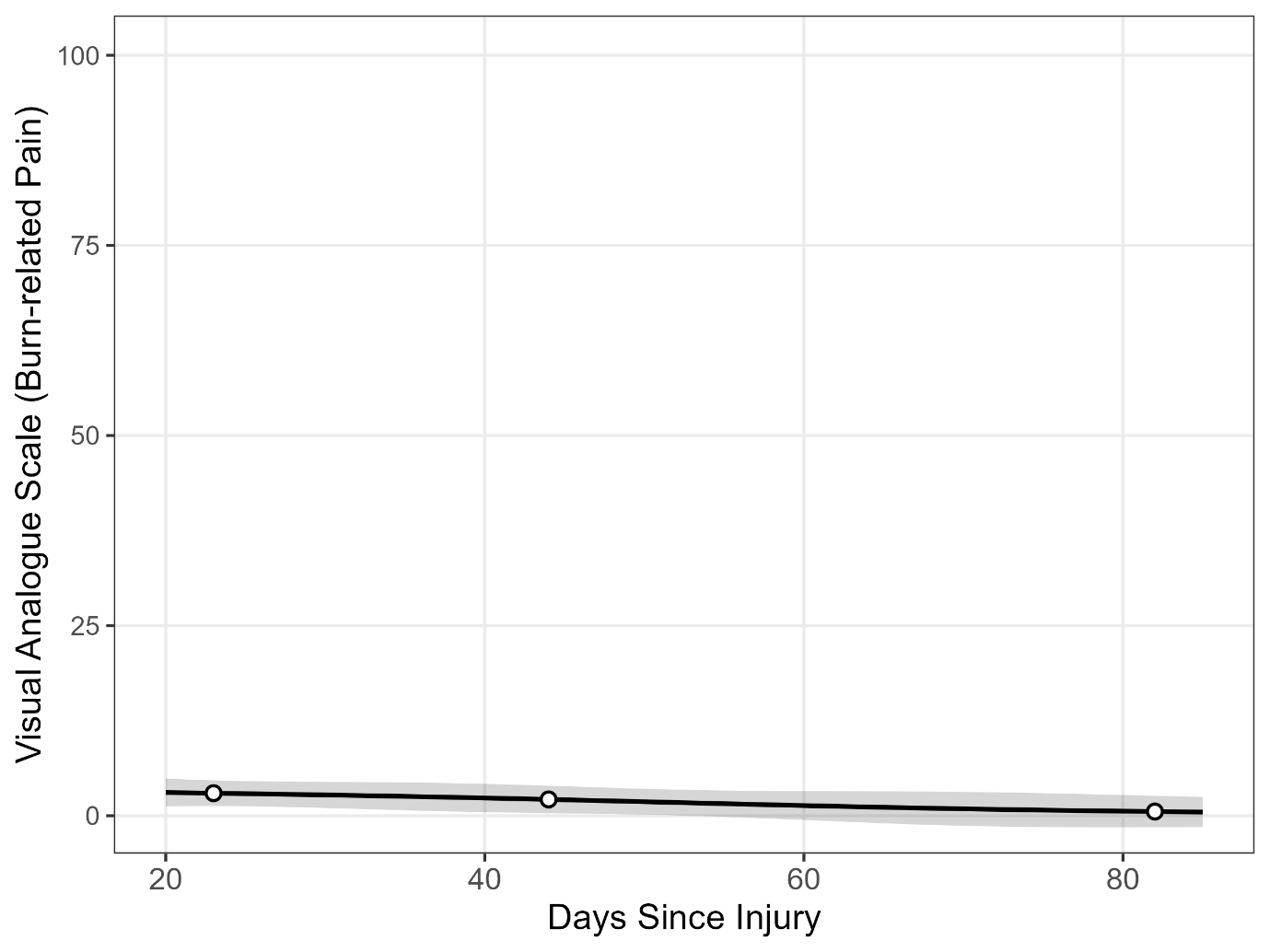
Figure S4.6. Perceived burn-related pain ratings across recovery. Model-predicted Visual Analogue Scale scores (black line ± 95% CI: shaded area) are shown between 20 and 85 days post-injury. White circles indicate estimated means at 23, 44, and 82 days. Visual Analogue Scale scores remained low and stable over time, with no significant differences between timepoints (all adjusted *ps* ≥ .295).

***S4.7 Semmes-Weinstein Monofilament Test***

Methods. A 6-piece monofilament kit (Touch-Test Sensory Monofilaments, North Coast Medical, USA) was used to examine touch sensitivity (6). The filament was applied at a 90° angle to the skin surface for 1 s with sufficient force to bend the filament. Participants were instructed to close their eyes and verbally report whether they felt the filament touch their skin. A successful detection of the filament was determined when participants responded with a ‘yes’ when the filament was bending. The full range of filament sizes from the kit were 2.83 – 6.65 grams. A thick filament was initially applied to familiarise participants with the sensation. The starting filament strength differed across participants due to the burn injury location (starting filament ranged from 4.31 – 6.65 grams). The filament strength was reduced when participants successfully detected 2 out of 3 stimuli. The test score reflects the finest filament the participants could successfully detect.

Results. Model-predicted monofilament detection thresholds (±95% CI) are shown across recovery for the most-affected (A–B) and least-affected (C–D) sides, measured above and below the injury site (Figure S4.7). A total of 25–27 participants contributed 63–70 observations per monofilament site. Testing was not performed in some participants when clinical dressings covered the appropriate sites or when the burn injury was located near sensitive regions such as the genital, chest, or facial areas. Monofilament detection thresholds at proximal and distal sites on both the most- and least-affected sides were modelled across 23, 44, and 82 days post-injury. Pairwise comparisons confirmed no significant changes over time: changes from day 23 to 44 ranged from −1.5% to +4.4% (all adjusted *p* ≥ .358), and from day 23 to 82 ranged from +0.3% to +2.9% (all adjusted *p* ≥ .953). Similarly, changes between days 44 and 82 ranged from −1.3% to +2.0% (all adjusted *p* = 1.000).

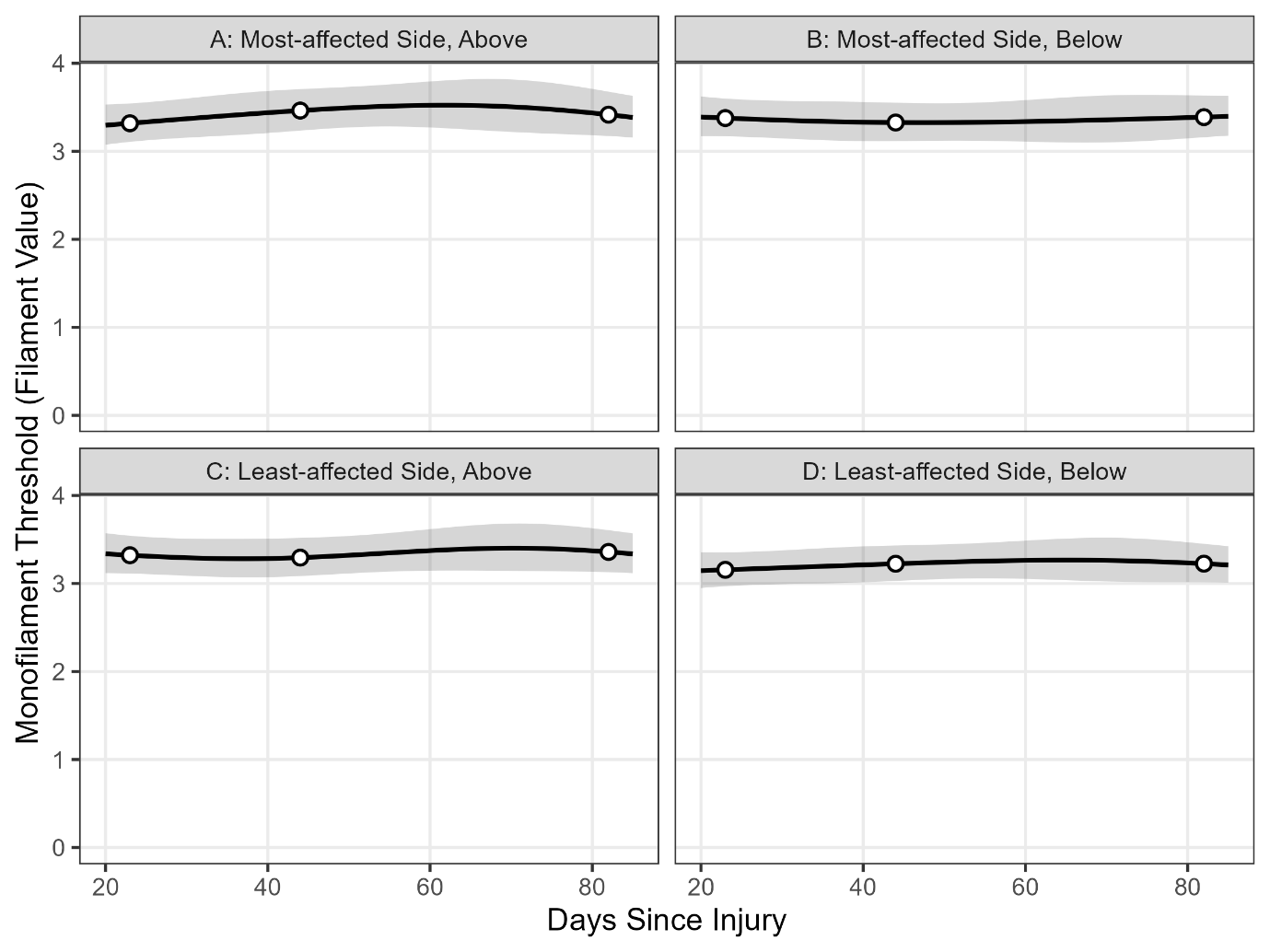

Figure S4.7. Monofilament detection thresholds across recovery. Model-predicted monofilament thresholds (black lines ± 95% CI: shaded area) are shown for the most-affected side (A–B) and least-affected side (C–D), measured above (left) and below (right) the injury site. White circles indicate estimated marginal means at 23, 44, and 82 days. Thresholds were stable across all sites and timepoints, with no significant differences observed (all adjusted *ps* ≥ .358).

***S4.8 Brush Touch Perception***

Methods. A lightweight paintbrush was used to assess touch perception. The sensory testing sites were brushed lightly with a backward and forward stroke, with participants asked to describe the brush sensations using one of these categorisations: normal (like felt during a familiarisation trial on unaffected skin on the least-affected side), painful (allodynia), stronger than expected but not painful (hyperaesthesia), weaker than expected (hypoaesthesia), or abnormal (paraesthesia).

Results. Because brush scores are descriptive categories rather than continuous measures, analysis focused on the distribution of responses rather than model-based score trajectories. A total of 19–25 observations were available per site in each time window. Brush responses were predominantly rated as normal at all sites and timepoints. In the early phase (0–30 days), 86–95% of responses were normal, with the remainder classified as weaker or stronger than expected (e.g., proximal, most-affected site: 86% normal, 5% weaker, 10% stronger). By 31–60 days, 92–100% of responses were normal, and by >60 days this increased further to 92–100% across all sites. Overall, non-normal responses were infrequent, occurred mainly in the first month post-injury, and were largely absent beyond 60 days.

***S4.9 Pressure Pain Threshold***

Methods. An algometer (FDX, Wagner Instruments, Greenwich, CT) with an 8-mm diameter hemispheric rubber tip was applied at the sensory testing locations as well as three standardised sites: (1) forehead (including a familiarisation trial at the start); (2) back of the hand to the most-affected side; and (3) back of the hand to the least-affected side (7). The pressure was increased at a constant rate of 1 kg/s until the participant reported that the pressure changed to pain. The test score reflects the force (kg) required to generate the onset of pain.

Results. Model-predicted pressure pain thresholds (±95% CI) are shown across recovery for the most-affected side (A), the least-affected side of the back of the hand (B), and the forehead (C) between 20 and 85 days post-injury (Figure S4.9). A total of 24–30 participants contributed 60–75 observations per pressure-pain site. Similar to the monofilament observations, data were not collected from all participants due to clinical dressings covering appropriate testing sites and when burn injury sites were located close to sensitive areas such as the genitals, chest, or face. Pressure pain ratings were modelled at 23, 44, and 82 days post-injury across proximal and distal sites on both the most- and least-affected sides as well as standardised locations on the back of the hands and forehead. Pairwise contrasts showed no statistically significant changes at any comparison: from 23 to 44 days changes ranged −5.2% to +0.9% (all adjusted *p* ≥ .657), from 23 to 82 days −2.2% to +8.5% (all adjusted *p* ≥ .548), and from 44 to 82 days −1.1% to +13.8% (all adjusted *p* ≥ .406). Overall, the pressure pain outcomes did not demonstrate meaningful change over the first 12 weeks post-injury.

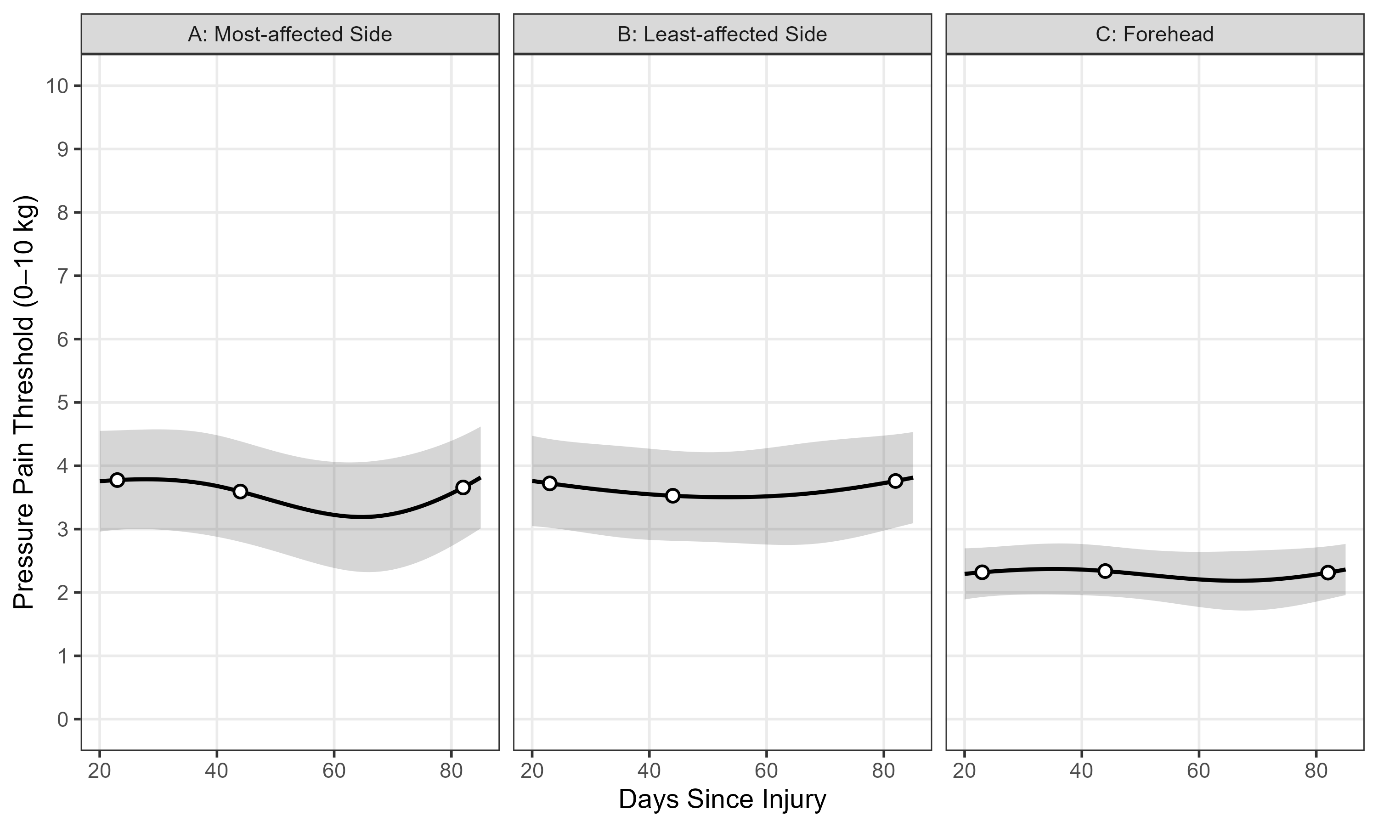

Figure S4.9. Pressure pain thresholds across recovery. Model-predicted pressure pain thresholds (black lines ± 95% CI: shaded area) are shown for the most-affected side of the back of the hand (A), the least-affected side of the back of the hand (B), and the forehead (C) between 20 and 85 days post-injury. White circles indicate estimated marginal means at 23, 44, and 82 days. Across all sites, thresholds were relatively stable over time, with modest fluctuations and no clear evidence of systematic increases or decreases.

***S4.10 Neuropen Perception Testing***

Methods. A dual-test peripheral neuropathy device (Neuropen, Owen Mumford, UK) was used to assess perceived sharp sensation using a 10 g von Frey monofilament and 40 g calibrated spring Neurotip (8). The filament was applied at a 90° angle to the skin for 1 s with sufficient force to bend the filament. The Neurotip was applied at a 90° angle to the skin for 2 s. Participants rated the sharpness of both the filament and Neurotip in two conditions: (1) a single application of the stimulus; (2) following five repeated applications of the stimulus administered 1 second apart. Sharpness was rated using a verbal rating scale ranging from 0 (not sharp) to 10 (extremely sharp).

Results. Figure S4.10 highlights the model-predicted Neuropen sharp (5) scores (±95% CI) across recovery for the most-affected (A–B) and least-affected (C–D) sides, measured above (left) and below (right) the injury site between 20 and 85 days post-injury. A total of 25 participants contributed 64–65 observations per dull and sharp Neuropen test site. Thresholds were modelled across 23, 44, and 82 days post-injury for both dull and sharp perception at proximal and distal sites on the most- and least-affected sides. Pairwise contrasts revealed no statistically significant changes over time. For dull tests, changes from day 23 to 44 ranged from −46% to +60%, and from day 23 to 82 from −8% to +112% (all adjusted *p* ≥ .502). For sharp tests, changes were smaller, ranging from −3% to −14% between days 23 and 44, and from −8% to −20% between days 23 and 82 (all adjusted *p* ≥ .391). Similarly, no significant differences were observed between days 44 and 82 (all adjusted *p* = 1.000). Overall, both dull and sharp Neuropen scores remained stable during the 12 weeks post-injury.

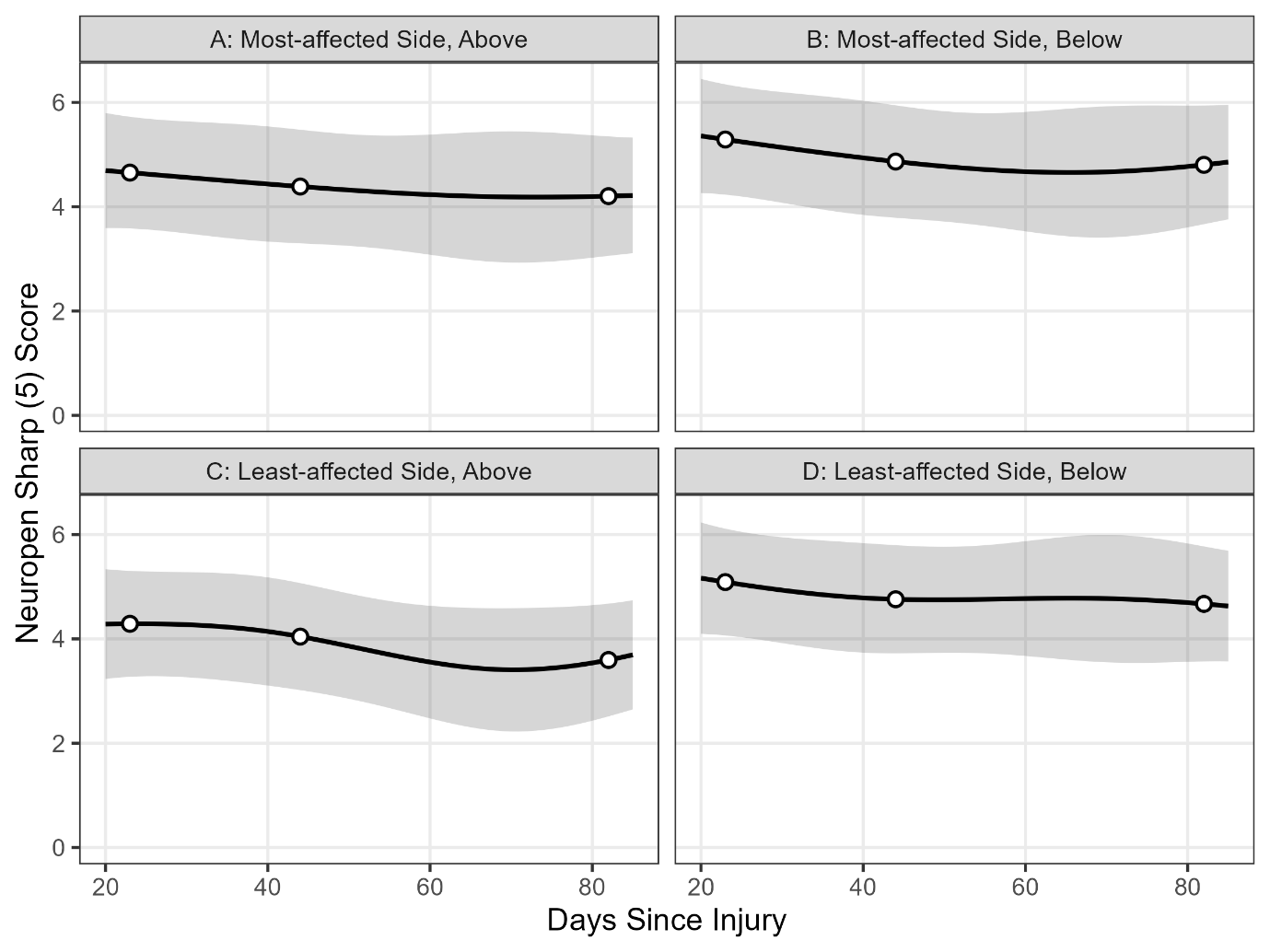

Figure S4.10. Neuropen sharp (5) scores across recovery. Model-predicted Neuropen sharp (5) scores (black lines ± 95% CI: shaded area) are shown for the most-affected side (A–B) and least-affected side (C–D), measured above (left panels) and below (right panels) the injury site between 20 and 85 days post-injury. White circles indicate estimated marginal means at 23, 44, and 82 days. Scores remained stable across all sites and timepoints, with no significant changes detected (all adjusted *p* = .391).

***S4.11 Grip Strength***

Methods. A handgrip force dynamometer (TTM Advanced Hand Dynamometer, Japan) was used to assess grip strength of both hands (9). Participants were instructed to sit in a chair and position their elbow at 90º with their elbow underneath their shoulder. The participants were advised to squeeze the dynamometer trigger ‘as fast and hard as possible’ for 3 s without changing their starting position. The order of hand testing was randomised, with each hand tested alternately in three trials. The best result (in kg) from each hand was then analysed.

Results. Figure S4.11 shows the model-predicted grip strength (±95% CI) across recovery for the most-affected side (A) and least-affected side (B), with estimates highlighted at 23, 44, and 82 days post-injury. A total of 34 participants contributed 90 observations per grip site (most- and least-affected hands). Grip strength scores were modelled across 23, 44, and 82 days post-injury. Pairwise contrasts showed small increases over time (e.g., +3.3% from day 23 to 82), but none reached statistical significance (all adjusted *p* ≥ .125). Overall, grip strength remained consistent across the 12-week follow-up period.

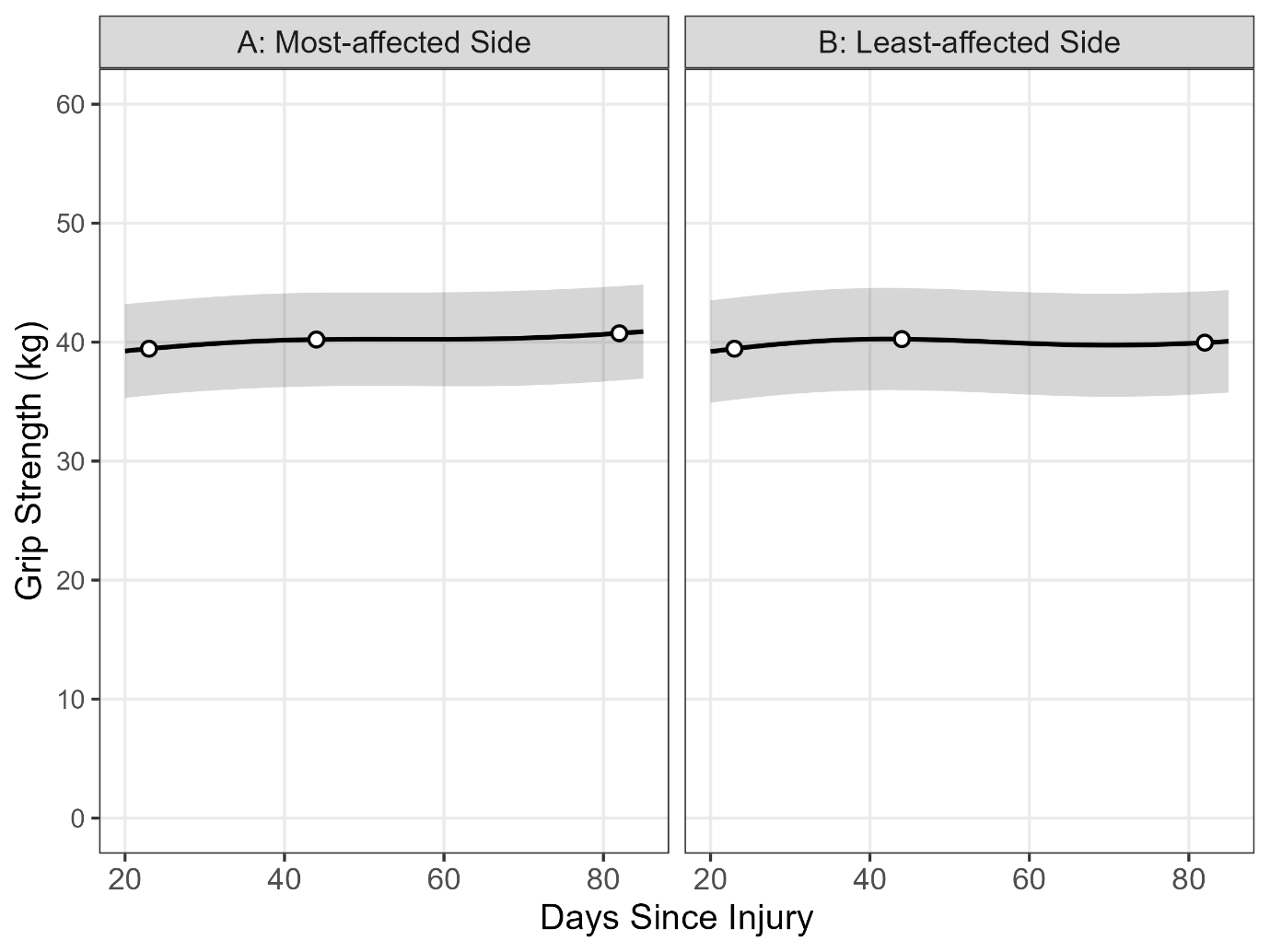

Figure S4.11. Model-predicted grip strength across recovery. Estimated marginal means (solid lines ± 95% CI shaded area) are shown for the most-affected side (panel A) and least-affected side (panel B) at 23, 44, and 82 days post-injury. Open circles highlight model estimates at those timepoints. Grip strength remained stable over time on both sides, with no statistically significant changes observed between 23, 44, and 82 days post-injury.

***S5. COMPARING SUBACUTE BURN PATIENTS TO NON INJURED CONTROLS***

***S5.1 Data Analysis***

Burns participants were included only if their assessments fell within prespecified post-injury windows (3 weeks: days 14–28; 6 weeks: days 35–49; 12 weeks: days 77–91). Controls completed a single assessment. The analytic sample comprised 31 burns participants and 35 controls. Attendance within the testing windows for burn participants was heterogeneous. Eight were captured across all three windows, five at both 3 and 6 weeks, eight at both 6 and 12 weeks, and one at 3 and 12 weeks. An additional nine participants were captured at a single window only (three each at 3, 6, and 12 weeks).

We analysed each Purdue Pegboard subtest separately using ordinary least squares (OLS) regression with fixed effects for GROUP (burns vs. control), TIMEPOINT (3, 6, and 12 weeks post-injury), and their interaction. We then used the model to estimate adjusted means for each group and timepoint and compared the burns group at 3, 6, and 12 weeks with the control group. For inference, we used a nonparametric, subject-level (cluster) bootstrap: participants were resampled with replacement 1,000 times, the same OLS model was refit to each bootstrap sample, and the contrasts were recomputed. Percentile 95% bootstrap confidence intervals were the decision criterion (intervals excluding zero were considered significant).

***S5.2 Pegboard Performance in Subacute Burn Patients and Controls***

Pegboard performance across recovery for controls and burns participants: dominant hand (A), non-dominant hand (B), and simple bilateral test (C) (Figure S5.2). Figure S5.2.1 highlights the assembly pegboard performance across recovery in controls and burns participants. For the dominant hand, the burns group scored significantly lower than controls at 3 weeks post-injury (−1.65; bootstrap 95% CI [−2.87, −0.49]. Differences at 6- and 12-weeks post-injury were not significant (6 weeks: −0.82; CI [−1.86, 0.20]; 12 weeks: −0.04; CI [−0.97, 0.95]), pointing to recovery to near-control levels. For the non-dominant hand, the between-group differences were modest and not significant at any timepoint: 3 weeks: −0.89; CI [−1.89, 0.09]; 6 weeks: −0.76; CI [−1.68, 0.11]; 12 weeks: −0.33; CI [−1.38, 0.69]. Similarly, for the simple bilateral subtest, all contrasts were non-significant: 3 weeks: −0.54; CI [−1.31, 0.23]; 6 weeks: −0.30; CI [−1.07, 0.49]; 12 weeks: −0.71; CI [−1.55, 0.15]. For the assembly subtest, the burns group scored significantly lower than controls group at 6 weeks post-injury: −3.40; CI [−6.49, −0.20]. However, at 3 weeks and 12 weeks post-injury, there were no significant between-group differences (3 weeks: −3.57; CI [−7.54, 0.62]; 12 weeks: −2.68; CI [−6.07, 0.68]).

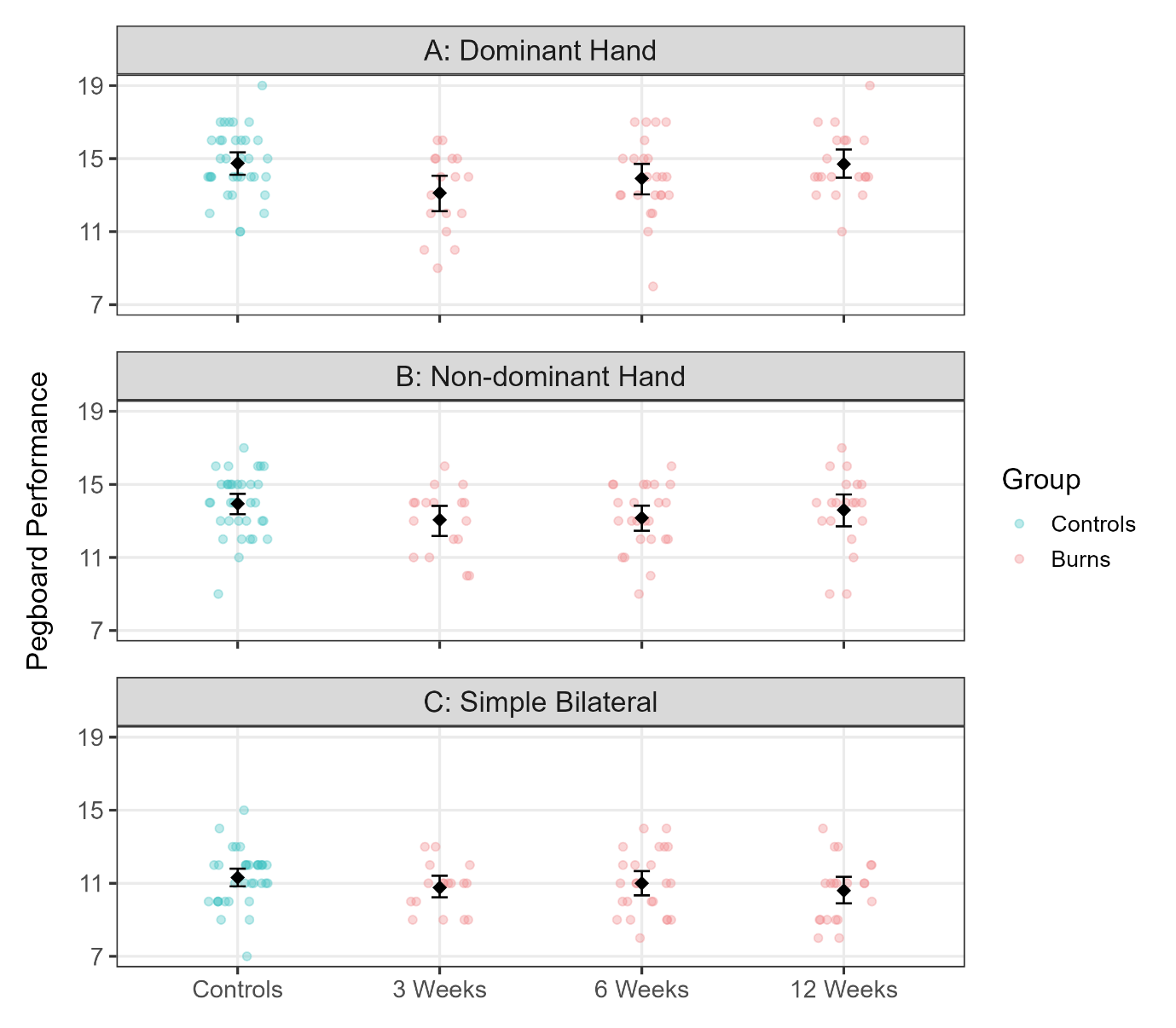

Figure S5.2. Pegboard performance in controls and burns participants across recovery for the dominant hand (A), non-dominant hand (B), and simple bilateral test (C). Individual scores are shown with group means ± 95% bootstrap confidence intervals (black diamonds and bars). The burns group performed significantly worse than controls at 3 weeks post-injury on the dominant hand (−1.65; bootstrap 95% CI [−2.87, −0.49]), whereas differences at 6 weeks and 12 weeks were not significant. For the non-dominant and simple bilateral tests, between-group contrasts at 3, 6, and 12 weeks were modest and not statistically significant, indicating recovery toward control levels.

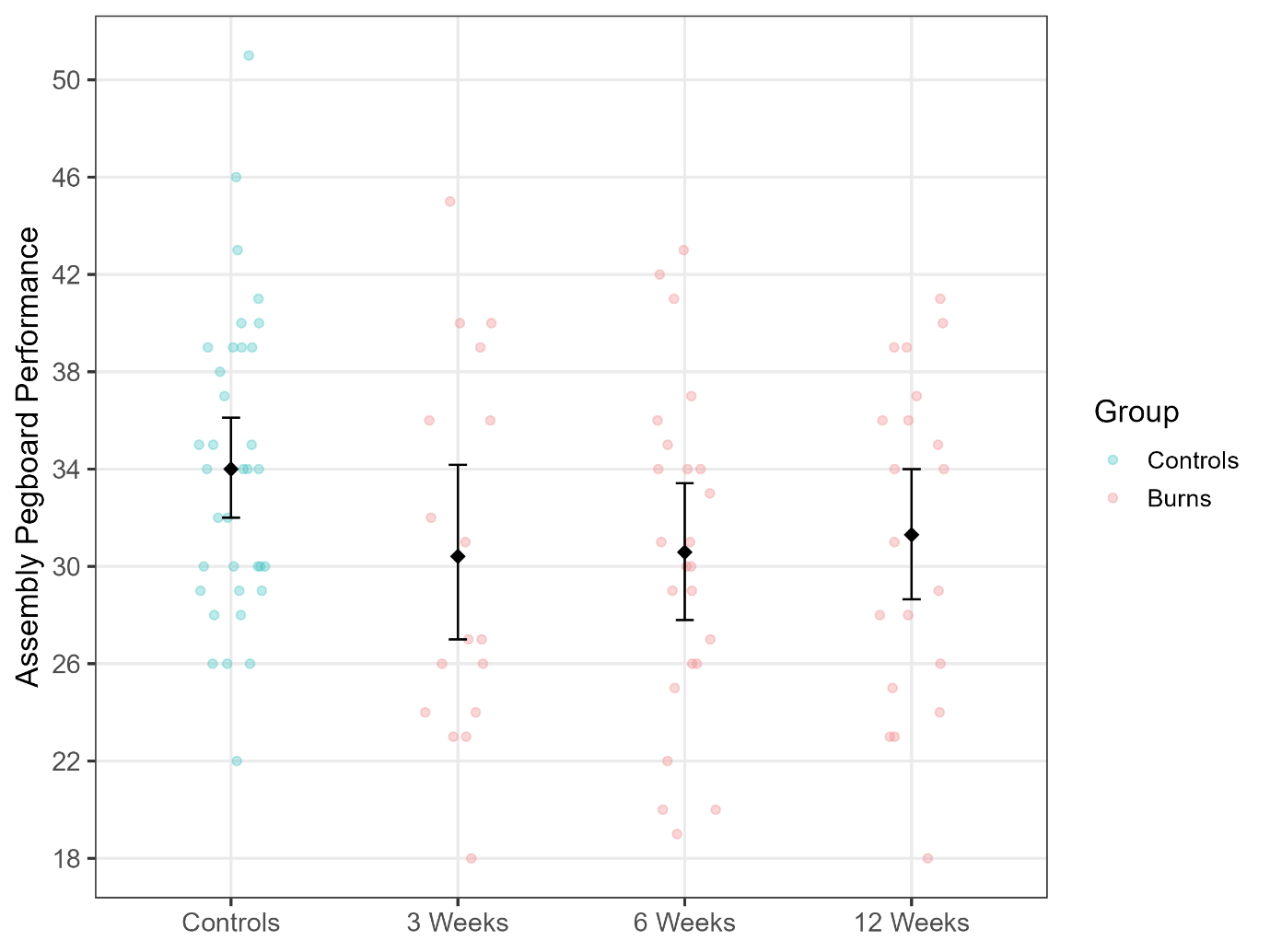

Figure S5.2.1. Assembly pegboard performance in controls and burns participants across recovery. Individual scores are shown with group means ± 95% bootstrap confidence intervals (black diamonds and bars). The burns group scored significantly lower than controls at 6 weeks post-injury (−3.40; bootstrap 95% CI [−6.49, −0.20]). At 3 weeks and 12 weeks, differences were not statistically significant.

***S5.3 SF36 Physical and Mental Component Scores in Subacute Burn Patients and Controls***

Figure S5.3 shows the SF36 summary scores in controls and burns participants across recovery for the Physical Component Summary (A) and Mental Component Summary (B). For Physical Component Summary, the burns group scored significantly lower than the control group at 3 weeks post-injury: −5.97; CI [−11.07, −0.80]. However, at 6 weeks and 12 weeks post-injury there were no significant between-group differences (6 weeks: −0.07; CI [−3.00, 2.78]; 12 weeks: +1.33; CI [−1.53, 3.77]). Similarly, for Mental Component Summary, the burns group scored significantly lower than the control group at 3 weeks post-injury: −4.45; CI [−8.52, −0.57]. However, at 6 weeks and 12 weeks post-injury there were no significant between-group differences (6 weeks: −3.12; CI [−7.17, 0.81]; 12 weeks: −2.39; CI [−6.59, 1.51]).

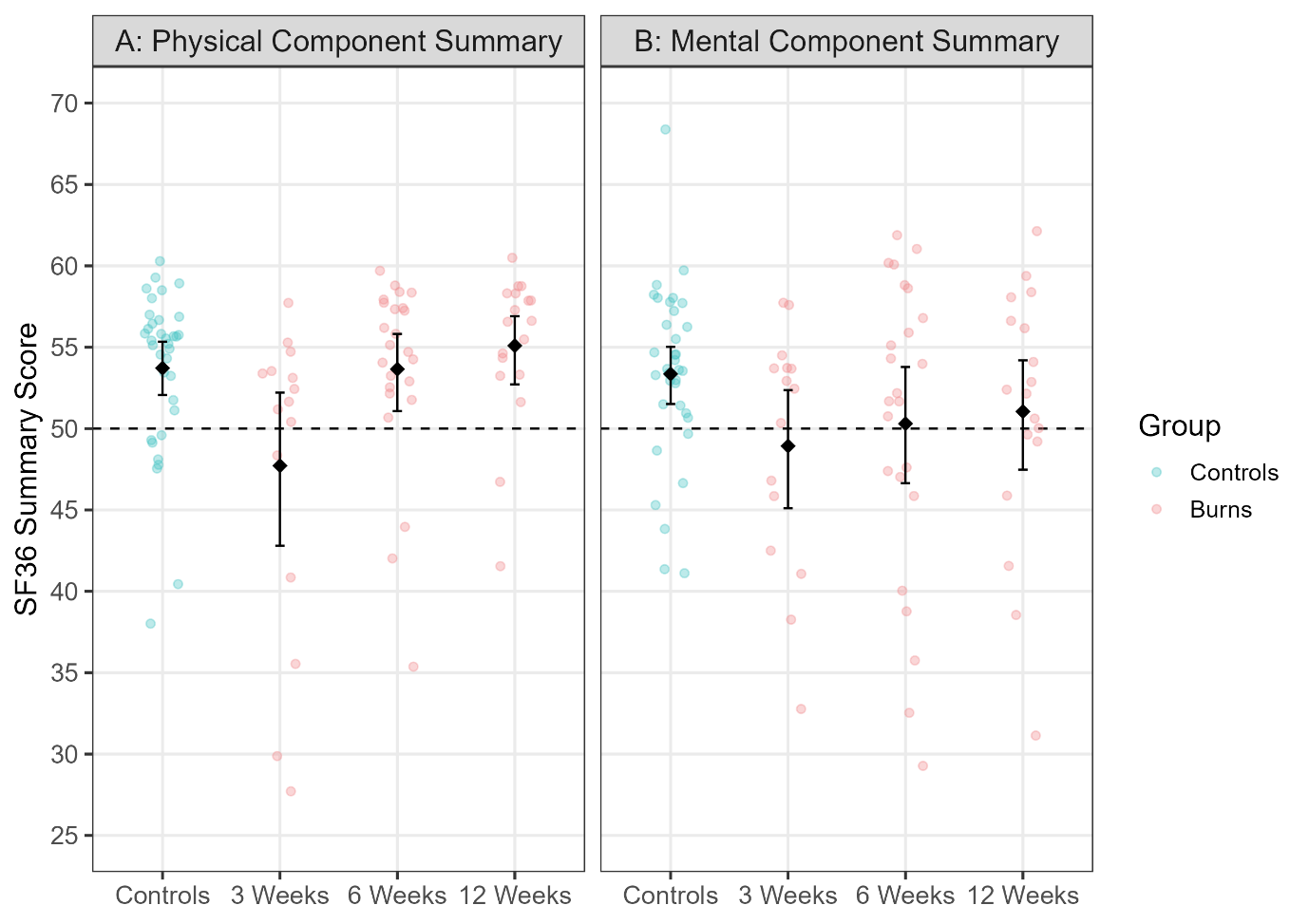

Figure S5.3. SF36 summary scores in controls and burns participants across recovery for the Physical Component Summary (A) and Mental Component Summary (B). Individual scores are shown with group means ± 95% bootstrap confidence intervals (black diamonds and bars). The burns group scored significantly lower than controls at 3 weeks post-injury for both components (Physical Component Summary: −5.97, bootstrap 95% CI [−11.07, −0.80]; Mental Component Summary: −4.45, bootstrap 95% CI [−8.52, −0.57]). At 6 and 12 weeks, between-group differences were not statistically significant.

***S5.4 Baseline Intracortical Inhibition in Subacute Burn Patients and Controls***

Figure S5.4 shows the baseline corticospinal excitability and intracortical inhibition in controls and burns participants at PRE (3, 6, and 12 weeks post-injury). At baseline (PRE), single-pulse amplitude was not different between the burns group and controls (all adjusted *p* ≥ .133). Baseline SICI was greater for controls than burns patients at 3 and 6 weeks post-injury (adjusted *p* = .048, .045) but not 12 weeks post-injury (adjusted *p* = .547). Baseline LICI was greater for controls than burns patients at 6 week post-injury (adjusted *p* = .039) but not 3 or 12 weeks post-injury (adjusted *p* = .420, .120). Overall, baseline (PRE) data suggest reduced intracortical inhibition in the burns group compared with controls early in recovery, particularly for SICI at 3–6 weeks and LICI at 6 weeks, with these differences diminishing by 12 weeks.

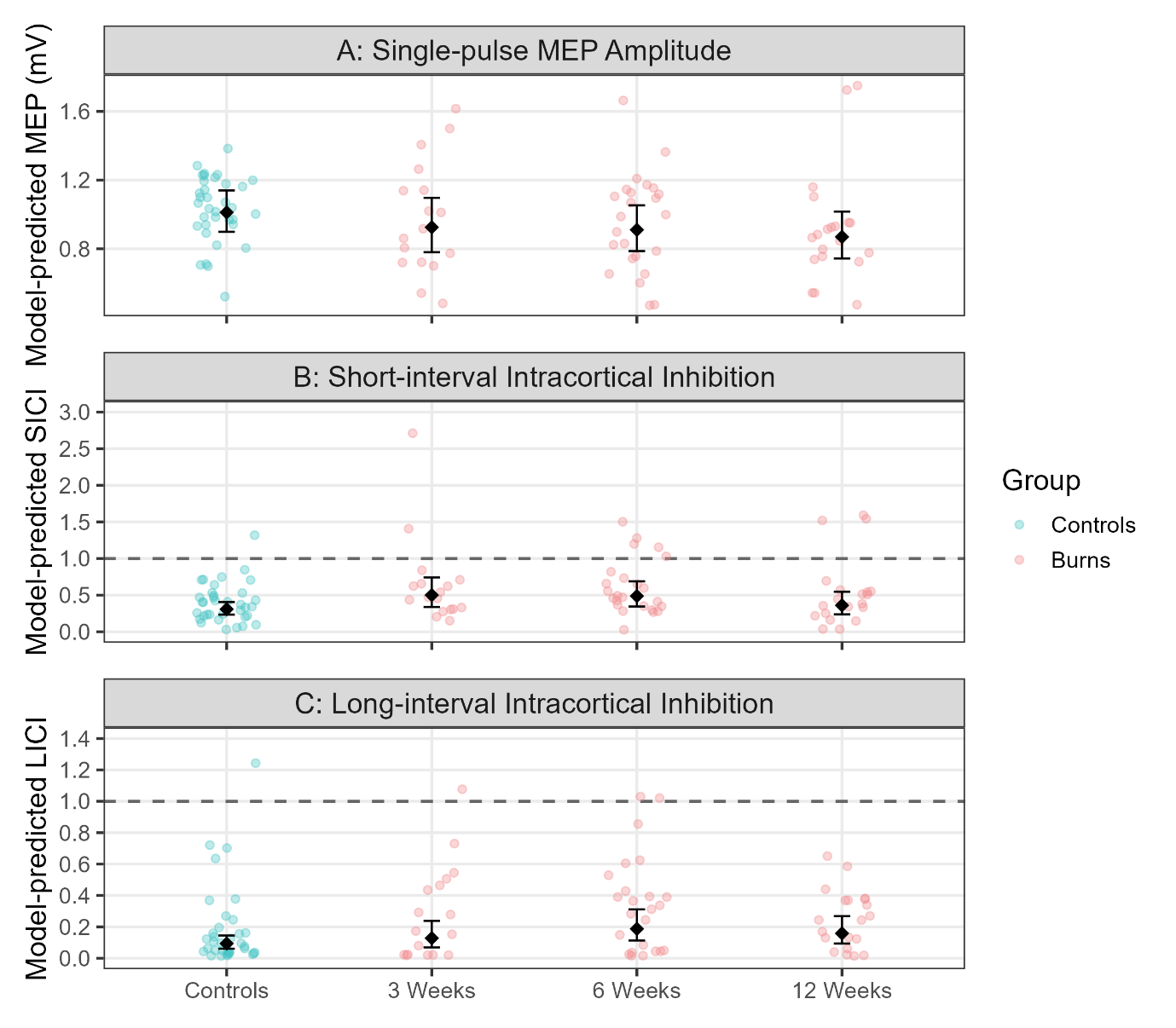

Figure S5.4. Baseline corticospinal excitability and intracortical inhibition in burn patients and controls. Model-predicted values at PRE are shown for controls and for burns participants at 3, 6, and 12 weeks post-injury. (A) Single-pulse MEP amplitude: Dots represent subject-level predicted values, with black diamonds and error bars indicating estimated marginal means (±95% CI). Baseline single-pulse amplitudes were 9–14% lower in burns versus controls, but group differences were not statistically significant. (B) Short-interval intracortical inhibition (SICI): Ratios < 1.0 indicate inhibition relative to single-pulse responses (dashed line = reference). Burns participants showed consistently higher SICI ratios (reduced inhibition) compared with controls, with significant differences at 3 and 6 weeks (adjusted *p* = .048 and .045, respectively). By 12 weeks, group differences had diminished and were not significant. (C) Long-interval intracortical inhibition (LICI). Burns participants demonstrated higher LICI ratios (reduced inhibition) than controls, with the largest difference observed at 6 weeks (adjusted *p* = .039). Differences at 3 and 12 weeks were not significant. These results suggest transient reductions in both GABA_A_- and GABA_B_-mediated inhibition following burn injury, most evident in the early recovery phase.

***S5.5 PAS-Induced Neuroplasticity in Recovery in Subacute Burn Patients and Controls***

Figures S5.5.1–S5.5.3 highlight model-predicted changes during PAS-induced neuroplasticity: (1) single-pulse MEP amplitudes (S5.5.1), (2) SICI ratios (S5.5.2), and (3) LICI ratios (S5.5.3). Data are shown for controls (top panels) and burn patients (bottom panels) across baseline (PRE) and three PAS blocks (0, 15, and 30 min post-PAS). Analyses focused on single-pulse MEP amplitude showed no reliable between-group differences in PAS-related change from PRE at 3 or 6 weeks (changes ranged from +3% to +35%; all adjusted *p* ≥ .071). At 12 weeks, controls exhibited a significantly greater increase at 15-min post-PAS (+45%, adjusted *p* = .036), while contrasts at POST-PAS and 30-min were not significant (changes ranged from +7% to +22%; adjusted *p* ≥ .245). Overall, PAS-induced changes in single-pulse MEP amplitude were largely comparable between groups, with only a transient difference detected at 12 weeks.

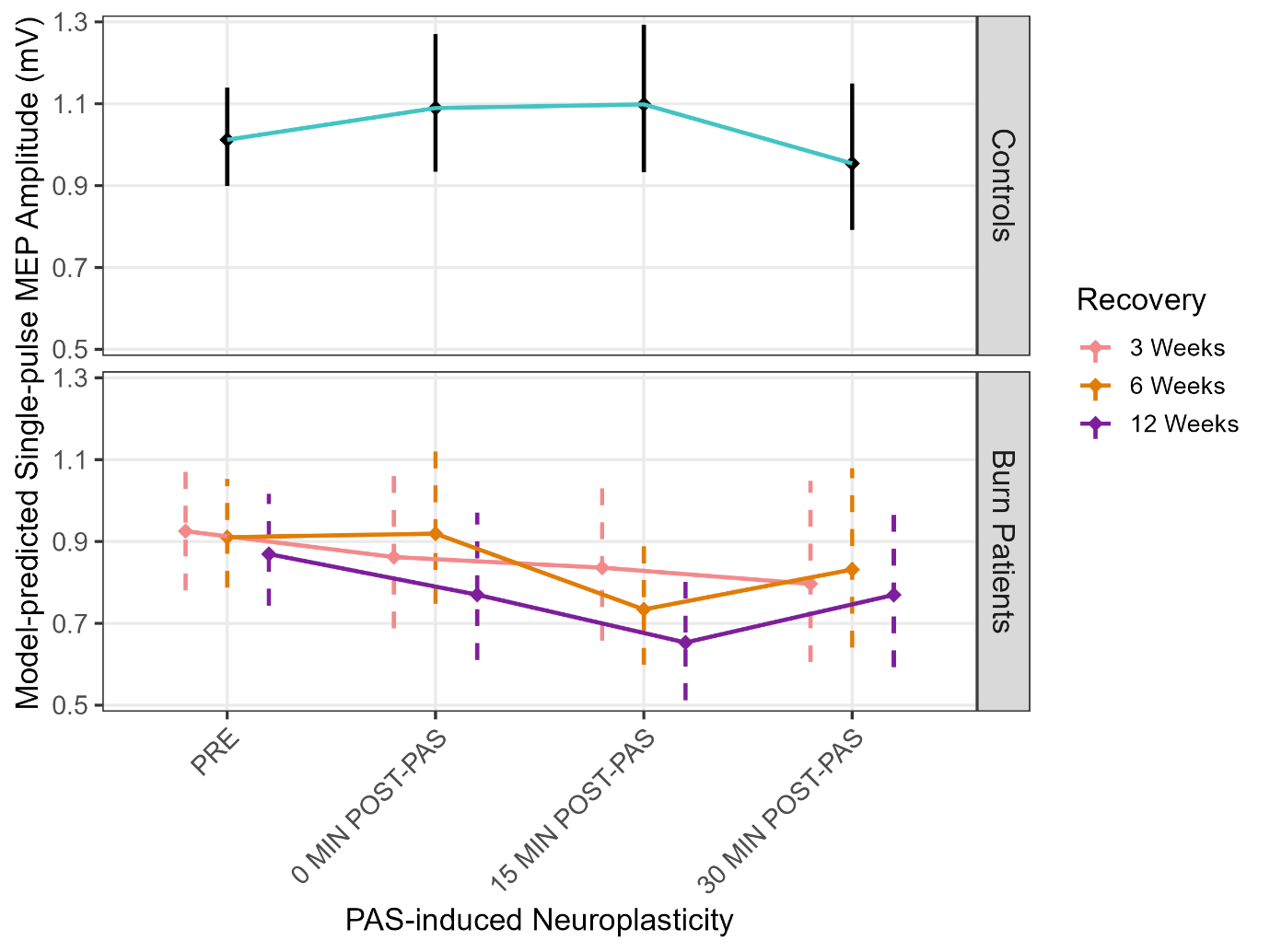

Figure S5.5.1. Model-predicted single-pulse MEP amplitudes during PAS-induced neuroplasticity. Data are shown for Controls (top panel) and Burn Patients (bottom panel) across baseline (PRE) and three PAS blocks (0, 15, and 30 min post-PAS). Diamonds denote group-level model-predicted means, with error bars representing 95% confidence intervals (CIs). For Controls, CIs are shown as solid black lines; for Burn Patients, CIs are displayed as dashed lines coloured by recovery stage (pink = 3 weeks, orange = 6 weeks, purple = 12 weeks). Lines connect model-predicted means for visual continuity. Statistical contrasts were performed relative to PRE. No reliable between-group differences were observed at 3 or 6 weeks (all adjusted *p* ≥ .071). At 12 weeks, controls showed a significantly greater increase at 15 min post-PAS (+45%, adjusted *p* = .036), though contrasts at 0 and 30 min were not significant. Overall, PAS-related changes were largely comparable between groups, with only a transient divergence at 12 weeks.

At 3 weeks post-injury, the change in SICI ratios from baseline to immediately post-PAS (but not the change from baseline to 15- or 30-minutes post-PAS) differed between burn patients and controls, with SICI decreasing post-PAS in controls compared with the burns group (-38%, adjusted *p* = .035). At 6 and 12 weeks post-injury, there were no statistical differences in SICI ratios across blocks between burn patients and controls (changes ranged from −10% to +18%; adjusted *p* ≥ .866).

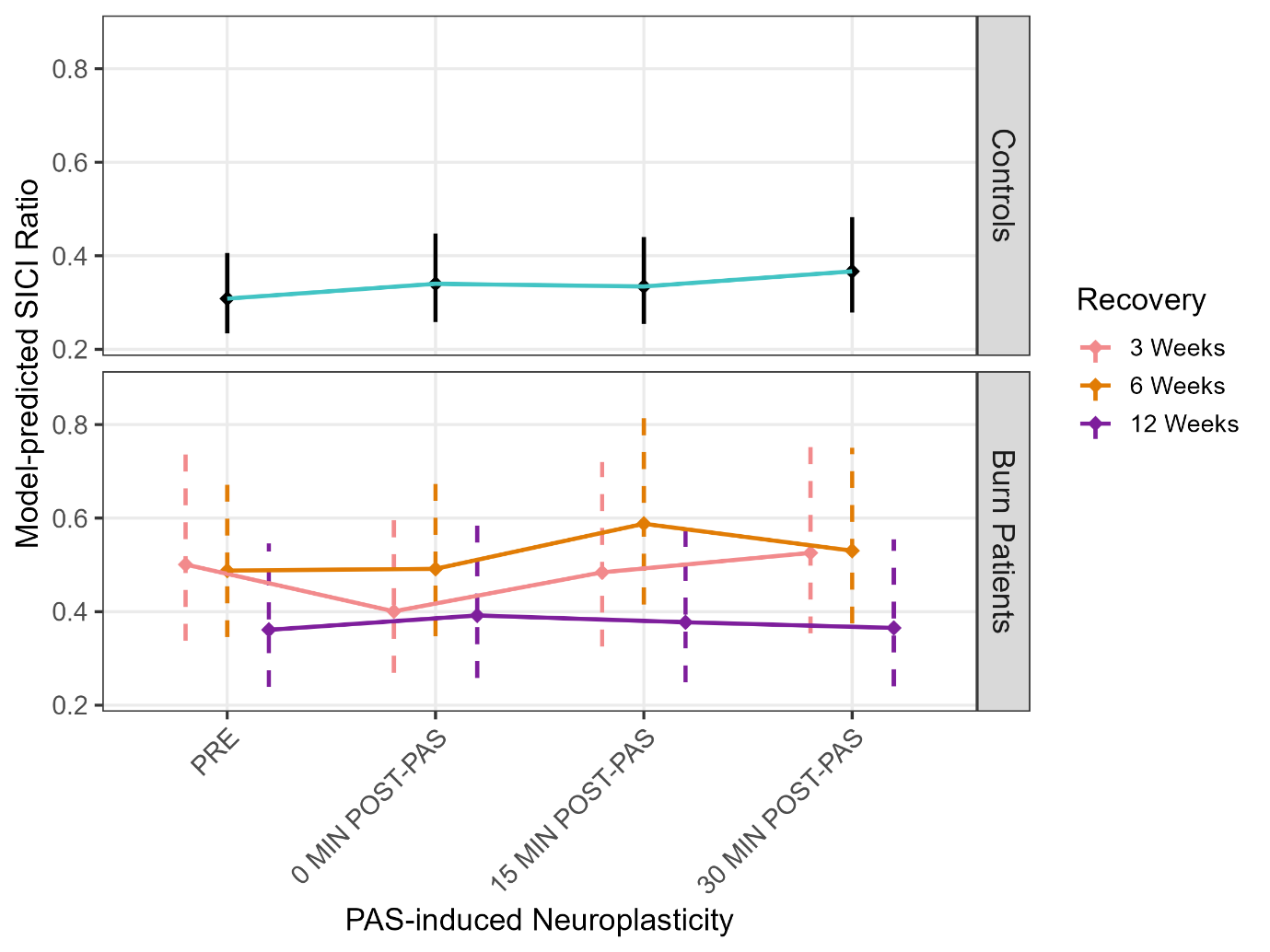
Figure S5.5.2. Model-predicted short-interval intracortical inhibition (SICI) ratios during PAS-induced neuroplasticity. Data are shown for Controls (top panel) and Burn Patients (bottom panel) across baseline (PRE) and three post-PAS blocks (0, 15, and 30 min). Diamonds denote group-level model-predicted means, with error bars representing 95% confidence intervals (CIs). For Controls, CIs are shown as solid black lines; for Burn Patients, CIs are displayed as dashed lines coloured by recovery stage (pink = 3 weeks, orange = 6 weeks, purple = 12 weeks). Lines connect model-predicted means for visual continuity. At 3 weeks post-injury, the change in SICI ratios from baseline to immediately post-PAS differed between groups, with controls showing an increase in SICI ratios (reflecting reduced inhibition) compared with burns patients (+38%, adjusted *p* = .035). No group differences were observed from baseline to 15 or 30 min post-PAS. At 6 and 12 weeks post-injury, there were no statistical differences in SICI ratios across blocks between burn patients and controls (changes ranged from −10% to +18%; all adjusted *ps* ≥ .866).

At 3 weeks post-injury, between-group contrasts confirmed that the burns group exhibited significantly greater increases in LICI ratios than controls at 15 min (+42%, adjusted *p* < .001) and 30 min post-PAS (+34%, adjusted *p* = .004), while the smaller difference immediately post-PAS (+20%, adjusted *p* = .230) was not significant. At 6 weeks, there were no statistical differences in LICI ratios between controls and the burns group across blocks (changes ranged from −19% to +26%: adjusted *p* ≥ .232). At 12 weeks post-injury, between-group contrasts confirmed that burns exhibited a significantly greater increase in LICI ratios than controls at 15 min post-PAS (+27%, adjusted *p* = .049), while differences at POST-PAS (+7%) and 30 min (−13%) were not significant (adjusted *p* ≥ .936).

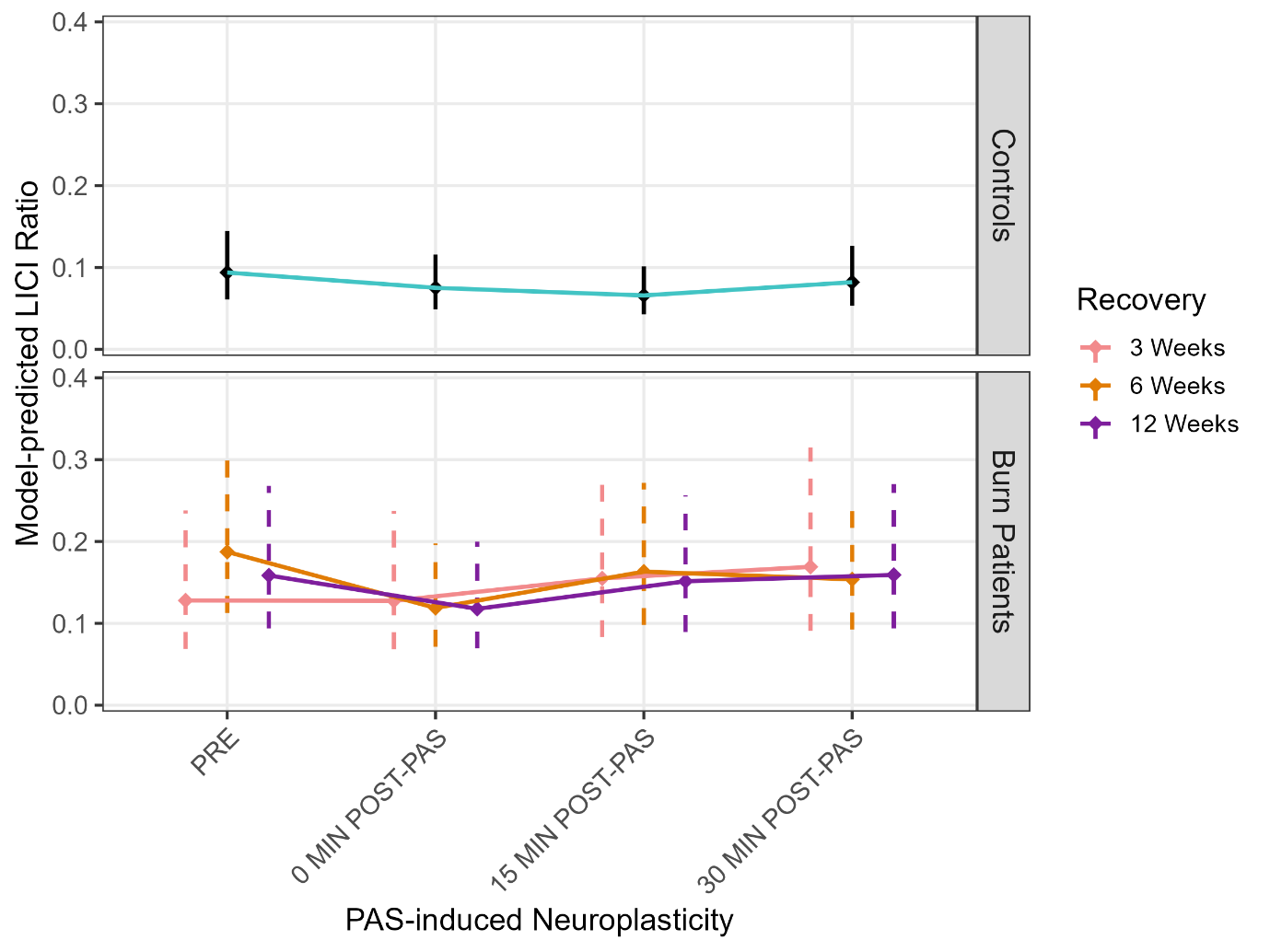

Figure S5.5.3. Model-predicted long-interval intracortical inhibition (LICI) ratios during PAS-induced neuroplasticity. Data are shown for Controls (top panel) and Burn Patients (bottom panel) across baseline (PRE) and three post-PAS blocks (0, 15, and 30 min). Diamonds denote group-level model-predicted means, with error bars representing 95% confidence intervals (CIs). For Controls, CIs are shown as solid black lines; for Burn Patients, CIs are displayed as dashed lines coloured by recovery stage (pink = 3 weeks, orange = 6 weeks, purple = 12 weeks). Lines connect model-predicted means for visual continuity. At 3 weeks post-injury, between-group contrasts confirmed that burns exhibited significantly greater increases than controls at 15 min (+42%, adjusted *p* < .001) and 30 min post-PAS (+34%, adjusted *p* = .004), while the smaller difference immediately post-PAS (+20%) was not significant (adjusted *p* = .230). At 6 weeks, no between-group differences were observed (changes ranged from −19% to +26%; all adjusted *p* ≥ .232). At 12 weeks, between-group contrasts confirmed a significantly greater increase in LICI in burns at 15 min post-PAS (+27%, adjusted *p* = .049), with differences at POST-PAS (+7%) and 30 min (−13%) not significant (all adjusted *p* ≥ .936).

***S6. ANALYSING SUBACUTE BURN PATIENTS WITH UPPER-LIMB BURN INJURIES***

Another GLMM model was fit to examine TMS outcomes across recovery trajectories in a subsample of participants with upper-limb burn injuries (n = 20). The model used a Gamma distribution with log link and included fixed effects for days since injury, PAS block, and stimulation condition, along with all interactions. A natural cubic spline with 3 degrees of freedom again captured non-linear effects of days since injury. The random-effects structure included participant-specific intercepts and uncorrelated slopes for days post-injury and stimulation condition. The model converged with a minor Hessian-related warning, but parameter estimates were stable. This targeted analysis allowed for more focused interpretation of baseline inhibition and PAS-induced neuroplasticity within a functionally relevant burn-injured subgroup.

***S6.1 Baseline SICI and LICI in Subacute Burn Patients with Upper-limb Injury***

Amplitude Differences

Amplitude ratios during the baseline (PRE) block were evaluated at 23, 44, and 82 days post-injury to track changes in intracortical inhibition during recovery. To ensure corticospinal excitability did not impact inhibitory ratio results, single-pulse amplitudes were evaluated and found to be consistent across timepoints, with model-estimated values of 0.93 (day 23), 1.05 (day 44), and 0.85 (day 82). Although these numerical differences reflect modest fluctuations, none of the pairwise contrasts were statistically significant (all adjusted *p* = 1.000). Similarly, baseline intracortical inhibition remained stable. LICI amplitude ratios were consistently low (0.13–0.11), and SICI ratios remained steady (0.41–0.40), with all between-day comparisons yielding minimal differences and adjusted *p* = 1.000. These findings suggest that baseline intracortical inhibition was unchanged throughout the subacute period for the subsample of patients who suffered an upper-limb burn injury.

Rate of Change Differences

Marginal slopes during the baseline (PRE) block were evaluated at 23-, 44-, and 82-days post-injury, with a focus on inhibition (LICI and SICI) relative to single-pulse trials, and on changes in corticospinal excitability to ensure inhibitory ratios were not confounded by shifts in baseline excitability. Slope estimates for single-pulse trials showed a transient reduction in corticospinal excitability, with a significant decrease from day 23 to 44 (−2.46%, adjusted *p* = .040). No significant changes were observed from day 23 to 82 (−1.58%, adjusted *p* = .188) or 44 to 82 (+0.90%, adjusted *p* = .234), suggesting limited recovery over time. In contrast, slope ratios for SICI and LICI (relative to single-pulse) remained stable across the subacute period. For SICI, estimated percent changes were −0.64% (23 to 44, adjusted *p* = 1.000), +0.31% (23 to 82, adjusted *p* = 1.000), and +0.95% (44 to 82, adjusted *p* = .795), indicating no significant change in SICI-induced inhibition. Similarly, LICI slope ratios showed no differences between timepoints: +0.77% (23 to 44, adjusted *p* = 1.000), −0.05% (23 to 82, adjusted *p* = 1.000), and −0.81% (44 to 82, adjusted *p* = 1.000). These findings suggest that while corticospinal excitability fluctuated early post-injury, intracortical inhibition remained stable for the subsample of patients with upper-limb burns.

***S6.2 Single-pulse MEP Amplitude and Intracortical Inhibition Following PAS in Upper-limb Injury Subsample***

Amplitude Differences

Amplitude ratios following PAS were evaluated at 23, 44, and 82 days post-injury to assess changes in intracortical inhibition and corticospinal excitability during recovery in individuals with upper-limb burn injuries. For single-pulse responses, model-estimated amplitudes declined across timepoints within each PAS block (e.g., –15% to –19% from day 23 to 82), but none of the between-day contrasts were statistically significant (all adjusted *p* = 1.000), indicating stable corticospinal excitability. No significant changes in intracortical inhibition were detected across days in any PAS block. LICI contrast differences ranged from –24% to +9% (all adjusted *p* ≥ .651), while SICI contrasts ranged from –29% to –2% (all adjusted *p* ≥ .292). These results indicate that the capacity for PAS-induced intracortical inhibition (LICI and SICI) was preserved throughout the subacute recovery period, with no systematic changes observed between 3 and 12 weeks post-injury.

Rate of Change Differences

Slope-based analyses were conducted at 23, 44, and 82 days post-injury to examine changes in PAS-induced intracortical inhibition and corticospinal excitability during subacute recovery. For single-pulse responses, model-estimated slopes declined modestly across timepoints within each PAS block (e.g., –2% to +2% from day 23 to 82), but none of the between-day contrasts were statistically significant (all adjusted *p* ≥ .432), indicating stable corticospinal excitability over time. Similarly, no significant changes in slope-based intracortical inhibition were observed across days in any PAS block. LICI slope contrast differences ranged from –2.2% to +1.8% (all adjusted *p* ≥ .243), while SICI slope contrast differences ranged from –3.1% to +2.1% (all adjusted *p* ≥ .209). These findings suggest that both PAS-induced excitability and inhibitory modulation remained stable from 3 to 12 weeks post-injury, with no evidence of delayed or progressive plasticity-related change during this recovery window.

***S6.3 Pegboard Performance in Individuals with Upper-limb Burn Injuries***

Pegboard Score Differences

Linear mixed models were used to examine changes in pegboard task performance at 23, 44, and 82 days post-injury in a subsample of 20 participants identified as having a burn located on the upper-limb (including upper arm, forearm, or hand). A significant improvement was observed for the dominant hand subtest between days 23 and 82, reflecting a +10.6% increase in performance (adjusted *p* = .020). No other comparisons reached statistical significance after Holm correction across the remaining subtests (non-dominant, simple bilateral, and assembly).

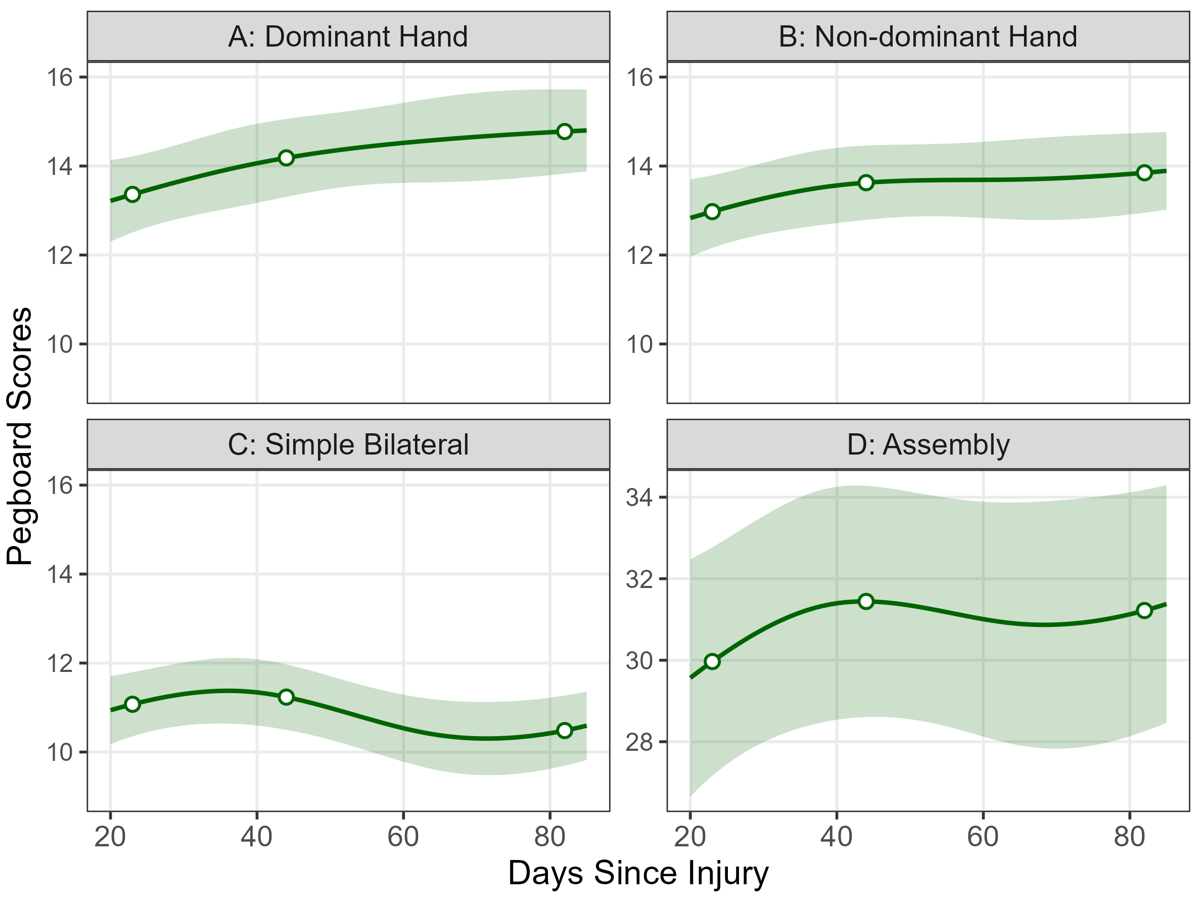

Figure S6.3. Modelled changes in pegboard performance across days 23, 44, and 82 post-injury in a subsample of burn survivors with arm injuries (n = 20). Estimated marginal means (lines) and 95% confidence intervals (shaded bands) are shown for the dominant hand (A), non-dominant hand (B), simple bilateral (C), and assembly (D) subtests.

Rate of Change Differences

Linear mixed models were used to examine changes in pegboard task performance at 23, 44, and 82 days post-injury in a subsample of 20 participants identified as having a burn located on the upper-limb (including upper arm, forearm, or hand). For the simple bilateral subtest, the rate of improvement significantly slowed between days 23 and 44 (–0.075 units/day, adjusted *p* = .038), before increasing again from day 44 to 82 (+0.067 units/day, adjusted *p* = .022). No significant changes in the rate of improvement were observed for the remaining subtests (dominant, non-dominant, and assembly; all adjusted *p* ≥ .520).

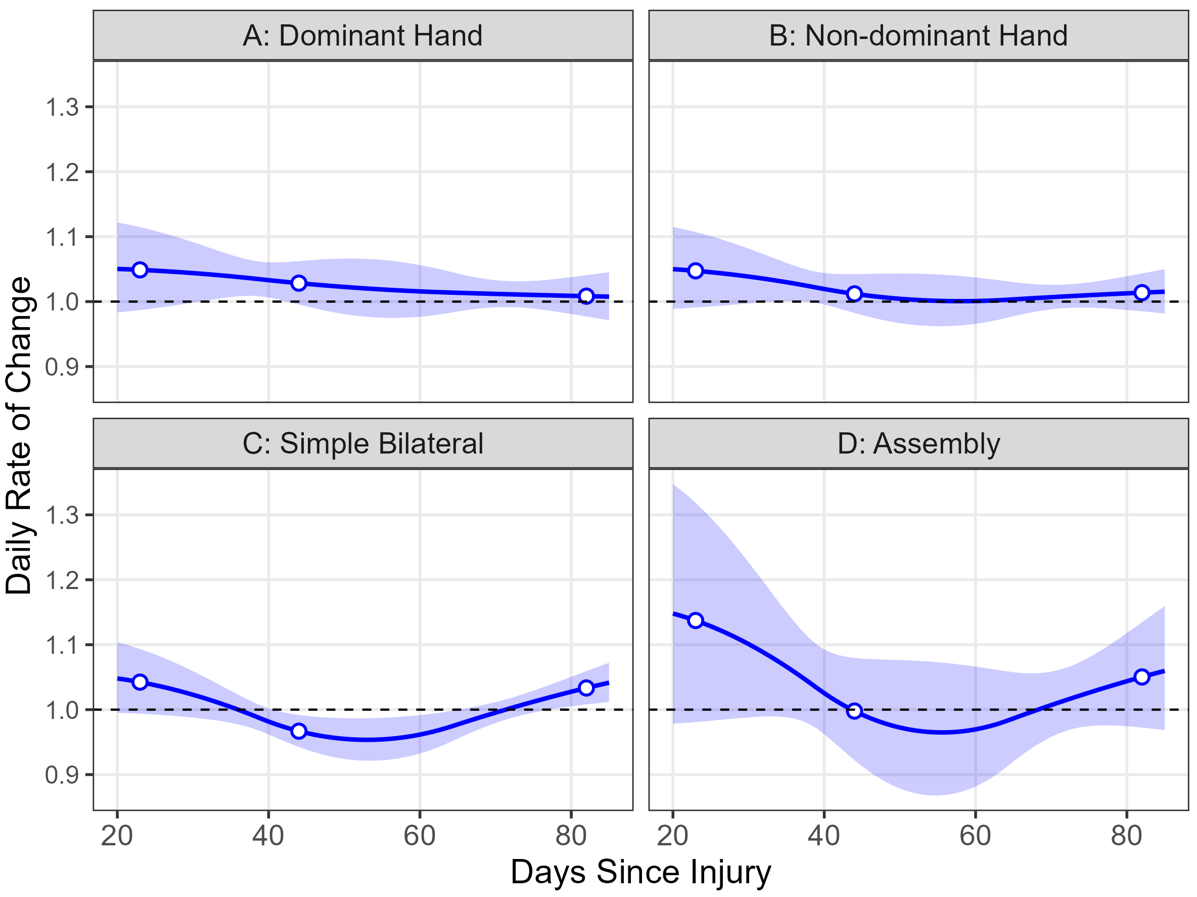

Figure S6.3.1. Estimated daily rate of change in pegboard performance at 23, 44, and 82 days post-injury for participants with an upper-limb burn (n = 20). Slopes were estimated from linear mixed models fit separately for each subtest using a spline function of days since injury. A significant slowing in the rate of improvement was observed between days 23 and 44 in the simple bilateral subtest (C; adjusted *p* = .038), followed by a re-acceleration by day 82 (adjusted *p* = .022). No other subtests showed statistically significant changes in the daily rate of change (all adjusted *ps* ≥ .520). Shaded bands represent 95% confidence intervals.

***S6.4 Short-form 36 Component Scores in Patients with Upper-limb Injuries***

Self-reported physical and mental health scores were evaluated at 23, 44, and 82 days post-injury in participants with an upper-limb burn injury using the SF36 Physical Component Summary and Mental Component Summary scores. For Physical Component Summary, a significant improvement was observed from day 23 to day 44 (+6.9%, adjusted *p* = .008), with a similar percent increase from day 23 to day 82 (+6.3%, adjusted *p* = .069). No meaningful change was detected between day 44 and day 82 (–0.6%, adjusted *p* = .834), indicating that gains in physical health stabilised by week 6. For Mental Component Summary, improvements were more gradual. Percent changes were +4.2% from day 23 to 44 (adjusted *p* = .395), +9.4% from day 23 to 82 (adjusted *p* = .115), and +4.9% from day 44 to 82 (adjusted *p* = .395), although none reached statistical significance. These findings indicate early improvements in perceived physical health, with more modest and variable gains in mental health over the subacute recovery period.

***References***

***S7* R *Code for Analysis***

\# Main manuscript ####

\# Pegboard ####

library(lme4)

library(emmeans)

library(dplyr)

library(tidyr)

library(stringr)

library(splines)

\# Scores

peg_tests <- c("dominant", "nondominant", "simple_bilateral", "assembly")

days_to_check <- c(23, 44, 82)

peg_model_results <- list()

peg_day_estimates <- list()

peg_day_comparisons <- list()

for (test in peg_tests) {

\# Fit the model

model <- lmer(

as.formula(paste(test, "~ ns(DAYS, df = 3) + (1 | ID)")),

data = df_track_peg_final1

)

\# Estimated marginal means at each timepoint

emms <- emmeans(model, ~ DAYS, at = list(DAYS = days_to_check))

est_df <- as.data.frame(emms) %>%

mutate(Test = test)

peg_day_estimates[[test]] <- est_df

\# Pairwise comparisons: Later - Earlier

contr_df <- contrast(emms, method = "revpairwise", adjust = "holm") %>%

as.data.frame() %>%

mutate(Test = test)

\# Extract numeric days from contrast labels

day_labels <- str_extract_all(as.character(contr_df$contrast), "\\d+")

parsed_days <- do.call(rbind, lapply(day_labels, as.numeric))

colnames(parsed_days) <- c("Later", "Earlier")

\# Calculate percent change: (Later - Earlier) / Earlier

contr_df <- contr_df %>%

bind_cols(as.data.frame(parsed_days)) %>%

left_join(est_df %>% select(DAYS, emmean), by = c("Earlier" = "DAYS")) %>%

mutate(

contrast = paste(Later, Earlier, sep = " - "),

percent_change = (estimate / emmean) * 100

) %>%

select(contrast, estimate, SE, df, t.ratio, p.value, Test, percent_change)

peg_day_comparisons[[test]] <- contr_df

peg_model_results[[test]] <- model

}

\# Combine results across all pegboard tests

peg_day_estimates_all <- bind_rows(peg_day_estimates)

peg_day_comparisons_all <- bind_rows(peg_day_comparisons)

\# Print output

print(peg_day_estimates_all)

print(peg_day_comparisons_all)

\# Slopes

peg_tests <- c("dominant", "nondominant", "simple_bilateral", "assembly")

days_to_check <- c(23, 44, 82)

\# Storage lists

peg_trend_estimates <- list()

peg_trend_comparisons <- list()

peg_model_results <- list()

for (test in peg_tests) {

\# Fit model with spline

model <- lmer(

as.formula(paste(test, "~ ns(DAYS, df = 3) + (1 | ID)")),

data = df_track_peg_final1

)

\# Estimate slopes (trends) at specified DAYS

emtr <- emtrends(model, ~ DAYS, var = "DAYS", at = list(DAYS = days_to_check))

peg_trend_estimates[[test]] <- as.data.frame(emtr) %>%

mutate(Test = test)

\# Pairwise contrasts of slopes

contrasts <- contrast(emtr, method = "revpairwise", adjust = "holm")

peg_trend_comparisons[[test]] <- as.data.frame(contrasts) %>%

mutate(Test = test)

\# Save model

peg_model_results[[test]] <- model

}

\# Combine across tests

peg_trend_estimates_all <- bind_rows(peg_trend_estimates)

peg_trend_comparisons_all <- bind_rows(peg_trend_comparisons)

\# Output

print(peg_trend_estimates_all)

print(peg_trend_comparisons_all)

\# SF36 ####

\# Scores

sf36_vars <- c("PCS", "MCS")

days_to_check <- c(23, 44, 82) # Choose relevant days in your data

results_list <- list()

estimates_list <- list()

comparisons_list <- list()

for (var in sf36_vars) {

\# Fit mixed model with natural spline for DAYS

model <- lmer(

as.formula(paste(var, "~ ns(DAYS, df = 3) + (1 | ID)")),

data = sf36_scored_new

)

\# Estimated marginal means at specific DAYS

emms <- emmeans(model, ~ DAYS, at = list(DAYS = days_to_check))

est_df <- as.data.frame(emms) %>%

mutate(Variable = var)

estimates_list[[var]] <- est_df

\# Pairwise comparisons between days (later vs earlier)

contr_df <- contrast(emms, method = "revpairwise", adjust = "holm") %>%

as.data.frame() %>%

mutate(Variable = var)

\# Extract numeric days from contrast labels

day_labels <- str_extract_all(as.character(contr_df$contrast), "\\d+")

parsed_days <- do.call(rbind, lapply(day_labels, as.numeric))

colnames(parsed_days) <- c("Later", "Earlier")

\# Calculate percent change: (Later - Earlier) / Earlier * 100

contr_df <- contr_df %>%

bind_cols(as.data.frame(parsed_days)) %>%

left_join(est_df %>% select(DAYS, emmean), by = c("Earlier" = "DAYS")) %>%

mutate(

contrast = paste(Later, Earlier, sep = " - "),

percent_change = (estimate / emmean) * 100

) %>%

select(contrast, estimate, SE, df, t.ratio, p.value, Variable, percent_change)

comparisons_list[[var]] <- contr_df

results_list[[var]] <- model

}

\# Combine results

estimates_all <- bind_rows(estimates_list)

comparisons_all <- bind_rows(comparisons_list)

\# Print results

print(estimates_all)

print(comparisons_all)

\# Slopes

sf36_vars <- c("PCS", "MCS")

days_to_check <- c(23, 44, 82)

results_list <- list()

trend_estimates_list <- list()

trend_comparisons_list <- list()

for (var in sf36_vars) {

\# Fit mixed model with natural spline for DAYS

model <- lmer(

as.formula(paste(var, "~ ns(DAYS, df = 3) + (1 | ID)")),

data = sf36_scored_new

)

\# Estimate marginal trends (slopes) at each day

emtr <- emtrends(model, ~ DAYS, var = "DAYS", at = list(DAYS = days_to_check))

trend_df <- as.data.frame(emtr) %>%

mutate(Variable = var)

trend_estimates_list[[var]] <- trend_df

\# Pairwise comparisons of slopes across days

contr_df <- contrast(emtr, method = "revpairwise", adjust = "holm") %>%

as.data.frame() %>%

mutate(Variable = var)

\# Extract numeric day labels from contrast names

day_labels <- str_extract_all(as.character(contr_df$contrast), "\\d+")

parsed_days <- do.call(rbind, lapply(day_labels, as.numeric))

colnames(parsed_days) <- c("Later", "Earlier")

\# Percent change in slope: (Later - Earlier) / |Earlier| * 100

contr_df <- contr_df %>%

bind_cols(as.data.frame(parsed_days)) %>%

left_join(trend_df %>% select(DAYS, trend = DAYS.trend), by = c("Earlier" = "DAYS")) %>%

mutate(

contrast = paste(Later, Earlier, sep = " - "),

percent_change = (estimate / abs(trend)) * 100

) %>%

select(contrast, estimate, SE, df, t.ratio, p.value, Variable, percent_change)

trend_comparisons_list[[var]] <- contr_df

results_list[[var]] <- model

}

\# Combine results

slope_estimates_all <- bind_rows(trend_estimates_list)

slope_contrasts_all <- bind_rows(trend_comparisons_list)

\# Print results

print(slope_estimates_all)

print(slope_contrasts_all)

\# MT and 1 mV ####

\# Intensities

tms_vars <- c("OnemV", "RMT")

days_to_check <- c(23, 44, 82)

tms_model_results <- list()

tms_day_estimates <- list()

tms_day_comparisons <- list()

for (var in tms_vars) {

\# Fit model using natural spline

model <- lmer(

as.formula(paste(var, "~ ns(DAYS, df = 3) + (1 | ID)")),

data = mt

)

\# Estimated marginal means

emms <- emmeans(model, ~ DAYS, at = list(DAYS = days_to_check))

est_df <- as.data.frame(emms) %>%

mutate(Variable = var)

tms_day_estimates[[var]] <- est_df

\# Pairwise comparisons

contr_df <- contrast(emms, method = "revpairwise", adjust = "holm") %>%

as.data.frame() %>%

mutate(Variable = var)

\# Extract day labels

day_labels <- str_extract_all(as.character(contr_df$contrast), "\\d+")

parsed_days <- do.call(rbind, lapply(day_labels, as.numeric))

colnames(parsed_days) <- c("Later", "Earlier")

\# Join earlier day mean for percent change

contr_df <- contr_df %>%

bind_cols(as.data.frame(parsed_days)) %>%

left_join(est_df %>% select(DAYS, emmean), by = c("Earlier" = "DAYS")) %>%

mutate(

contrast = paste(Later, Earlier, sep = " - "),

percent_change = (estimate / emmean) * 100

) %>%

select(contrast, estimate, SE, df, t.ratio, p.value, Variable, percent_change)

tms_day_comparisons[[var]] <- contr_df

tms_model_results[[var]] <- model

}

tms_day_estimates_all <- bind_rows(tms_day_estimates)

tms_day_comparisons_all <- bind_rows(tms_day_comparisons)

print(tms_day_estimates_all)

print(tms_day_comparisons_all)

\# Slopes

tms_vars <- c("OnemV", "RMT")

days_to_check <- c(23, 44, 82)

\# Storage lists

tms_trend_estimates <- list()

tms_trend_comparisons <- list()

tms_model_results <- list()

for (var in tms_vars) {

\# Fit model with spline

model <- lmer(

as.formula(paste(var, "~ ns(DAYS, df = 3) + (1 | ID)")),

data = mt

)

\# Estimate slope of outcome with respect to DAYS at specific timepoints

emtr <- emtrends(model, ~ DAYS, var = "DAYS", at = list(DAYS = days_to_check))

tms_trend_estimates[[var]] <- as.data.frame(emtr) %>%

mutate(Variable = var)

\# Pairwise contrasts of slopes between days

contrast_results <- contrast(emtr, method = "revpairwise", adjust = "holm")

tms_trend_comparisons[[var]] <- as.data.frame(contrast_results) %>%

mutate(Variable = var)

\# Save model

tms_model_results[[var]] <- model

}

\# Combine into one data frame

tms_trend_estimates_all <- bind_rows(tms_trend_estimates)

tms_trend_comparisons_all <- bind_rows(tms_trend_comparisons)

\# Output

print(tms_trend_estimates_all)

print(tms_trend_comparisons_all)

\# Baseline TMS ####

\# Amplitude

\# SP

days_to_check <- c(23, 44, 82)

results_list <- list()

for (d in days_to_check) {

emms_day <- emmeans(

track_final_model_bobyqa,

~ STATE,

at = list(DAYS = d, BLOCK = "PRE")

)

df <- as.data.frame(emms_day)

\# Determine correct mean column

mean_col <- if ("emmean" %in% names(df)) "emmean" else "response"

\# Extract SP row and manually assign value

sp_row <- df[df$STATE == "SP", ]

sp_row$estimate <- sp_row[[mean_col]]

sp_row$DAY <- d

results_list[[as.character(d)]] <- sp_row[, c("DAY", "estimate", "SE")]

}

\# Combine results

df_sp_all_days <- bind_rows(results_list)

\# Pairwise comparisons function

pairwise_day_contrasts <- function(df, value_col = "estimate", se_col = "SE", day_col = "DAY") {

days <- sort(unique(df[[day_col]]))

results <- list()

for(i in 1:(length(days)-1)) {

for(j in (i+1):length(days)) {

d1 <- days[i]

d2 <- days[j]

row1 <- df[df[[day_col]] == d1, ]

row2 <- df[df[[day_col]] == d2, ]

est_diff <- row2[[value_col]] - row1[[value_col]]

se_diff <- sqrt(row2[[se_col]]^2 + row1[[se_col]]^2)

z <- est_diff / se_diff

p <- 2 * pnorm(-abs(z))

ratio <- exp(est_diff)

percent_change <- (ratio - 1) * 100

results[[paste(d2, "vs", d1)]] <- data.frame(

contrast = "SP amplitude",

day_comparison = paste(d2, "vs", d1),

estimate = est_diff,

SE = se_diff,

z.ratio = z,

p.value = p,

ratio = ratio,

percent_change = percent_change

)

}

}

bind_rows(results)

}

\# Run pairwise comparison

sp_amplitude_contrasts <- pairwise_day_contrasts(df_sp_all_days)

\# Apply Holm correction

sp_amplitude_contrasts <- sp_amplitude_contrasts %>%

mutate(p.adjusted = p.adjust(p.value, method = "holm"))

\# Correct Holm approach

sp_amplitude_contrasts <- sp_amplitude_contrasts %>%

group_by(contrast) %>%

mutate(p.adjusted = p.adjust(p.value, method = "holm")) %>%

ungroup()

\# Print results

print(as.data.frame(sp_amplitude_contrasts, digits = 4))

\# Model values

df_sp_all_days <- df_sp_all_days %>%

mutate(

response = exp(estimate),

lower_CI = exp(estimate - 1.96 * SE),

upper_CI = exp(estimate + 1.96 * SE)

)

print(df_sp_all_days, digits = 4)

\# ICI

days_to_check <- c(23, 44, 82)

results_list <- list()

for (d in days_to_check) {

\# 1. Estimate marginal means for STATE at each DAY (PRE block only)

emms_day <- emmeans(

track_final_model_bobyqa,

~ STATE,

at = list(DAYS = d, BLOCK = "PRE")

)

\# 2. Compute contrasts: LICI - SP, SICI - SP

ici_diff <- contrast(

emms_day,

method = "trt.vs.ctrl",

ref = "SP"

)

\# 3. Save results

df <- as.data.frame(ici_diff) %>%

mutate(DAY = d)

results_list[[as.character(d)]] <- df

}

\# 4. Combine all timepoints

df_all_days <- bind_rows(results_list) %>%

arrange(contrast, DAY) %>%

mutate(

ratio = exp(estimate),

percent_change = (ratio - 1) * 100

)

\# 5. Compare contrast estimates across days (e.g., 82 vs 23)

pairwise_day_contrasts <- function(df, value_col = "estimate", se_col = "SE", day_col = "DAY") {

days <- sort(unique(df[[day_col]]))

results <- list()

for(i in 1:(length(days)-1)) {

for(j in (i+1):length(days)) {

d1 <- days[i]

d2 <- days[j]

row1 <- df[df[[day_col]] == d1, ]

row2 <- df[df[[day_col]] == d2, ]

est_diff <- row2[[value_col]] - row1[[value_col]]

se_diff <- sqrt(row2[[se_col]]^2 + row1[[se_col]]^2)

z <- est_diff / se_diff

p <- 2 * pnorm(-abs(z))

ratio <- exp(est_diff)

percent_change <- (ratio - 1) * 100

results[[paste(d2, "vs", d1)]] <- data.frame(

day_comparison = paste(d2, "vs", d1),

estimate = est_diff,

SE = se_diff,

z.ratio = z,

p.value = p,

ratio = ratio,

percent_change = percent_change

)

}

}

bind_rows(results)

}

\# 6. Apply the function grouped by contrast

day_pairwise_df <- df_all_days %>%

group_by(contrast) %>%

nest() %>%

mutate(

pairwise = map(data, ~pairwise_day_contrasts(.x))

) %>%

unnest(pairwise) %>%

ungroup()

\# 7. Final cleaned output with Holm adjustment

essential_day_pairwise_df <- day_pairwise_df %>%

select(

contrast,

day_comparison,

estimate,

SE,

p.value,

ratio,

percent_change

) %>%

mutate(p.adjusted = p.adjust(p.value, method = "holm"))

\# Correct Holm approach

essential_day_pairwise_df <- day_pairwise_df %>%

select(

contrast,

day_comparison,

estimate,

SE,

p.value,

ratio,

percent_change

) %>%

group_by(contrast) %>%

mutate(p.adjusted = p.adjust(p.value, method = "holm")) %>%

ungroup()

\# 8. View result

print(as.data.frame(essential_day_pairwise_df), digits = 4)

\# Model ratios

model_amplitude_ratios <- df_all_days %>%

select(DAY, contrast, estimate, SE, ratio) %>%

mutate(

lower_CI = exp(estimate - 1.96 * SE),

upper_CI = exp(estimate + 1.96 * SE)

)

print(model_amplitude_ratios, digits = 4)

\# Slope

\# SP

days_to_check <- c(23, 44, 82)

sp_results_list <- list()

for (d in days_to_check) {

emtr_day <- emtrends(

track_final_model_bobyqa,

~ STATE,

var = "DAYS",

at = list(DAYS = d, BLOCK = "PRE") # Restrict to PRE block

)

emtr_day_sp <- as.data.frame(emtr_day) %>%

filter(STATE == "SP") %>%

mutate(DAY = d)

sp_results_list[[as.character(d)]] <- emtr_day_sp

}

\# Combine SP-only slope estimates

sp_df_all_days <- bind_rows(sp_results_list) %>%

select(DAY, estimate = DAYS.trend, SE)

\# Function to compare SP slopes between days

pairwise_day_contrasts <- function(df, value_col = "estimate", se_col = "SE", day_col = "DAY") {

days <- sort(unique(df[[day_col]]))

results <- list()

for(i in 1:(length(days)-1)) {

for(j in (i+1):length(days)) {

d1 <- days[i]

d2 <- days[j]

row1 <- df[df[[day_col]] == d1, ]

row2 <- df[df[[day_col]] == d2, ]

est_diff <- row2[[value_col]] - row1[[value_col]]

se_diff <- sqrt(row2[[se_col]]^2 + row1[[se_col]]^2)

z <- est_diff / se_diff

p <- 2 * pnorm(-abs(z))

ratio <- exp(est_diff)

percent_change <- (ratio - 1) * 100

results[[paste(d2, "vs", d1)]] <- data.frame(

contrast = "SP slope",

day_comparison = paste(d2, "vs", d1),

estimate = est_diff,

SE = se_diff,

p.value = p,

ratio = ratio,

percent_change = percent_change

)

}

}

bind_rows(results)

}

\# Compute pairwise comparisons

sp_day_pairwise_df <- pairwise_day_contrasts(sp_df_all_days)

\# Add Holm-corrected p-values

sp_day_pairwise_df <- sp_day_pairwise_df %>%

mutate(p.adjusted = p.adjust(p.value, method = "holm"))

\# Correct Holm approach

sp_day_pairwise_df <- sp_day_pairwise_df %>%

group_by(contrast) %>%

mutate(p.adjusted = p.adjust(p.value, method = "holm")) %>%

ungroup()

\# Final output

print(as.data.frame(sp_day_pairwise_df, digits = 4))

\# Model values

sp_df_all_days <- sp_df_all_days %>%

mutate(

slope_ratio_per_day = exp(estimate),

lower_CI = exp(estimate - 1.96 * SE),

upper_CI = exp(estimate + 1.96 * SE)

)

print(sp_df_all_days, digits = 4)

\# ICI

days_to_check <- c(23, 44, 82)

results_list <- list()

\# 2. Extract emtrends and compute contrasts for each day

for (d in days_to_check) {

emtr_day <- emtrends(

track_final_model_bobyqa,

~ STATE,

var = "DAYS",

at = list(DAYS = d, BLOCK = "PRE") # Restrict to PRE block

)

ici_diff <- contrast(

emtr_day,

method = "trt.vs.ctrl",

ref = "SP"

)

df <- as.data.frame(ici_diff) %>%

mutate(DAY = d)

results_list[[as.character(d)]] <- df

}

\# 3. Combine all results and calculate ratios

df_all_days <- bind_rows(results_list) %>%

arrange(contrast, DAY) %>%

mutate(

ratio = exp(estimate),

percent_change = (ratio - 1) * 100

)

\# 4. Define pairwise comparison function

pairwise_day_contrasts <- function(df, value_col = "estimate", se_col = "SE", day_col = "DAY") {

days <- sort(unique(df[[day_col]]))

results <- list()

for (i in 1:(length(days)-1)) {

for (j in (i+1):length(days)) {

d1 <- days[i]

d2 <- days[j]

row1 <- df[df[[day_col]] == d1, ]

row2 <- df[df[[day_col]] == d2, ]

est_diff <- row2[[value_col]] - row1[[value_col]]

se_diff <- sqrt(row2[[se_col]]^2 + row1[[se_col]]^2)

z <- est_diff / se_diff

p <- 2 * pnorm(-abs(z))

ratio <- exp(est_diff)

percent_change <- (ratio - 1) * 100

results[[paste(d2, "vs", d1)]] <- data.frame(

day_comparison = paste(d2, "vs", d1),

estimate = est_diff,

SE = se_diff,

z.ratio = z,

p.value = p,

ratio = ratio,

percent_change = percent_change

)

}

}

bind_rows(results)

}

\# 5. Run pairwise comparisons within each contrast group

day_pairwise_df <- df_all_days %>%

group_by(contrast) %>%

nest() %>%

mutate(pairwise = map(data, ~pairwise_day_contrasts(.x))) %>%

unnest(pairwise) %>%

ungroup()

\# 6. Apply Holm correction only once

essential_day_pairwise_df <- day_pairwise_df %>%

select(

contrast,

day_comparison,

estimate,

SE,

p.value,

ratio,

percent_change

) %>%

mutate(p.adjusted = p.adjust(p.value, method = "holm"))

\# Correct Holm approach

essential_day_pairwise_df <- day_pairwise_df %>%

select(

contrast,

day_comparison,

estimate,

SE,

p.value,

ratio,

percent_change

) %>%

group_by(contrast) %>%

mutate(p.adjusted = p.adjust(p.value, method = "holm")) %>%

ungroup()

\# 7. View result

print(as.data.frame(essential_day_pairwise_df), digits = 4)

\# Slope model ratios

model_slope_ratios <- df_all_days %>%

select(DAY, contrast, estimate, SE, ratio, percent_change) %>%

mutate(

lower_CI = exp(estimate - 1.96 * SE),

upper_CI = exp(estimate + 1.96 * SE)

)

print(model_slope_ratios, digits = 4)

\# POST-PAS ####

\# Amplitude

\# SP

days_to_check <- c(23, 44, 82)

results_list <- list()	# For SP contrasts (BLOCK - PRE)

sp_response_list <- list()	# For raw SP means per BLOCK × DAY

for (d in days_to_check) {

\# 1. Estimate marginal means for BLOCK × STATE at each DAY

emm_day <- emmeans(

track_final_model_bobyqa,

~ BLOCK * STATE,

at = list(DAYS = d)

)

\# 2. Keep SP only (reliable across all versions)

emm_sp <- summary(emm_day) %>%

filter(STATE == "SP") %>%

select(BLOCK, estimate = emmean, SE) %>%

mutate(STATE_trt.vs.ctrl = "SP mean", DAY = d)

\# 3. Store raw SP values

sp_response_list[[as.character(d)]] <- emm_sp

\# 4. Manually compute BLOCK vs PRE contrasts

blocks_to_compare <- c("POST-PAS", "15 MIN POST-PAS", "30 MIN POST-PAS")

block_contrasts <- lapply(blocks_to_compare, function(block) {

pre <- emm_sp %>% filter(BLOCK == "PRE")

comp <- emm_sp %>% filter(BLOCK == block)

est <- comp$estimate - pre$estimate

se <- sqrt(comp$SE^2 + pre$SE^2)

data.frame(

BLOCK_pairwise = paste0("PRE - (", block, ")"),

STATE_trt.vs.ctrl = "SP mean",

estimate = est,

SE = se

)

}) %>%

bind_rows() %>%

mutate(DAY = d)

results_list[[as.character(d)]] <- block_contrasts

}

\# 5. Combine contrast results

df_all_days <- bind_rows(results_list) %>%

arrange(STATE_trt.vs.ctrl, BLOCK_pairwise, DAY)

\# 6. Clean contrast labels and compute response-scale interpretation

df_all_days_cleaned <- df_all_days %>%

mutate(

BLOCK_pairwise = sub("^PRE - \\((.*)\\)$", "\\1 - PRE", BLOCK_pairwise),

ratio = exp(estimate),

percent_change = (ratio - 1) * 100

)

\# 7. Compute pairwise day contrasts within each BLOCK comparison

pairwise_day_contrasts <- function(df, value_col = "estimate", se_col = "SE", day_col = "DAY") {

days <- sort(unique(df[[day_col]]))

results <- list()

for (i in 1:(length(days) - 1)) {

for (j in (i + 1):length(days)) {

d1 <- days[i]; d2 <- days[j]

row1 <- df[df[[day_col]] == d1, ]

row2 <- df[df[[day_col]] == d2, ]

est_diff <- row2[[value_col]] - row1[[value_col]]

se_diff <- sqrt(row2[[se_col]]^2 + row1[[se_col]]^2)

z <- est_diff / se_diff

p <- 2 * pnorm(-abs(z))

ratio <- exp(est_diff)

percent_change <- (ratio - 1) * 100

results[[paste(d2, "vs", d1)]] <- data.frame(

day_comparison = paste(d2, "vs", d1),

estimate = est_diff,

SE = se_diff,

z.ratio = z,

p.value = p,

ratio = ratio,

percent_change = percent_change

)

}

}

bind_rows(results)

}

\# 8. Apply pairwise comparison function

day_pairwise_df <- df_all_days_cleaned %>%

group_by(STATE_trt.vs.ctrl, BLOCK_pairwise) %>%

nest() %>%

mutate(pairwise = map(data, ~pairwise_day_contrasts(.x))) %>%

unnest(pairwise) %>%

ungroup()

\# 9. Apply Holm correction

essential_day_pairwise_df <- day_pairwise_df %>%

select(

BLOCK_pairwise,

STATE_trt.vs.ctrl,

day_comparison,

estimate,

SE,

p.value,

ratio,

percent_change

) %>%

group_by(BLOCK_pairwise, STATE_trt.vs.ctrl) %>%

mutate(p.adjusted = p.adjust(p.value, method = "holm")) %>%

ungroup()

\# 10. Print contrast results

print(as.data.frame(essential_day_pairwise_df), digits = 4)

\# 11. Combine and print raw SP means by BLOCK × DAY

sp_response_df <- bind_rows(sp_response_list) %>%

arrange(BLOCK, DAY) %>%

mutate(

ratio = exp(estimate),

percent_change = (ratio - 1) * 100

)

print(sp_response_df, digits = 4)

\# ICI

days_to_check <- c(23, 44, 82)

results_list <- list()	# For PRE comparisons across days

response_results_list <- list() # For raw LICI/SP and SICI/SP marginal means

for (d in days_to_check) {

\# 1. Estimate marginal means for BLOCK × STATE at each DAY (no GROUP)

emm_day <- emmeans(

track_final_model_bobyqa,

~ BLOCK * STATE,

at = list(DAYS = d)

)

\# 2. Compute within-BLOCK differences (LICI - SP, SICI - SP)

ici_diff <- contrast(

emm_day,

interaction = "trt.vs.ctrl",

by = "BLOCK",

ref = "SP"

)

\# 3. Compare across BLOCKs for each contrast (PRE vs others only)

block_contrasts <- contrast(

ici_diff,

interaction = "pairwise",

by = "STATE_trt.vs.ctrl",

ref = "PRE",

adjust = "none"

)

\# 4a. Store and tag with day, keep only PRE comparisons (will flip sign below)

df_block <- as.data.frame(block_contrasts) %>%

filter(grepl("^PRE - ", BLOCK_pairwise)) %>%

mutate(DAY = d)

results_list[[as.character(d)]] <- df_block

\# 4b. Save raw LICI/SP and SICI/SP marginal means at each BLOCK

df_response <- as.data.frame(ici_diff) %>%

mutate(

DAY = d,

ratio = exp(estimate),

percent_change = (ratio - 1) * 100

)

response_results_list[[as.character(d)]] <- df_response

}

\# 5. Combine results

df_all_days <- bind_rows(results_list) %>%

arrange(STATE_trt.vs.ctrl, BLOCK_pairwise, DAY)

\# 6. Flip contrast direction to BLOCK - PRE and recompute response scale

df_all_days_flipped <- df_all_days %>%

mutate(

BLOCK_pairwise = sub("^PRE - \\((.*)\\)", "\\1 - PRE", BLOCK_pairwise),

estimate = -estimate,

ratio = exp(estimate),

percent_change = (ratio - 1) * 100

)

\# 7. Pairwise differences by day

pairwise_day_contrasts <- function(df, value_col = "estimate", se_col = "SE", day_col = "DAY") {

days <- sort(unique(df[[day_col]]))

results <- list()

for (i in 1:(length(days) - 1)) {

for (j in (i + 1):length(days)) {

d1 <- days[i]

d2 <- days[j]

row1 <- df[df[[day_col]] == d1, ]

row2 <- df[df[[day_col]] == d2, ]

est_diff <- row2[[value_col]] - row1[[value_col]]

se_diff <- sqrt(row2[[se_col]]^2 + row1[[se_col]]^2)

z <- est_diff / se_diff

p <- 2 * pnorm(-abs(z))

ratio <- exp(est_diff)

percent_change <- (ratio - 1) * 100

results[[paste(d2, "vs", d1)]] <- data.frame(

day_comparison = paste(d2, "vs", d1),

estimate = est_diff,

SE = se_diff,

z.ratio = z,

p.value = p,

ratio = ratio,

percent_change = percent_change

)

}

}

bind_rows(results, .id = NULL)

}

\# 8. Apply the function by contrast

day_pairwise_df <- df_all_days_flipped %>%

group_by(STATE_trt.vs.ctrl, BLOCK_pairwise) %>%

nest() %>%

mutate(

pairwise = map(data, ~pairwise_day_contrasts(.x))

) %>%

unnest(pairwise) %>%

ungroup()

\# 9. Tidy up and apply Holm correction

essential_day_pairwise_df <- day_pairwise_df %>%

select(

BLOCK_pairwise,

STATE_trt.vs.ctrl,

day_comparison,

estimate,

SE,

p.value,

ratio,

percent_change

) %>%

group_by(BLOCK_pairwise, STATE_trt.vs.ctrl) %>%

mutate(

p.adjusted = p.adjust(p.value, method = "holm")

) %>%

ungroup()

\# 10. Print pairwise results

print(as.data.frame(essential_day_pairwise_df), digits = 4)

\# 11. Combine and print raw response values (LICI/SP, SICI/SP at each DAY × BLOCK)

response_by_block_day <- bind_rows(response_results_list) %>%

arrange(STATE_trt.vs.ctrl, BLOCK, DAY) %>%

select(DAY, BLOCK, STATE_trt.vs.ctrl, estimate, SE, ratio, percent_change, p.value)

print(response_by_block_day, digits = 4)

\# Slope

\# SP

days_to_check <- c(23, 44, 82)

results_list <- list()	# For SP slope contrasts (BLOCK - PRE)

sp_response_list <- list()	# For raw SP slopes per BLOCK × DAY

for (d in days_to_check) {

\# 1. Estimate slopes for BLOCK × STATE at each DAY

emtr_day <- emtrends(

track_final_model_bobyqa,

~ BLOCK * STATE,

var = "DAYS",

at = list(DAYS = d)

)

\# 2. Keep SP only

df_sp <- as.data.frame(emtr_day) %>%

filter(STATE == "SP") %>%

select(BLOCK, estimate = DAYS.trend, SE) %>%

mutate(STATE_trt.vs.ctrl = "SP slope", DAY = d)

\# 3. Store raw SP values

sp_response_list[[as.character(d)]] <- df_sp

\# 4. Manually compute BLOCK vs PRE contrasts

blocks_to_compare <- c("POST-PAS", "15 MIN POST-PAS", "30 MIN POST-PAS")

block_contrasts <- lapply(blocks_to_compare, function(block) {

pre <- df_sp %>% filter(BLOCK == "PRE")

comp <- df_sp %>% filter(BLOCK == block)

est <- comp$estimate - pre$estimate

se <- sqrt(comp$SE^2 + pre$SE^2)

data.frame(

BLOCK_pairwise = paste0("PRE - (", block, ")"),

STATE_trt.vs.ctrl = "SP slope",

estimate = est,

SE = se

)

}) %>%

bind_rows() %>%

mutate(DAY = d)

results_list[[as.character(d)]] <- block_contrasts

}

\# 5. Combine contrast results

df_all_days <- bind_rows(results_list) %>%

arrange(STATE_trt.vs.ctrl, BLOCK_pairwise, DAY)

\# 6. Clean contrast labels and compute response-scale interpretation

df_all_days_cleaned <- df_all_days %>%

mutate(

BLOCK_pairwise = sub("^PRE - \\((.*)\\)$", "\\1 - PRE", BLOCK_pairwise),

ratio = exp(estimate),

percent_change = (ratio - 1) * 100

)

\# 7. Compute pairwise day contrasts within each BLOCK comparison

pairwise_day_contrasts <- function(df, value_col = "estimate", se_col = "SE", day_col = "DAY") {

days <- sort(unique(df[[day_col]]))

results <- list()

for (i in 1:(length(days)-1)) {

for (j in (i+1):length(days)) {

d1 <- days[i]; d2 <- days[j]

row1 <- df[df[[day_col]] == d1, ]

row2 <- df[df[[day_col]] == d2, ]

est_diff <- row2[[value_col]] - row1[[value_col]]

se_diff <- sqrt(row2[[se_col]]^2 + row1[[se_col]]^2)

z <- est_diff / se_diff

p <- 2 * pnorm(-abs(z))

ratio <- exp(est_diff)

percent_change <- (ratio - 1) * 100

results[[paste(d2, "vs", d1)]] <- data.frame(

day_comparison = paste(d2, "vs", d1),

estimate = est_diff,

SE = se_diff,

z.ratio = z,

p.value = p,

ratio = ratio,

percent_change = percent_change

)

}

}

bind_rows(results)

}

\# 8. Apply pairwise comparison function

day_pairwise_df <- df_all_days_cleaned %>%

group_by(STATE_trt.vs.ctrl, BLOCK_pairwise) %>%

nest() %>%

mutate(pairwise = map(data, ~pairwise_day_contrasts(.x))) %>%

unnest(pairwise) %>%

ungroup()

\# 9. Apply Holm correction

essential_day_pairwise_df <- day_pairwise_df %>%

select(

BLOCK_pairwise,

STATE_trt.vs.ctrl,

day_comparison,

estimate,

SE,

p.value,

ratio,

percent_change

) %>%

group_by(BLOCK_pairwise, STATE_trt.vs.ctrl) %>%

mutate(p.adjusted = p.adjust(p.value, method = "holm")) %>%

ungroup()

\# 10. Print contrast results

print(as.data.frame(essential_day_pairwise_df), digits = 4)

\# 11. Combine and print raw SP slope estimates by BLOCK × DAY

sp_response_df <- bind_rows(sp_response_list) %>%

arrange(BLOCK, DAY) %>%

mutate(

ratio = exp(estimate),

percent_change = (ratio - 1) * 100

)

print(sp_response_df, digits = 4)

\# ICI

days_to_check <- c(23, 44, 82)

results_list <- list()	# For PRE comparisons across days

response_results_list <- list() # For raw LICI/SP and SICI/SP values

for (d in days_to_check) {

\# 1. Estimate slopes for BLOCK × STATE at each DAY (no GROUP)

emtr_day <- emtrends(

track_final_model_bobyqa,

~ BLOCK * STATE,

var = "DAYS",

at = list(DAYS = d)

)

\# 2. Compute within-BLOCK differences (LICI - SP, SICI - SP)

ici_diff <- contrast(

emtr_day,

interaction = "trt.vs.ctrl",

by = "BLOCK",

ref = "SP"

)

\# 3. Compare across BLOCKs for each contrast (PRE vs others only)

block_contrasts <- contrast(

ici_diff,

interaction = "pairwise",

by = "STATE_trt.vs.ctrl",

ref = "PRE",

adjust = "none"

)

\# 4a. Store and tag with day, keep only PRE comparisons (will flip sign below)

df_block <- as.data.frame(block_contrasts) %>%

filter(grepl("^PRE - ", BLOCK_pairwise)) %>%

mutate(DAY = d)

results_list[[as.character(d)]] <- df_block

\# 4b. Save raw LICI/SP and SICI/SP estimates at each BLOCK

df_response <- as.data.frame(ici_diff) %>%

mutate(

DAY = d,

ratio = exp(estimate),

percent_change = (ratio - 1) * 100

)

response_results_list[[as.character(d)]] <- df_response

}

\# 5. Combine results

df_all_days <- bind_rows(results_list) %>%

arrange(STATE_trt.vs.ctrl, BLOCK_pairwise, DAY)

\# 6. Flip contrast direction to BLOCK - PRE and recompute response scale

df_all_days_flipped <- df_all_days %>%

mutate(

BLOCK_pairwise = sub("^PRE - \\((.*)\\)", "\\1 - PRE", BLOCK_pairwise),

estimate = -estimate,

ratio = exp(estimate),

percent_change = (ratio - 1) * 100

)

\# 7. Pairwise differences by day

pairwise_day_contrasts <- function(df, value_col = "estimate", se_col = "SE", day_col = "DAY") {

days <- sort(unique(df[[day_col]]))

results <- list()

for (i in 1:(length(days) - 1)) {

for (j in (i + 1):length(days)) {

d1 <- days[i]

d2 <- days[j]

row1 <- df[df[[day_col]] == d1, ]

row2 <- df[df[[day_col]] == d2, ]

est_diff <- row2[[value_col]] - row1[[value_col]]

se_diff <- sqrt(row2[[se_col]]^2 + row1[[se_col]]^2)

z <- est_diff / se_diff

p <- 2 * pnorm(-abs(z))

ratio <- exp(est_diff)

percent_change <- (ratio - 1) * 100

results[[paste(d2, "vs", d1)]] <- data.frame(

day_comparison = paste(d2, "vs", d1),

estimate = est_diff,

SE = se_diff,

z.ratio = z,

p.value = p,

ratio = ratio,

percent_change = percent_change

)

}

}

bind_rows(results, .id = NULL)

}

\# 8. Apply the function by contrast

day_pairwise_df <- df_all_days_flipped %>%

group_by(STATE_trt.vs.ctrl, BLOCK_pairwise) %>%

nest() %>%

mutate(

pairwise = map(data, ~pairwise_day_contrasts(.x))

) %>%

unnest(pairwise) %>%

ungroup()

\# 9. Tidy up and apply Holm correction

essential_day_pairwise_df <- day_pairwise_df %>%

select(

BLOCK_pairwise,

STATE_trt.vs.ctrl,

day_comparison,

estimate,

SE,

p.value,

ratio,

percent_change

) %>%

group_by(BLOCK_pairwise, STATE_trt.vs.ctrl) %>%

mutate(

p.adjusted = p.adjust(p.value, method = "holm")

) %>%

ungroup()

\# 10. Print pairwise results

print(as.data.frame(essential_day_pairwise_df), digits = 4)

\# 11. Combine and print raw response values (LICI/SP, SICI/SP at each DAY × BLOCK)

response_by_block_day <- bind_rows(response_results_list) %>%

arrange(STATE_trt.vs.ctrl, BLOCK, DAY) %>%

select(DAY, BLOCK, STATE_trt.vs.ctrl, estimate, SE, ratio, percent_change, p.value)

print(response_by_block_day, digits = 4)

\# Supplementary Materials ####

\# Discrete analysis

\# SF36 ####

library(emmeans)

library(dplyr)

library(tibble)

\# === Bootstrap TRACK TP{1,2,3} vs CON TP1 for one outcome (LM + cluster bootstrap) ===

boot_trk_vs_con <- function(var, dat, nsim = 1000) {

wanted <- c("TRACK TP1 - CON TP1", "TRACK TP2 - CON TP1", "TRACK TP3 - CON TP1")

\## Parametric (reference) using plain LM

m	<- stats::lm(stats::as.formula(paste(var, "~ GROUP * TP")), data = dat)

emm <- emmeans::emmeans(m, ~ GROUP:TP)

ref <- which(emm@grid$GROUP == "CON" & emm@grid$TP == "TP1")

param <- as.data.frame(

summary(emmeans::contrast(emm, "trt.vs.ctrl", ref = ref), infer = c(TRUE, TRUE))

) |>

dplyr::filter(contrast %in% wanted) |>

dplyr::select(contrast, estimate, SE, df, lower.CL, upper.CL, t.ratio, p.value)

\## Nonparametric cluster bootstrap (resample IDs WITH replacement)

boot_mat <- replicate(nsim, {

ids <- sample(unique(dat$ID), replace = TRUE)

\# Proper cluster resample: duplicate whole IDs when drawn multiple times

boot <- dplyr::bind_rows(lapply(ids, function(i) dat[dat$ID == i, , drop = FALSE]))

m2 <- tryCatch(

stats::lm(stats::as.formula(paste(var, "~ GROUP * TP")), data = boot),

error = function(e) NULL

)

if (is.null(m2)) return(rep(NA_real_, 3))

emm2 <- tryCatch(emmeans::emmeans(m2, ~ GROUP:TP), error = function(e) NULL)

if (is.null(emm2)) return(rep(NA_real_, 3))

\# Recompute the reference cell within the bootstrap sample

ref2 <- which(emm2@grid$GROUP == "CON" & emm2@grid$TP == "TP1")

if (length(ref2) != 1) return(rep(NA_real_, 3))

r2 <- tryCatch(emmeans::contrast(emm2, "trt.vs.ctrl", ref = ref2, adjust = "none"),

error = function(e) NULL)

if (is.null(r2)) return(rep(NA_real_, 3))

df2 <- as.data.frame(r2)

out <- df2$estimate[match(wanted, df2$contrast)]

if (any(is.na(out))) return(rep(NA_real_, 3))

out

})

boot_mat <- t(boot_mat)

boot_mat <- boot_mat[stats::complete.cases(boot_mat), , drop = FALSE]

if (nrow(boot_mat) == 0) {

warning("No successful bootstrap samples for ", var)

return(tibble::tibble())

}

colnames(boot_mat) <- wanted

boot <- tibble::tibble(

contrast = colnames(boot_mat),

boot_mean = apply(boot_mat, 2, mean),

boot_se	= apply(boot_mat, 2, stats::sd),

boot_lo	= apply(boot_mat, 2, stats::quantile, probs = 0.025),

boot_hi	= apply(boot_mat, 2, stats::quantile, probs = 0.975),

n_success = nrow(boot_mat)

)

dplyr::left_join(param, boot, by = "contrast")

}

\# === Helper: Means & SD by GROUP × TP ===

mean_sd_by_group_tp <- function(dat, var) {

dat |>

dplyr::group_by(GROUP, TP) |>

dplyr::summarise(

Mean = mean(.data[[var]], na.rm = TRUE),

SD	= stats::sd(.data[[var]], na.rm = TRUE),

n	= dplyr::n(),

.groups = "drop"

) |>

dplyr::arrange(GROUP, TP)

}

\# === Examples (SF-36 dataset) ===

set.seed(123)

boot_trk_vs_con("PCS", dat = sf36_discrete_scored)

boot_trk_vs_con("MCS", dat = sf36_discrete_scored)

cat("\n--- Means & SD by GROUP × TP (PCS) ---\n")

print(mean_sd_by_group_tp(sf36_discrete_scored, "PCS"))

cat("\n--- Means & SD by GROUP × TP (MCS) ---\n")

print(mean_sd_by_group_tp(sf36_discrete_scored, "MCS"))

\# Pegboard ####

library(emmeans)

library(dplyr)

library(tibble)

boot_trk_vs_con <- function(var, dat = trackcontrol_pegboard_discrete_final, nsim = 1000) {

wanted <- c("TRACK TP1 - CON TP1","TRACK TP2 - CON TP1","TRACK TP3 - CON TP1")

\## Parametric (reference) using plain LM

m	<- lm(as.formula(paste(var, "~ GROUP * TP")), data = dat)

emm <- emmeans::emmeans(m, ~ GROUP:TP)

ref <- which(emm@grid$GROUP == "CON" & emm@grid$TP == "TP1")

param <- as.data.frame(

summary(emmeans::contrast(emm, "trt.vs.ctrl", ref = ref), infer = c(TRUE, TRUE))

) %>%

dplyr::filter(contrast %in% wanted) %>%

dplyr::select(contrast, estimate, SE, df, lower.CL, upper.CL, t.ratio, p.value)

\## Nonparametric cluster bootstrap (resample IDs WITH replacement)

boot_mat <- replicate(nsim, {

ids <- sample(unique(dat$ID), replace = TRUE)

boot <- dplyr::bind_rows(lapply(ids, function(i) dat[dat$ID == i, , drop = FALSE]))

m2 <- tryCatch(lm(as.formula(paste(var, "~ GROUP * TP")), data = boot), error = function(e) NULL)

if (is.null(m2)) return(rep(NA_real_, 3))

emm2 <- tryCatch(emmeans::emmeans(m2, ~ GROUP:TP), error = function(e) NULL)

if (is.null(emm2)) return(rep(NA_real_, 3))

\# IMPORTANT: recompute the reference cell for the bootstrap sample

ref2 <- which(emm2@grid$GROUP == "CON" & emm2@grid$TP == "TP1")

if (length(ref2) != 1) return(rep(NA_real_, 3))

r2 <- tryCatch(emmeans::contrast(emm2, "trt.vs.ctrl", ref = ref2, adjust = "none"),

error = function(e) NULL)

if (is.null(r2)) return(rep(NA_real_, 3))

df2 <- as.data.frame(r2)

out <- df2$estimate[match(wanted, df2$contrast)]

if (any(is.na(out))) return(rep(NA_real_, 3))

out

})

boot_mat <- t(boot_mat)

boot_mat <- boot_mat[stats::complete.cases(boot_mat), , drop = FALSE]

if (nrow(boot_mat) == 0) {

warning("No successful bootstrap samples for ", var)

return(tibble::tibble())

}

colnames(boot_mat) <- wanted

boot <- tibble::tibble(

contrast = colnames(boot_mat),

boot_mean = apply(boot_mat, 2, mean),

boot_se	= apply(boot_mat, 2, sd),

boot_lo	= apply(boot_mat, 2, stats::quantile, probs = 0.025),

boot_hi	= apply(boot_mat, 2, stats::quantile, probs = 0.975),

n_success = nrow(boot_mat)

)

dplyr::left_join(param, boot, by = "contrast")

}

\# Examples

set.seed(123)

boot_trk_vs_con("dominant")

boot_trk_vs_con("nondominant")

boot_trk_vs_con("simple_bilateral")

boot_trk_vs_con("assembly")

mean_sd_by_group_tp <- function(dat, var) {

dat %>%

group_by(GROUP, TP) %>%

summarise(

Mean = mean(.data[[var]], na.rm = TRUE),

SD	= sd(.data[[var]], na.rm = TRUE),

n	= dplyr::n(),

.groups = "drop"

) %>%

arrange(GROUP, TP)

}

\## Examples

cat("\n--- Means & SD by GROUP × TP (DOM) ---\n")

print(mean_sd_by_group_tp(trackcontrol_pegboard_discrete_final, "dominant"))

cat("\n--- Means & SD by GROUP × TP (NON) ---\n")

print(mean_sd_by_group_tp(trackcontrol_pegboard_discrete_final, "nondominant"))

cat("\n--- Means & SD by GROUP × TP (SIM) ---\n")

print(mean_sd_by_group_tp(trackcontrol_pegboard_discrete_final, "simple_bilateral"))

cat("\n--- Means & SD by GROUP × TP (ASS) ---\n")

print(mean_sd_by_group_tp(trackcontrol_pegboard_discrete_final, "assembly"))

\# TMS ####

\# SP PRE - change models

\# --- EMMs at SP, PRE (by GROUP) ---

emm_sp_pre <- emmeans(

TP3_bobyqa,

~ GROUP,

at	= list(STATE = "SP", BLOCK = "PRE"),

type = "link"

)

\# 1) GROUP estimates on the response scale (means & 95% CI)

sp_estimates <- summary(emm_sp_pre, type = "response") %>%

as.data.frame() %>%

transmute(

GROUP,

emmean = response, # estimated mean on response scale

lower = asymp.LCL,

upper = asymp.UCL,

SE, df

)

print(sp_estimates)

\# 2) TRACK vs CON contrast (ratio & % change)

group_contrast_sp_pre <- contrast(

emm_sp_pre,

method = "trt.vs.ctrl",

ref	= "CON",

adjust = "holm"

)

sp_contrast <- summary(group_contrast_sp_pre, type = "response", infer = c(TRUE, TRUE)) %>%

as.data.frame() %>%

mutate(

ratio	= if ("ratio" %in% names(.)) ratio else response,

percent_change = round((ratio - 1) * 100, 1)

) %>%

transmute(

contrast,

ratio,

lower = asymp.LCL,

upper = asymp.UCL,

SE, df, z.ratio, p.value,

percent_change

)

print(sp_contrast)

\# ICI PRE

\# --- Step 1: EMMs at PRE for each STATE within GROUP ---

emm_pre <- emmeans(

TP1_bobyqa,

~ STATE | GROUP,

at	= list(BLOCK = "PRE"),

type = "link"

)

\# --- Step 2: Within-group contrasts (LICI/SP, SICI/SP) ---

cont_ici_pre <- contrast(

emm_pre,

method = "trt.vs.ctrl",

ref	= "SP",

by	= "GROUP",

adjust = "none"

)

\# Get the within-group ratios on response scale

within_ratios <- summary(cont_ici_pre, type = "response", infer = c(TRUE, TRUE)) %>%

as.data.frame() %>%

mutate(

ratio = if ("ratio" %in% names(.)) ratio else response,

percent_change = round((ratio - 1) * 100, 1)

) %>%

select(GROUP, contrast, ratio, asymp.LCL, asymp.UCL, percent_change, SE, df, z.ratio, p.value)

print(within_ratios)

\# --- Step 3: Between-group ratio-of-ratios (TRACK vs CON for each inhibition ratio) ---

between_pre <- contrast(

cont_ici_pre,

method = "trt.vs.ctrl",

ref	= "CON",

by	= "contrast",

adjust = "holm"

)

out_pre <- summary(between_pre, type = "response", infer = c(TRUE, TRUE)) %>%

as.data.frame() %>%

mutate(

ratio = if ("ratio" %in% names(.)) ratio else response,

percent_change = round((ratio - 1) * 100, 1)

) %>%

select(contrast1, contrast, ratio, asymp.LCL, asymp.UCL, percent_change, SE, df, z.ratio, p.value)

print(out_pre)

\# PAS effects

emm_sp <- emmeans(TP3_bobyqa,

~ GROUP * BLOCK,

at = list(STATE = "SP"),

type = "link")

\# Contrast: GROUP × BLOCK, comparing each BLOCK to PRE

cont_sp <- contrast(emm_sp,

interaction = c("pairwise", "trt.vs.ctrl"),

ref = "PRE",

adjust = "holm")

\# Back-transform and add fold-change and percent-change columns

df_sp <- as.data.frame(cont_sp)

df_sp$fold_change_ratio <- round(exp(df_sp$estimate), 3)

df_sp$percent_change <- round((df_sp$fold_change_ratio - 1) * 100, 1)

\# View SP-only interaction results

df_sp

\# SP scores

emmeans(TP3_bobyqa, ~ GROUP | BLOCK,

at = list(STATE = "SP"),

type = "response")

\# ICI

emm_all <- emmeans(TP3_bobyqa,

~ GROUP * BLOCK * STATE,

type = "response")

cont_all <- contrast(emm_all,

interaction = c("pairwise", "trt.vs.ctrl", "trt.vs.ctrl"),

ref = c("PRE", "SP"),

adjust = "holm")

df_all <- as.data.frame(cont_all)

df_all$percent_change <- round((df_all$ratio - 1) * 100, 1)

print(df_all)

confint(cont_all)

\# Continuous analysis - Surveys

library(dplyr)

library(stringr)

library(lme4)

library(emmeans)

library(splines)

\# BSHS

bshs_vars <- c("physical_score", "generic_score")

\# number of items contributing to each raw sum

n_items <- c(physical_score = 9, generic_score = 21)

days_to_check <- c(23, 44, 82)

track_bshsscore <- track_bshsscore %>%

mutate(DAYS = as.numeric(DAYS)) %>%

filter(!is.na(ID), !is.na(DAYS))

results_list <- list()

estimates_list <- list()

comparisons_list <- list()

for (var in bshs_vars) {

df_use <- track_bshsscore %>%

filter(!is.na(.data[[var]])) %>%

transmute(

ID, DAYS,

\# convert raw sum (0–4 per item) to 0–100%

value = (.data[[var]] / (n_items[[var]] * 4)) * 100

)

\# Fit mixed model with natural spline for DAYS

model <- lmer(

value ~ ns(DAYS, df = 3) + (1 | ID),

data = df_use

)

results_list[[paste0(var, "_pct")]] <- model

\# EMMs at specified DAYS

emms <- emmeans(model, ~ DAYS, at = list(DAYS = days_to_check))

est_df <- as.data.frame(emms) %>%

mutate(Variable = paste0(var, "_pct"))

estimates_list[[paste0(var, "_pct")]] <- est_df

\# Pairwise (later - earlier), Holm-adjusted

contr_df <- contrast(emms, method = "revpairwise", adjust = "holm") %>%

as.data.frame() %>%

mutate(Variable = paste0(var, "_pct"))

\# Parse the "Later - Earlier" numeric day labels from contrast

day_labels <- str_extract_all(as.character(contr_df$contrast), "\\d+")

parsed_days <- do.call(rbind, lapply(day_labels, as.numeric))

colnames(parsed_days) <- c("Later", "Earlier")

\# Percent change = (Later - Earlier) / Earlier * 100

contr_df <- contr_df %>%

bind_cols(as.data.frame(parsed_days)) %>%

left_join(est_df %>% select(DAYS, emmean), by = c("Earlier" = "DAYS")) %>%

mutate(

contrast = paste(Later, Earlier, sep = " - "),

percent_change = (estimate / emmean) * 100

) %>%

select(contrast, estimate, SE, df, t.ratio, p.value, Variable, percent_change)

comparisons_list[[paste0(var, "_pct")]] <- contr_df

}

\# Combine results

estimates_all <- bind_rows(estimates_list)

comparisons_all <- bind_rows(comparisons_list)

\# Print

print(estimates_all)	# emmeans now 0–100%

print(comparisons_all)

\# --- Count participants and observations contributing to each BSHS variable ---

bshs_counts <- lapply(bshs_vars, function(var) {

df_use <- track_bshsscore %>%

filter(!is.na(.data[[var]]))

tibble(

Variable = var,

n_IDs = n_distinct(df_use$ID),

n_obs = nrow(df_use)

)

}) %>% bind_rows()

print(bshs_counts)

\# QuickDASH

days_to_check <- c(23, 44, 82)

qd_items <- paste0("quickdash", 1:11)

\# ---- 1) SCORE QUICKDASH (0–100) ----

track_quickdashv_scored <- track_quickdashv %>%

\# Coerce item columns to numeric and set out-of-range to NA

mutate(across(all_of(qd_items),

~ {x <- suppressWarnings(as.numeric(.)); ifelse(!is.na(x) & !(x %in% 1:5), NA_real_, x)})) %>%

rowwise() %>%

mutate(

quickdash_items_answered = sum(!is.na(c_across(all_of(qd_items)))),

quickdash_raw_mean = ifelse(

quickdash_items_answered >= 10,

mean(c_across(all_of(qd_items)), na.rm = TRUE),

NA_real_

),

\# Transform to 0–100: ((mean - 1)/4)*100

quickdash_score = ifelse(is.na(quickdash_raw_mean), NA_real_,

((quickdash_raw_mean - 1) / 4) * 100),

quickdash_valid = !is.na(quickdash_score)

) %>%

ungroup()

\# ---- 2) PREP DATA FOR MODEL ----

\# Keep just what we need and ensure DAYS numeric

quickdash_df <- track_quickdashv_scored %>%

mutate(DAYS = as.numeric(DAYS)) %>%

filter(!is.na(ID), !is.na(DAYS), quickdash_valid) %>%

select(ID, DAYS, value = quickdash_score)

\# ---- 3) FIT MIXED MODEL ----

qd_model <- lmer(

value ~ ns(DAYS, df = 3) + (1 | ID),

data = quickdash_df

)

\# ---- 4) EMMs at specified DAYS ----

qd_emms <- emmeans(qd_model, ~ DAYS, at = list(DAYS = days_to_check))

estimates_all <- as.data.frame(qd_emms) %>%

mutate(Variable = "quickdash_score") %>%

select(DAYS, emmean, SE, df, lower.CL, upper.CL, Variable)

\# ---- 5) Pairwise (later - earlier), Holm-adjusted ----

qd_contr <- contrast(qd_emms, method = "revpairwise", adjust = "holm") %>%

as.data.frame() %>%

mutate(Variable = "quickdash_score")

\# Parse numeric day labels from the contrast names

day_labels <- str_extract_all(as.character(qd_contr$contrast), "\\d+")

parsed_days <- do.call(rbind, lapply(day_labels, as.numeric))

colnames(parsed_days) <- c("Later", "Earlier")

\# Percent change = (Later - Earlier) / Earlier * 100

comparisons_all <- qd_contr %>%

bind_cols(as.data.frame(parsed_days)) %>%

left_join(estimates_all %>% select(DAYS, emmean), by = c("Earlier" = "DAYS")) %>%

mutate(

contrast = paste(Later, Earlier, sep = " - "),

percent_change = (estimate / emmean) * 100

) %>%

select(contrast, estimate, SE, df, t.ratio, p.value, Variable, percent_change)

\# ---- 6) Print ----

print(estimates_all)

print(comparisons_all)

\# ---- Report sample size info ----

qd_counts <- quickdash_df %>%

summarise(

n_IDs = n_distinct(ID),

n_obs = n()

)

print(qd_counts)

\# LLFI

days_to_check <- c(23, 44, 82)

\# --- Prep data: keep needed cols, ensure DAYS is numeric ---

llfiscore_use <- llfiscore %>%

mutate(DAYS = as.numeric(DAYS)) %>%

filter(!is.na(ID), !is.na(DAYS), !is.na(percentage_score)) %>%

select(ID, DAYS, value = percentage_score)

\# --- Fit mixed model with spline over DAYS ---

llfi_model <- lmer(

value ~ ns(DAYS, df = 3) + (1 | ID),

data = llfiscore_use

)

\# --- EMMs at specified DAYS ---

llfi_emms <- emmeans(llfi_model, ~ DAYS, at = list(DAYS = days_to_check))

estimates_all <- as.data.frame(llfi_emms) %>%

mutate(Variable = "LLFI percentage_score") %>%

select(DAYS, emmean, SE, df, lower.CL, upper.CL, Variable)

\# --- Pairwise (later - earlier), Holm-adjusted ---

llfi_contr <- contrast(llfi_emms, method = "revpairwise", adjust = "holm") %>%

as.data.frame() %>%

mutate(Variable = "LLFI percentage_score")

\# Parse numeric day labels and compute % change relative to Earlier day

day_labels <- str_extract_all(as.character(llfi_contr$contrast), "\\d+")

parsed_days <- do.call(rbind, lapply(day_labels, as.numeric))

colnames(parsed_days) <- c("Later", "Earlier")

comparisons_all <- llfi_contr %>%

bind_cols(as.data.frame(parsed_days)) %>%

left_join(estimates_all %>% select(DAYS, emmean), by = c("Earlier" = "DAYS")) %>%

mutate(

contrast = paste(Later, Earlier, sep = " - "),

\# percent change: (Later - Earlier) / Earlier * 100

percent_change = (estimate / emmean) * 100

) %>%

select(contrast, estimate, SE, df, t.ratio, p.value, Variable, percent_change)

\# --- Output ---

print(estimates_all)

print(comparisons_all)

llfi_counts <- llfiscore %>%

mutate(DAYS = as.numeric(DAYS)) %>%

filter(!is.na(ID), !is.na(DAYS), !is.na(percentage_score)) %>%

summarise(

n_IDs = n_distinct(ID),

n_obs = n()

)

print(llfi_counts)

\# POSAS

library(dplyr)

library(lme4)

library(emmeans)

library(splines)

library(stringr)

days_to_check <- c(23, 44, 82)

\# Use posasscore_mean directly

posas_df <- posasscore_mean %>%

mutate(DAYS = as.numeric(DAYS)) %>%	# already numeric, harmless

filter(!is.na(ID), !is.na(DAYS), !is.na(score_pct)) %>%

select(ID, DAYS, value = score_pct)

\# Mixed model

posas_model <- lmer(

value ~ ns(DAYS, df = 3) + (1 | ID),

data = posas_df

)

\# EMMs at the target days

posas_emms <- emmeans(posas_model, ~ DAYS, at = list(DAYS = days_to_check))

estimates_all <- as.data.frame(posas_emms) %>%

mutate(Variable = "posas_score_pct")

\# Pairwise contrasts (later - earlier), Holm-adjusted

contr_df <- contrast(posas_emms, method = "revpairwise", adjust = "holm") %>%

as.data.frame() %>%

mutate(Variable = "posas_score_pct")

\# Add % change relative to the earlier day

day_labels <- str_extract_all(as.character(contr_df$contrast), "\\d+")

parsed_days <- do.call(rbind, lapply(day_labels, as.numeric))

colnames(parsed_days) <- c("Later", "Earlier")

comparisons_all <- contr_df %>%

bind_cols(as.data.frame(parsed_days)) %>%

left_join(estimates_all %>% select(DAYS, emmean), by = c("Earlier" = "DAYS")) %>%

mutate(

contrast = paste(Later, Earlier, sep = " - "),

percent_change = (estimate / emmean) * 100

) %>%

select(contrast, estimate, SE, df, t.ratio, p.value, Variable, percent_change)

print(estimates_all)

print(comparisons_all)

\# --- ID and observation counts ---

posas_counts <- posas_df %>%

summarise(

n_IDs = n_distinct(ID),

n_obs = n()

)

print(posas_counts)

\# painDETECT

library(dplyr)

library(lme4)

library(emmeans)

library(splines)

library(stringr)

days_to_check <- c(23, 44, 82)

\# 1) Build painDETECT total (raw sum; NA = no pain -> 0 is fine per your setup)

paindetect_df <- track_pain %>%

mutate(

pd_radiating = ifelse(is.na(pd_radiating), 0, pd_radiating),

pd_course	= ifelse(is.na(pd_course), 0, pd_course),

across(pd_1:pd_7, ~ ifelse(is.na(.), 0, .)),

\# Official recodes

pd_radiating_recode = ifelse(pd_radiating == 1, 2, 0), # 1→+2, else 0

pd_course_recode = case_when(

pd_course == 1 ~ 0, # persistent w/ slight fluctuations

pd_course == 2 ~ -1, # persistent w/ pain attacks

pd_course %in% c(3,4) ~ 1,

TRUE ~ 0

),

total_score = pd_radiating_recode + pd_course_recode +

pd_1 + pd_2 + pd_3 + pd_4 + pd_5 + pd_6 + pd_7

) %>%

mutate(DAYS = as.numeric(DAYS)) %>%

filter(!is.na(ID), !is.na(DAYS)) %>%

select(ID, DAYS, value = total_score)

\# 2) Mixed model (random intercept for ID; spline over DAYS)

pd_model <- lmer(

value ~ ns(DAYS, df = 3) + (1 | ID),

data = paindetect_df

)

\# 3) EMMs at 23/44/82

pd_emms <- emmeans(pd_model, ~ DAYS, at = list(DAYS = days_to_check))

estimates_all <- as.data.frame(pd_emms) %>%

mutate(Variable = "painDETECT_total")

\# 4) Pairwise contrasts (later - earlier), Holm-adjusted

contr_df <- contrast(pd_emms, method = "revpairwise", adjust = "holm") %>%

as.data.frame() %>%

mutate(Variable = "painDETECT_total")

\# 5) Relative % change vs earlier day (optional)

day_labels <- str_extract_all(as.character(contr_df$contrast), "\\d+")

parsed_days <- do.call(rbind, lapply(day_labels, as.numeric))

colnames(parsed_days) <- c("Later", "Earlier")

comparisons_all <- contr_df %>%

bind_cols(as.data.frame(parsed_days)) %>%

left_join(estimates_all %>% select(DAYS, emmean), by = c("Earlier" = "DAYS")) %>%

mutate(

contrast = paste(Later, Earlier, sep = " - "),

percent_change = (estimate / emmean) * 100

) %>%

select(contrast, estimate, SE, df, t.ratio, p.value, Variable, percent_change)

print(estimates_all)	# emmeans on raw painDETECT scale (-1..38)

print(comparisons_all)	# contrasts; percent_change is relative to earlier Mean

\# --- ID and observation counts ---

pd_counts <- paindetect_df %>%

summarise(

n_IDs = n_distinct(ID),

n_obs = n()

)

print(pd_counts)

\# Sensory Tests

\# VAS

days_to_check <- c(23, 44, 82)

\# --- Prep data: select VAS ---

vas_df <- track_sensory_results %>%

mutate(DAYS = as.numeric(DAYS)) %>%	# ensure numeric

filter(!is.na(ID), !is.na(DAYS), !is.na(vas)) %>%

select(ID, DAYS, value = vas)

\# --- Sample size info ---

n_IDs	<- n_distinct(vas_df$ID)

n_obs	<- nrow(vas_df)

cat("Included in analysis:", n_IDs, "unique participants,", n_obs, "total observations\n")

\# --- Fit mixed model (random intercept for ID; spline for DAYS) ---

vas_model <- lmer(

value ~ ns(DAYS, df = 3) + (1 | ID),

data = vas_df

)

\# --- Estimated marginal means at 23, 44, 82 days ---

vas_emms <- emmeans(vas_model, ~ DAYS, at = list(DAYS = days_to_check))

estimates_all <- as.data.frame(vas_emms) %>%

mutate(Variable = "VAS_score")

\# --- Pairwise contrasts (later - earlier), Holm adjusted ---

contr_df <- contrast(vas_emms, method = "revpairwise", adjust = "holm") %>%

as.data.frame() %>%

mutate(Variable = "VAS_score")

\# --- Parse contrasts and compute percent change ---

day_labels <- str_extract_all(as.character(contr_df$contrast), "\\d+")

parsed_days <- do.call(rbind, lapply(day_labels, as.numeric))

colnames(parsed_days) <- c("Later", "Earlier")

comparisons_all <- contr_df %>%

bind_cols(as.data.frame(parsed_days)) %>%

left_join(estimates_all %>% select(DAYS, emmean), by = c("Earlier" = "DAYS")) %>%

mutate(

contrast = paste(Later, Earlier, sep = " - "),

percent_change = (estimate / emmean) * 100

) %>%

select(contrast, estimate, SE, df, t.ratio, p.value, Variable, percent_change)

\# --- Output ---

print(estimates_all)	# model-predicted VAS means at 23/44/82 days

print(comparisons_all)

\# Grip strength

days_to_check <- c(23, 44, 82)

\# Variables to analyze

grip_vars <- c("grip_m", "grip_l")

results_list <- list()

estimates_list <- list()

comparisons_list <- list()

for (var in grip_vars) {

\# --- Prep data ---

grip_df <- track_sensory_results %>%

mutate(DAYS = as.numeric(DAYS)) %>%

filter(!is.na(ID), !is.na(DAYS), !is.na(.data[[var]])) %>%

select(ID, DAYS, value = all_of(var))

\# --- Sample size info ---

n_IDs <- n_distinct(grip_df$ID)

n_obs <- nrow(grip_df)

cat("Variable:", var, "|", n_IDs, "unique participants,", n_obs, "total observations\n")

\# --- Fit mixed model (random intercept for ID; spline for DAYS) ---

grip_model <- lmer(

value ~ ns(DAYS, df = 3) + (1 | ID),

data = grip_df

)

results_list[[var]] <- grip_model

\# --- Estimated marginal means at 23, 44, 82 days ---

grip_emms <- emmeans(grip_model, ~ DAYS, at = list(DAYS = days_to_check))

est_df <- as.data.frame(grip_emms) %>%

mutate(Variable = var)

estimates_list[[var]] <- est_df

\# --- Pairwise contrasts (later - earlier), Holm adjusted ---

contr_df <- contrast(grip_emms, method = "revpairwise", adjust = "holm") %>%

as.data.frame() %>%

mutate(Variable = var)

\# --- Parse contrasts and compute percent change ---

day_labels <- str_extract_all(as.character(contr_df$contrast), "\\d+")

parsed_days <- do.call(rbind, lapply(day_labels, as.numeric))

colnames(parsed_days) <- c("Later", "Earlier")

contr_df <- contr_df %>%

bind_cols(as.data.frame(parsed_days)) %>%

left_join(est_df %>% select(DAYS, emmean), by = c("Earlier" = "DAYS")) %>%

mutate(

contrast = paste(Later, Earlier, sep = " - "),

percent_change = (estimate / emmean) * 100

) %>%

select(contrast, estimate, SE, df, t.ratio, p.value, Variable, percent_change)

comparisons_list[[var]] <- contr_df

}

\# --- Combine results ---

estimates_all	<- bind_rows(estimates_list)

comparisons_all <- bind_rows(comparisons_list)

\# --- Output ---

print(estimates_all)

print(comparisons_all)

\# PPT

days_to_check <- c(23, 44, 82)

\# Variables to analyze

pain_vars <- c("pain_m1", "pain_m2", "pain_mbh",

"pain_l1", "pain_l2", "pain_lbh", "pain_fh")

results_list <- list()

estimates_list <- list()

comparisons_list <- list()

for (var in pain_vars) {

\# --- Prep data ---

pain_df <- track_sensory_results %>%

mutate(DAYS = as.numeric(DAYS)) %>%

filter(!is.na(ID), !is.na(DAYS), !is.na(.data[[var]])) %>%

select(ID, DAYS, value = all_of(var))

\# --- Sample size info ---

n_IDs <- n_distinct(pain_df$ID)

n_obs <- nrow(pain_df)

cat("Variable:", var, "|", n_IDs, "unique participants,", n_obs, "total observations\n")

\# --- Fit mixed model ---

pain_model <- lmer(

value ~ ns(DAYS, df = 3) + (1 | ID),

data = pain_df

)

results_list[[var]] <- pain_model

\# --- Estimated marginal means ---

pain_emms <- emmeans(pain_model, ~ DAYS, at = list(DAYS = days_to_check))

est_df <- as.data.frame(pain_emms) %>%

mutate(Variable = var)

estimates_list[[var]] <- est_df

\# --- Pairwise contrasts (Holm adjusted) ---

contr_df <- contrast(pain_emms, method = "revpairwise", adjust = "holm") %>%

as.data.frame() %>%

mutate(Variable = var)

\# --- Parse contrasts and compute % change ---

day_labels <- str_extract_all(as.character(contr_df$contrast), "\\d+")

parsed_days <- do.call(rbind, lapply(day_labels, as.numeric))

colnames(parsed_days) <- c("Later", "Earlier")

contr_df <- contr_df %>%

bind_cols(as.data.frame(parsed_days)) %>%

left_join(est_df %>% select(DAYS, emmean), by = c("Earlier" = "DAYS")) %>%

mutate(

contrast = paste(Later, Earlier, sep = " - "),

percent_change = (estimate / emmean) * 100

) %>%

select(contrast, estimate, SE, df, t.ratio, p.value, Variable, percent_change)

comparisons_list[[var]] <- contr_df

}

\# --- Combine results ---

estimates_all	<- bind_rows(estimates_list)

comparisons_all <- bind_rows(comparisons_list)

\# --- Output ---

print(estimates_all)

print(comparisons_all)

\# Monofilament

library(tidyverse)

library(lme4)

library(emmeans)

library(splines)

library(stringr)

days_to_check <- c(23, 44, 82)

\# Variables to analyze

mono_vars <- c("mono_m1", "mono_m2", "mono_l1", "mono_l2")

results_list	<- list()

estimates_list	<- list()

comparisons_list <- list()

for (var in mono_vars) {

\# --- Prep data ---

mono_df <- track_sensory_results %>%

mutate(DAYS = as.numeric(DAYS)) %>%

filter(!is.na(ID), !is.na(DAYS), !is.na(.data[[var]])) %>%

select(ID, DAYS, value = all_of(var))

\# --- Quick info ---

n_IDs <- n_distinct(mono_df$ID); n_obs <- nrow(mono_df)

cat("Variable:", var, "|", n_IDs, "unique participants,", n_obs, "total observations\n")

\# --- Fit mixed model on log10 scale ---

mono_model <- lmer(

log10(value) ~ ns(DAYS, df = 3) + (1 | ID),

data = mono_df

)

results_list[[var]] <- mono_model

\# --- EMMs at 23/44/82 (compute on log10 scale; back-transform to original) ---

mono_emms <- emmeans(mono_model, ~ DAYS, at = list(DAYS = days_to_check))

est_df <- as.data.frame(mono_emms) %>%

mutate(

Variable	= var,

emmean_bt	= 10^emmean, # back-transformed mean

lower.CL_bt = 10^lower.CL, # back-transformed CI

upper.CL_bt = 10^upper.CL

)

estimates_list[[var]] <- est_df

\# --- Pairwise contrasts (Holm adjusted) on log10 scale ---

contr_df <- contrast(mono_emms, method = "revpairwise", adjust = "holm") %>%

as.data.frame() %>%

mutate(Variable = var)

\# --- Parse contrast labels + % change on ORIGINAL scale ---

day_labels <- str_extract_all(as.character(contr_df$contrast), "\\d+")

parsed_days <- do.call(rbind, lapply(day_labels, as.numeric))

colnames(parsed_days) <- c("Later", "Earlier")

contr_df <- contr_df %>%

bind_cols(as.data.frame(parsed_days)) %>%

\# On log10 scale, 'estimate' is a difference in logs -> ratio on original scale = 10^estimate

mutate(

ratio	= 10^estimate, # Later / Earlier on original scale

percent_change = (ratio - 1) * 100, # % change on original scale

contrast	= paste(Later, Earlier, sep = " - ")

) %>%

select(contrast, estimate, SE, df, t.ratio, p.value, Variable, ratio, percent_change)

comparisons_list[[var]] <- contr_df

}

\# --- Combine results ---

estimates_all	<- bind_rows(estimates_list)

comparisons_all <- bind_rows(comparisons_list)

\# --- Outputs (back-transformed EMMs + original-scale ratios) ---

print(

estimates_all %>%

select(Variable, DAYS, emmean_bt, lower.CL_bt, upper.CL_bt)

)

print(

comparisons_all %>%

select(Variable, contrast, ratio, percent_change, t.ratio, p.value)

)

\# Neuropen

days_to_check <- c(23, 44, 82)

\# --- Variables to analyze ---

neuro_vars <- c(

"neuro_dull1_m1","neuro_dull5_m1","neuro_sharp1_m1","neuro_sharp5_m1",

"neuro_dull1_m2","neuro_dull5_m2","neuro_sharp1_m2","neuro_sharp5_m2",

"neuro_dull1_l1","neuro_dull5_l1","neuro_sharp1_l1","neuro_sharp5_l1",

"neuro_dull1_l2","neuro_dull5_l2","neuro_sharp1_l2","neuro_sharp5_l2"

)

results_list <- list()

estimates_list <- list()

comparisons_list <- list()

for (var in neuro_vars) {

\# --- Prep data ---

neuro_df <- track_sensory_results %>%

dplyr::mutate(DAYS = as.numeric(DAYS)) %>%

dplyr::filter(!is.na(ID), !is.na(DAYS), !is.na(.data[[var]])) %>%

dplyr::select(ID, DAYS, value = dplyr::all_of(var))

\# --- Observational summary (per variable) ---

n_IDs <- dplyr::n_distinct(neuro_df$ID)

n_obs <- nrow(neuro_df)

cat("Variable:", var, "|", n_IDs, "unique participants,", n_obs, "total observations\n")

\# --- Fit mixed model (random intercept for ID; spline for DAYS) ---

neuro_model <- lme4::lmer(

value ~ splines::ns(DAYS, df = 3) + (1 | ID),

data = neuro_df

)

results_list[[var]] <- neuro_model

\# --- Estimated marginal means at 23/44/82 days ---

neuro_emms <- emmeans::emmeans(neuro_model, ~ DAYS, at = list(DAYS = days_to_check))

est_df <- as.data.frame(neuro_emms) %>%

dplyr::mutate(Variable = var)

estimates_list[[var]] <- est_df

\# --- Pairwise contrasts (later - earlier), Holm adjusted ---

contr_df <- emmeans::contrast(neuro_emms, method = "revpairwise", adjust = "holm") %>%

as.data.frame() %>%

dplyr::mutate(Variable = var)

\# --- Parse contrasts and compute % change vs earlier day ---

day_labels <- stringr::str_extract_all(as.character(contr_df$contrast), "\\d+")

parsed_days <- do.call(rbind, lapply(day_labels, as.numeric))

colnames(parsed_days) <- c("Later", "Earlier")

contr_df <- contr_df %>%

dplyr::bind_cols(as.data.frame(parsed_days)) %>%

dplyr::left_join(est_df %>% dplyr::select(DAYS, emmean), by = c("Earlier" = "DAYS")) %>%

dplyr::mutate(

contrast = paste(Later, Earlier, sep = " - "),

percent_change = (estimate / emmean) * 100

) %>%

dplyr::select(contrast, estimate, SE, df, t.ratio, p.value, Variable, percent_change)

comparisons_list[[var]] <- contr_df

}

\# --- Combine results ---

estimates_all <- dplyr::bind_rows(estimates_list)

comparisons_all <- dplyr::bind_rows(comparisons_list)

\# --- Output ---

print(estimates_all)

print(comparisons_all)

\# Brush

\# --- Variables to summarise ---

brush_vars <- c("brush_m1", "brush_m2", "brush_l1", "brush_l2")

\# --- Summarise frequencies & percentages ---

brush_summary <- track_sensory_results %>%

select(ID, DAYS, time_window, all_of(brush_vars)) %>%

pivot_longer(cols = all_of(brush_vars),

names_to = "Variable",

values_to = "Response") %>%

filter(!is.na(Response)) %>%

group_by(Variable, time_window, Response) %>%

summarise(n = n(), .groups = "drop") %>%

group_by(Variable, time_window) %>%

mutate(

total = sum(n),

percent = round((n / total) * 100, 1)

) %>%

arrange(Variable, time_window, Response)

\# --- Print summary ---

print(as.data.frame(brush_summary))

\# Load ####

\# Load new environment

load("TS_analysis_objects_v1.RData")
